# Leisure time sedentary behaviour and risks of breast, colorectal, and prostate cancer: A Mendelian randomization analysis

**DOI:** 10.1101/2023.03.01.23286492

**Authors:** Nikos Papadimitriou, Nabila Kazmi, Niki Dimou, Konstantinos K Tsilidis, Richard M Martin, Sarah J Lewis, Brigid M Lynch, Michael Hoffmeister, Sun-Seog Kweon, Li Li, Roger L Milne, Lori C Sakoda, Robert E Schoen, Amanda I Phipps, Jane C Figueiredo, Ulrike Peters, Suzanne C. Dixon-Suen, Marc J Gunter, Neil Murphy

## Abstract

Sedentary behaviours have been associated with increased risks of some common cancers in epidemiological studies; however, it is unclear if these associations are causal. We examined potential causal associations between self-reported leisure television watching and computer use and risks of breast, colorectal, and prostate cancer using a two-sample Mendelian randomization framework. Genetic variants were identified from a recent genome-wide association study (GWAS). Cancer data were obtained from cancer GWAS consortia. Additional sensitivity analyses were applied to examine the robustness of the results. A 1-standard deviation increment in hours of television watching increased risk of breast (OR: 1.15, 95% confidence interval [CI]: 1.05,1.26) and colorectal cancer (OR: 1.32, 95%CI: 1.16,1.49) with little evidence of an association for prostate cancer risk. In multivariable models adjusted for years of education, the effect estimates for television watching were attenuated (breast cancer, OR: 1.08, 95%CI: 0.92,1.27; colorectal cancer, OR: 1.08, 95%CI: 0.90,1.31). Post-hoc analyses showed that years of education might have a possible confounding and mediating role in the association between television watching with breast and colorectal cancer. Consistent results were observed by sex (colorectal cancer), anatomical subsites, and cancer subtypes. There was little evidence of associations between computer use and cancer risk. We found evidence of positive associations between hours of television watching and risks of breast and colorectal cancer. However, these findings should be interpreted cautiously given the complex role of education. Future studies using objective measures of exposure can provide new insights into the possible role of sedentary behaviour in cancer development.

**Novelty and impact:** Evidence from observational studies that examined associations between sedentary behaviours and common cancers is mixed and causality is uncertain. In our Mendelian randomization analyses, higher levels of leisure television watching were found to increase the risks of breast and colorectal cancer, suggesting that the that the promotion of lowering sedentary behaviour time could be an effective strategy in the primary prevention of these commonly diagnosed cancers.

**Article category:** Cancer Epidemiology

## Introduction

Breast, colorectal, and prostate cancer are three of the most common malignancies collectively accounting for an estimated 29% of new cancer cases in 2020 (1). Sedentary behaviour is defined as any waking behaviour characterized by energy expenditure ≤1.5 metabolic equivalents while in a sitting, reclining, or lying posture (2). The most common sedentary activities are television watching and computer use; these are more accurately recalled than total sedentary time and are therefore commonly used as surrogates of sedentary behaviour (3). A recent US study reported that approximately two-thirds of adults spent two or more hours each day watching television and around 50% spend more than one hour using their computer outside work (4). Studies in the UK and in the US estimated that adults on average spend five to six hours per day sitting (4, 5). Given such a high prevalence, sedentary behaviours represent an important public health challenge as they have been linked with multiple adverse health outcomes (6, 7).

Numerous observational studies have examined the associations between sedentary behaviours and the risks of breast, colorectal, and prostate cancer (8). A meta-analysis of case-control and cohort studies reported that sedentary behaviour was not associated with colorectal cancer risk (8). More recently, however, a UK Biobank analysis, found that greater volumes of television watching was associated with elevated colon cancer risk (9).The aforementioned meta-analysis did not observe any significant associations between sedentary behaviour and risk of prostate cancer (8). For breast cancer, when the meta-analysis included cohort studies only, sedentary behaviour was associated with a higher breast cancer risk (8). Clarifying causal associations from such observational evidence is hampered by inherent biases of the study design, such as residual confounding and reverse causality (10–12). Mendelian randomization (MR) is an alternative way to investigate potential causal associations. MR uses germline genetic variants as proxies (or instrumental variables) for exposures of interest to make causal inferences between an exposure and an outcome (13). Unlike traditional observational epidemiology, MR can be largely free of conventional confounding owing to the random independent assignment of alleles during meiosis (14). In addition, multivariable MR methods have been developed to adjust for confounding if found to be present. There should be no reverse causation in MR studies, as germline genetic variants are fixed at conception and are consequently unaffected by the disease process (14). A recent MR analysis reported a positive effect estimate for television watching with lung cancer risk (15). However, similar analyses investigating possible causal effects of sedentary behaviours for other common cancers have not been conducted.

We used a two-sample MR framework to examine potential causal associations between self-reported sedentary behaviours and risks of breast, colorectal, and prostate cancer. Genetic variants associated with leisure television watching and computer use were identified from a recent genome-wide association study (GWAS) (16) and we then examined how these genetic variants related to risks of breast, colorectal, and prostate cancer using large-scale GWAS consortia data (17–19).

## Materials and Methods

### Data on leisure sedentary behaviours

Summary-level data on duration of leisure sedentary behaviours were obtained from a recently published GWAS conducted in 408,815 participants of European ancestry from the UK Biobank using BOLT-LMM v2.3beta2, using a mixed linear model correcting for population structure and cryptic relatedness (16). To ascertain the duration of the sedentary behaviours, participants within the UK Biobank were asked three questions, “In a typical DAY, how many hours do you spend watching television?”, “In a typical DAY, how many hours do you spend using the computer? (Do not include using a computer at work)” and “In a typical DAY, how many hours do you spend driving?”(16). This GWAS identified 209 and 52 genome-wide-significant single nucleotide polymorphisms (SNPs) (P-value < 5×10^-8^) for leisure television watching and computer use respectively using a linkage disequilibrium (LD) of R^2^ < 0.005 within a five megabase window (Supplemental Tables 1-2). The GWAS also identified five genetic variants associated with driving; however, we did not include these instruments in our MR analyses due to low statistical power (see Statistical power, below). The 261 SNPs included in both instruments were identified in 204 loci demonstrating a partial overlap between the two phenotypes with 22 common loci. The selected SNPs explained approximately 2% and 0.5% of the variability in television watching and computer use respectively.

### Data on breast, colorectal, and prostate cancer

Summary data for the associations of the above genetic variants with breast cancer were obtained from a GWAS of 247,173 women (133,384 breast cancer cases and 113,789 controls) of European ancestry from the Breast Cancer Association Consortium (19). We included six related outcomes in our analyses (overall, luminal A, luminal B, luminal B HER2 negative, HER2 enriched, and triple negative breast cancer).

For colorectal cancer, summary data from 98,715 participants (52,775 colorectal cancer cases and 45,940 controls) were drawn from a meta-analysis within the ColoRectal Transdisciplinary Study, the Colon Cancer Family Registry, and the Genetics and Epidemiology of Colorectal Cancer consortia (17). We included five outcomes in our analyses (overall colorectal cancer, colorectal cancer for men, colorectal cancer for women, colon cancer, and rectal cancer). The summary statistics did not include UK Biobank study to avoid potential overlap with the leisure sedentary behaviours GWAS.

For prostate cancer, summary data from a meta-analysis of 140,254 (79,148 prostate cancer cases and 61,106 controls) men of European ancestry in the Prostate Cancer Association Group to Investigate Cancer-Associated Alterations in the Genome and the Genetic Associations and Mechanisms in Oncology/Elucidating Loci Involved in Prostate Cancer Susceptibility consortia (18). The same consortia also conducted a GWAS of aggressive prostate cancer involving 15,167 cases and 58,308 controls, in which cancer cases were defined as aggressive based on the following characteristics: Gleason score ≥8, Prostate-Specific Antigen (PSA)>100 ng/mL, metastatic disease (M1) or death from prostate cancer (18).

All cancer estimates for the two exposures of interest are provided in Supplemental Tables 3-8. All participants provided written informed consent. Ethics were approved by respective institutional review boards.

### Statistical power

The statistical power was calculated *a priori* using an online tool at http://cnsgenomics.com/shiny/mRnd/ (20). Under the scenario of a type 1 error of 5%, for leisure television use an expected OR per 1 standard deviation (SD) ≥ 1.09, ≥ 1.14 and ≥ 1.11 was needed to have adequate statistical power (> 80%) for overall breast, colorectal and prostate cancer respectively. Supplemental Table 9 presents the power estimates for the three exposures of interest by subtypes or subsites of breast, colorectal, and prostate cancer.

### Statistical analysis

A two-sample MR approach using summary data and the fixed-effect IVW method was implemented. All results correspond to an OR per 1-SD increment in genetically-predicted hours of leisure sedentary behaviour (television watching: 1.5 hours/day; computer use: 1.2 hours/day). The heterogeneity of the causal estimates by cancer subtype (breast cancer), subsite (colorectal cancer) and sex (colorectal cancer only) was investigated by calculating the I^2^ metric using a fixed effect meta-analysis model (21).

### Sensitivity analyses

MR studies have three main assumptions that must be satisfied in order for their causal estimates to be valid, which in the context of this study are: 1) the genetic instrument is strongly associated with the levels of exposure (sedentary behaviour); 2) the genetic instrument is not associated with any potential confounder of the exposure (sedentary behaviour)—outcome (cancer) association; and 3) the genetic instrument does not affect the outcome (cancer) independently of the exposure (sedentary behaviour) (i.e. exclusion of horizontal pleiotropy). The strength of each genetic instrument can be evaluated through the F-statistic (provided by the initial GWAS) (16). Several sensitivity analyses were conducted to identify and correct for the presence of horizontal pleiotropy in the results from the main analysis. Cochran’s Q was computed to quantify heterogeneity across the individual causal effects, with a P-value ≤ 0.05 indicating the presence of pleiotropy, and consequently, a random effects IVW MR analysis was used (21, 22). MR-Egger regression was performed in which the intercept term can deviate from zero allowing estimation of the causal effect even in the presence of invalid genetic variants. Large deviations from zero represent the presence of horizontal pleiotropic effects across the genetic variants. In such a case, the slope of the MR-Egger regression provides valid MR estimates when the pleiotropic effects of the genetic variants are independent from the genetic associations with the exposure (23, 24). Moreover, causal estimates were also computed using the weighted-median method that can give valid MR estimates under the presence of horizontal pleiotropy when up to 50% of the included instruments are invalid (25). The MR pleiotropy residual sum and outlier test (MR-PRESSO) was also used to assess the presence of pleiotropy. The MR-PRESSO test relies on a regression framework to identify outlying genetic variants which may potentially be pleiotropic, we then reran the analysis after excluding these outlying variants (26). We also examined the selected genetic instruments and their proxies (r^2^ > 0.8) and their associations with secondary phenotypes (P-value < 5 × 10−8) in populations of European descent in Phenoscanner (http://www.phenoscanner.medschl.cam.ac.uk/) to explore potential pleiotropy of the included SNPs. Since several of the genetic variants were also associated with adiposity or education-related phenotypes - such as body mass index (BMI) and educational attainment - we performed multivariable MR to investigate whether any initial significant associations for sedentary behaviour are confounded by these two traits as well as additional secondary traits such as lifetime smoking and alcohol consumption which have previously been linked with cancer risk (27–29). For BMI, summary data from a GWAS meta-analysis of about 700,000 participants of European descent within the Genetic Investigation of ANthropometric Traits (GIANT) consortium and UK Biobank was obtained (30). For years of educational attainment, we obtained summary level data from a published GWAS of 1.1 million participants of European descent within the Social Science Genetic Association Consortium and which measured the number of completed years of schooling among those individuals (31). Data on alcohol consumption (drinks per week) was drawn from a GWAS of 1.2 million individuals (32). The data for lifetime smoking was obtained from a recent GWAS and MR study on causal effects of lifetime smoking on risk for depression and schizophrenia (33). In the current analysis we used data of 766,345 participants which is publicly available. All relevant summary statistics for the multivariable MR analyses is given in supplemental tables 10-17. For multivariable MR, we also calculated two variables: the conditional F_early life body size_ F_adult body size_ which can be used to examine how much variance the genetic variants explain on the main (sedentary behaviours) and secondary exposures (e.g., years of education); F values over 10 suggest little evidence of weak instrument bias (34). Finally, as a post-hoc analysis based on the results from the multivariable MR and trying to understand the observed attenuation, we also conducted a bidirectional MR study to examine the associations between sedentary behaviours and the four secondary traits (BMI, years of education, alcohol consumption, and lifetime smoking) (supplemental tables 18-21).

All the analyses were conducted using the MendelianRandomization and TwoSampleMR packages, while the LD clumping (LD < 0.001) in the multivariable MR analyses between SNPs of sedentary behaviour phenotypes with those for the secondary traits was done using the ieugwasr R package (https://mrcieu.github.io/ieugwasr/) and the R programming language (version 4.1.2) (35–37). Reporting guidelines for MR studies were followed (38, 39).

## Results

### MR estimates for leisure television watching

A 1 SD (1.5 hours/day) increment in genetically-predicted duration of leisure television watching increased breast cancer risk (OR per 1 SD: 1.15, 95% confidence interval [CI]: 1.05, 1.26, P-value: 0.002) (Table 1). Similar magnitude positive effect estimates were found for all molecular subtypes of breast cancer (I^2^ = 0%, P-heterogeneity=0.98) (Table 1).

**Table1:** Mendelian Randomization estimates for sedentary behaviour and breast cancer risk

| Methods | Leisure television watching |  |  |  | Leisure computer use |  |  |  |
| --- | --- | --- | --- | --- | --- | --- | --- | --- |
|  | Estimates (OR)* | 95% CI | P-value | P-value for pleiotropy<sup>†</sup> or heterogeneity<sup>‡</sup> | Estimates (OR)* | 95% CI | P-value | P-value for pleiotropy<sup>†</sup> or heterogeneity<sup>‡</sup> |
| **breast cancer** |  |  |  |  |  |  |  |  |
| Inverse-variance weighted | 1.15 | 1.05, 1.26 | 0.002 | $1 \times 10^{-17}$ | 1.01 | 0.84, 1.23 | 0.89 | $1 \times 10^{-9}$ |
| MR-Egger | 1.48 | 0.98, 2.23 | 0.06 | 0.22 | 0.69 | 0.19, 2.48 | 0.57 | 0.55 |
| Weighted median | 1.16 | 1.05, 1.27 | 0.003 |  | 1.06 | 0.87, 1.28 | 0.57 |  |
| MR-PRESSO | 1.12 | 1.03, 1.20 | 0.008 | $3 \times 10^{-8}$ | 1.04 | 0.88, 1.23 | 0.62 | $8 \times 10^{-4}$ |
| **Luminal A breast cancer** |  |  |  |  |  |  |  |  |
| Inverse-variance weighted | 1.20 | 1.06, 1.35 | 0.002 | $6 \times 10^{-19}$ | 1.06 | 0.84, 1.34 | 0.62 | $4 \times 10^{-6}$ |
| MR-Egger | 1.55 | 0.90, 2.69 | 0.11 | 0.34 | 1.58 | 0.35, 7.10 | 0.55 | 0.60 |
| Weighted median | 1.15 | 1.01, 1.31 | 0.03 |  | 1.06 | 0.83, 1.35 | 0.66 |  |
| MR-PRESSO | 1.14 | 1.03, 1.26 | 0.01 | $3 \times 10^{-7}$ | 1.06 | 0.87, 1.31 | 0.54 | 0.003 |
| **Luminal B breast cancer** |  |  |  |  |  |  |  |  |
| Inverse-variance weighted | 1.14 | 0.94, 1.38 | 0.19 | 0.03 | 0.89 | 0.58, 1.36 | 0.58 | 0.02 |
| MR-Egger | 1.16 | 0.47, 2.89 | 0.74 | 0.96 | 1.95 | 0.12, 30.3 | 0.63 | 0.57 |
| Weighted median | 1.13 | 0.86, 1.48 | 0.40 |  | 0.97 | 0.57, 1.67 | 0.92 |  |
| MR-PRESSO |  |  |  |  | 0.82 | 0.57, 1.17 | 0.28 | 0.11 |
| **Luminal B HER2 negative breast cancer** |  |  |  |  |  |  |  |  |
| Inverse-variance weighted | 1.14 | 0.96, 1.36 | 0.13 | 0.004 | 1.03 | 0.76, 1.40 | 0.84 | 0.19 |
| MR-Egger | 1.07 | 0.48, 2.39 | 0.86 | 0.88 | 0.27 | 0.04, 2.25 | 0.23 | 0.22 |
| Weighted median | 1.30 | 1.03, 1.63 | 0.03 |  | 1.15 | 0.76, 1.75 | 0.52 |  |
| MR-PRESSO |  |  |  |  |  |  |  |  |
| **HER2 enriched breast cancer** |  |  |  |  |  |  |  |  |
| Inverse-variance weighted | 1.21 | 0.91, 1.60 | 0.19 | 0.02 | 0.67 | 0.40, 1.13 | 0.13 | 0.69 |
| MR-Egger | 1.31 | 0.35, 4.95 | 0.68 | 0.90 | 0.08 | 0.00, 2.16 | 0.13 | 0.20 |
| Weighted median | 1.25 | 0.84, 1.86 | 0.28 |  | 0.65 | 0.31, 1.35 | 0.25 |  |
| MR-PRESSO |  |  |  |  |  |  |  |  |
| **Triple negative breast cancer** |  |  |  |  |  |  |  |  |
| Inverse-variance weighted | 1.16 | 0.99, 1.35 | 0.06 | 0.10 | 0.68 | 0.50, 0.93 | 0.02 | 0.24 |
| MR-Egger | 1.54 | 0.72, 3.29 | 0.27 | 0.45 | 0.41 | 0.05, 3.35 | 0.40 | 0.63 |
| Weighted median | 1.31 | 1.04, 1.67 | 0.02 |  | 0.73 | 0.47, 1.14 | 0.16 |  |
| MR-PRESSO |  |  |  |  |  |  |  |  |

Abbreviations: CI, confidence intervals; MR: Mendelian Randomization; OR: odds ratio; MR-PRESSO: MR pleiotropy residual sum and outlier test
The estimates correspond to a standard deviation increase in duration of sedentary activity
P-value or pleiotropy based on MR-Egger intercept
P-value for heterogeneity based on Q statistic

A 1 SD increment in genetically-predicted duration of leisure television watching increased colorectal cancer risk (OR per 1 SD: 1.32, 95% CI: 1.16, 1.49, P-value: 2×10^-5^) with similar significant estimates being observed for men and women (I^2^ = 42%, P-heterogeneity=0.19) and by subsite (I^2^ = 45%, P-heterogeneity=0.17) (Table 2).

**Table2:**
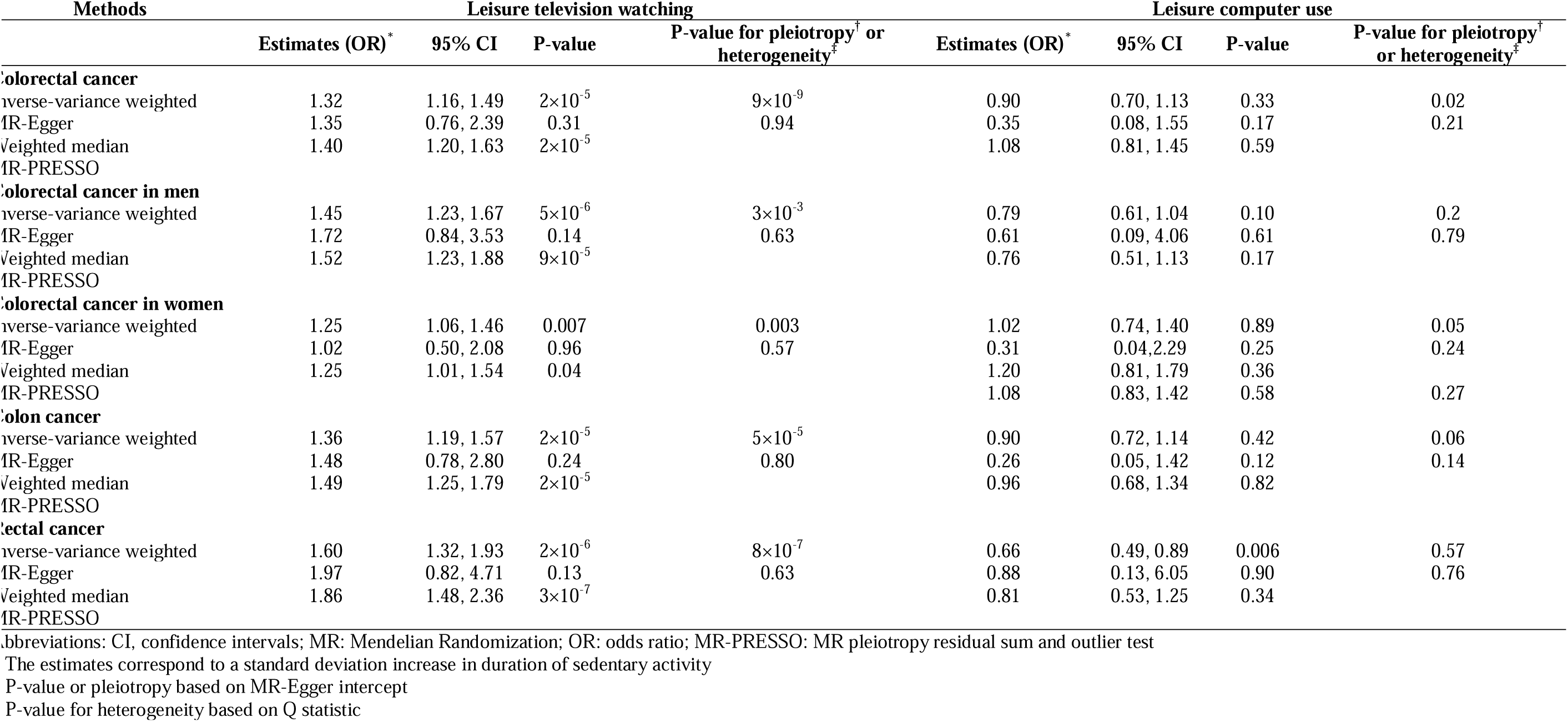
Mendelian Randomization estimates for sedentary behaviour and colorectal cancer risk

There was little evidence that a 1 SD increment in genetically-predicted duration of leisure television watching was associated with risk of overall (OR per 1 SD: 0.94, 95% CI: 0.84, 1.06, P-value:0.34) or aggressive (OR per 1 SD: 0.95, 95% CI: 0.81, 1.13, P-value:0.59) (overall vs aggressive; I^2^ = 0%, P-heterogeneity=0.92) prostate cancer (Table 3).

**Table3:** Mendelian Randomization estimates for sedentary behaviour and prostate cancer risk

| Methods | Leisure television watching |  |  |  | Leisure computer use |  |  |  |
| --- | --- | --- | --- | --- | --- | --- | --- | --- |
|  | Estimates (OR) * | 95% CI | P-value | P-value for pleiotropy<sup>†</sup> or heterogeneity<sup>‡</sup> | Estimates (OR) * | 95% CI | P-value | P-value for pleiotropy<sup>†</sup> or heterogeneity<sup>‡</sup> |
| **Prostate cancer** |  |  |  |  |  |  |  |  |
| Inverse-variance weighted | 0.94 | 0.84, 1.06 | 0.34 | $3 \times 10^{-12}$ | 1.08 | 0.89, 1.34 | 0.42 | 0.01 |
| MR-Egger | 1.19 | 0.71, 1.99 | 0.51 | 0.37 | 0.70 | 0.19, 2.56 | 0.59 | 0.5 |
| Weighted median | 0.94 | 0.83, 1.08 | 0.41 |  | 1.13 | 0.88, 1.46 | 0.33 |  |
| MR-PRESSO | 0.92 | 0.84, 1.02 | 0.13 | $1 \times 10^{-5}$ | 1.14 | 0.96, 1.35 | 0.13 | 0.09 |
| **Advanced prostate cancer** |  |  |  |  |  |  |  |  |
| Inverse-variance weighted | 0.95 | 0.81, 1.13 | 0.59 | $3 \times 10^{-4}$ | 0.91 | 0.69, 1.22 | 0.54 | 0.1 |
| MR-Egger | 1.46 | 0.68, 3.16 | 0.33 | 0.26 | 1.05 | 0.14, 8.17 | 0.96 | 0.89 |
| Weighted median | 0.82 | 0.66, 1.02 | 0.07 |  | 0.96 | 0.62, 11.48 | 0.84 |  |
| MR-PRESSO |  |  |  |  |  |  |  |  |

Abbreviations: CI, confidence intervals; MR: Mendelian Randomization; OR: odds ratio; MR-PRESSO: MR pleiotropy residual sum and outlier test
The estimates correspond to a standard deviation increase in duration of sedentary activity
P-value or pleiotropy based on MR-Egger intercept
P-value for heterogeneity based on Q statistic

Based on the Cochran’s Q values there was evidence of heterogeneity of SNP effects for most outcomes except for triple negative breast cancer (Tables 1-3). Scatter plots (with coloured lines representing the slopes of the different regression analyses) and funnel plots of the association between leisure television watching and the risk of breast, colorectal and prostate cancer risk are presented in Supplemental Figures 1-6.

**Figure 1:** Associations of leisure time television watching and computer use with breast and colorectal cancer after adjusting for the four secondary traits. The black dot corresponds to the 1-SD odds ratio and the corresponding error bar to the 95% confidence interval. Abbreviations: BMI: Body mass index; IVW: inverse variance weighting; SD: standard deviation

The multivariable MR analysis adjusting for years of education led to the attenuation of all effect estimates between genetically-predicted television watching and the risk of breast (OR per 1 SD: 1.08, 95% CI: 0.92, 1.27) and colorectal cancer (OR per 1 SD: 1.08, 95% CI: 0.90, 1.31) (Figure 1, Supplemental Table 22). Additional attenuations were observed for the models adjusting for lifetime smoking. For women, risk estimates for colorectal cancer were attenuated towards the null in all multivariable MR models adjusting for each of the four secondary traits (Figure 1, Supplemental Table 22). Finally, genetically-predicted television watching was associated with HER2 negative, HER2 positive, and triple negative breast cancer after adjusting for BMI in the multivariable MR models with effect sizes ranging from 1.32 to 1.46 per SD (Figure 1).

### MR estimates for leisure computer use

There was little evidence of any causal effect of longer duration of genetically predicted leisure computer use with overall breast, colorectal, and prostate cancer (Tables 1-3). Inverse effect-estimates were found for triple negative breast cancer (OR per 1 SD: 0.68, 95% CI: 0.50, 0.93, P-value: 0.02) and rectal cancer (OR per 1 SD: 0.66, 95% CI: 0.49, 0.89, P-value: 6×10^-3^) (Tables 1,2). Despite this, little evidence of heterogeneity was found by breast cancer subtype (I^2^ = 36%, P-heterogeneity=0.17), colorectal cancer subsite (I^2^ = 45%, P-heterogeneity=0.15), or by prostate cancer status (overall vs aggressive; I^2^ = 0%, P-heterogeneity=0.34), or sex (colorectal cancer: I^2^ = 31%, P-heterogeneity=0.23).

Based on Cochran’s Q values, heterogeneity in SNP effects was found for overall breast cancer, luminal A breast cancer, luminal B breast cancer, and colorectal cancer. Scatter plots (with coloured lines representing the slopes of the different regression analyses) and funnel plots of the association between leisure computer use and risks of breast, colorectal and prostate cancer are presented in Supplemental Figures 7-12.

In the multivariable MR analysis for triple negative breast cancer, after adjusting for years of education, alcohol, or BMI the inverse effect estimates for genetically-predicted computer use found in the univariable MR analysis were no longer statistically significant with the new attenuated effect sizes ranging from 0.73 to 1.06 per SD (Figure 1, Supplemental Table 22). Similarly, the inverse effect estimates for rectal cancer observed in the univariable analysis were attenuated after adjusting for years of education or alcohol consumption (Figure 1, Supplemental Table 22).

### Evaluation of assumptions and sensitivity analyses

The strength of the genetic instruments according to the F-statistic was ≥10 for both exposures of interest and ranged between 23 and 164 (Supplemental Tables 1-3). Little evidence of directional pleiotropy was observed based on the MR-Egger’s test (MR-Egger intercept P-values > 0.05) (Tables 1-3). The effect estimates from MR Egger regression models were generally in the same direction with those from the main analysis but with wider confidence intervals (Tables 1-3). Similarly, the weighted-median approach effect estimates were consistent in direction and magnitude to the IVW models (Tables 1-3). The MR-PRESSO analysis identified several (10 in total) outlying SNPs (Supplemental Table 23); however, no major differences were observed when these outlying genetic variants were excluded from the analyses (Tables 1-3). After examining Phenoscanner, we found that several of the genetic variants were also associated with adiposity or education-related phenotypes, such as BMI and highest qualification (Supplemental Table 24). In the multivariable MR framework, the conditional F statistics were in general above 10 (indicating little evidence of weak instrument bias) for both our exposures of interest and the adjusting factors with a few exceptions. For models including television watching and years of education, conditional F statistics for both variables were below 10. Also, adjusting for BMI or years of education resulted in low F statistics (<10) for computer use.

### MR estimates for the bidirectional MR

In post-hoc analyses, inverse bidirectional associations were observed between the genetically-predicted duration of leisure television watching and years of education. A one SD increase in genetically-predicted duration of leisure television watching reduced years of education by 0.54 SD (95% CI: -0.58 to -0.49). Similarly, a one SD increase in genetically-predicted years of education reduced duration of leisure television watching by 0.63 SD (95% CI: -0.66 to -0.59) (Figure 2, Supplemental Tables 25,26). These observations taken together with the inverse effect estimate found for years of years of education with breast and colorectal cancer (Supplemental Table 27) point to education having a complex dual confounding and mediating role in the association between television watching with breast and colorectal cancer risk. Contrary to this, positive bidirectional associations were observed for genetically-predicted duration of leisure computer use (beta_computer use→education_0.59; 95% CI: 0.48 to 0.70 and beta_education→computer use_ 0.34; 95% CI: 0.30 to 0.37).

**Figure 2:** Bidirectional associations of leisure time television watching and computer use with the four secondary traits: ΒΜΙ, years of education, smoking, and alcohol. The solid lines correspond to the effects of time television watching and computer use on the four secondary traits while the dashed lines correspond to the effects of the four secondary traits on time television watching and computer use. The black colour corresponds to statistically significant associations and the grey colour to non-significant. All the results, odds ratios and 95% confidence intervals, correspond to a 1-SD change in the levels of the variables. Abbreviations: BMI: Body mass index

Additionally, positive bidirectional associations were observed between the genetically-predicted duration of leisure television watching with BMI and smoking status while, inverse bidirectional associations were observed between the genetically-predicted duration of leisure computer use and smoking status. Finally, alcohol consumption was inversely associated with computer use (Figure 2, Supplemental Tables 25,26).

## Discussion

In this MR analysis, a high level of genetically-predicted television watching increased risks of breast and colorectal cancer. The effect estimates for television watching were robust according to most of the univariable sensitivity analyses conducted to assess the influence of pleiotropy. After multivariable MR adjustment for years of education, the positive effects were attenuated; however, our post-hoc analyses suggest that education has a complex dual confounding and mediating role in the association between television watching with these cancers and adjustment for years of education is not appropriate. We found little evidence that genetically-predicted leisure computer use was associated with breast, colorectal, and prostate cancer.

Inconsistent results have been reported in prospective cohort studies that have examined the association between sedentary behaviours and breast cancer risk. A recent meta-analysis reported a statistically significant 10% higher risk for the highest sedentary behaviour group when compared with the lowest group (8). However, a recent study in UK Biobank found little evidence of any association between hours spent watching television and the risk of breast cancer (OR per 1 hour increase: 1.01, 95% CI: 0.99, 1.03) (9). In our analysis we initially observed positive associations between hours of television watching and the risk of breast cancer. However, these positive effect estimates were attenuated towards the null in our multivariable MR models adjusting for other risk factors, particularly years of education.

Numerous observational studies have investigated the associations between sedentary behaviours and colorectal cancer risk. Results from the most recent meta-analysis of case-control and cohort studies reported a non-significant 10% risk increase for colorectal cancer for the highest sedentary behaviour group when compared with the lowest group (RR=1.10, 95% CI: 0.96–1.26) (8). Television viewing time has been the most investigated sedentary behaviour trait and positive associations have been found with colon cancer (9, 40). A recent UK Biobank analysis reported that higher levels of television watching time were associated with greater colon cancer risk (HR per 1-hour increase, 1.04, 95% CI: 1.01–1.07; P-value=0.016), but not rectal cancer (9). The same UK Biobank study found no association between leisure computer use and colorectal cancer risk (9). Results from our univariable MR analyses were generally consistent with this prior observational evidence, with positive effect estimates found for television watching, and little evidence of an association between computer use and colorectal cancer risk, except of rectal cancer. However, like our breast results, these associations attenuated towards the null in multivariable MR models adjusted for years of education and smoking (colorectal; television watching) or alcohol (rectal; computer use).

We found little evidence of any associations between sedentary behaviours and prostate cancer risk, consistent with prior observational evidence (9, 40). The null effects we found were similar for overall and aggressive prostate cancer risk.

Strong genetic correlations have been reported between each of television watching (inverse) and computer use (positive) and years of education (r^TV^= -0.79 and r^PC^= 0.53) (16). The low conditional F statistics in our multivariable models including the sedentary behaviour traits with years of education provided a further indicator of strong correlations. A recent MR study reported an inverse association between years of education and breast (OR, 0.89, 95% CI: 0.83, 0.96; P-value = 0.001) and a positive association for prostate cancer (OR, 1.10, 95% CI: 1.01, 1.21; P-value = 0.035) (41). In agreement with that, we observed inverse effect estimates for years of education in our multivariable models for breast and also for colorectal cancer. An additional MR study found that higher educational attainment levels were further inversely associated with smoking, BMI, and sedentary behaviours, and positively with vigorous physical activity levels and alcohol consumption (42). Therefore, education may be a proxy for overall lifestyle, with higher educated individuals practising healthier lifestyle behaviours and actively participating in screening programs that lower their risk of developing cancer (41). Additionally, traits like sedentary behaviours, education, smoking, alcohol consumption, and obesity are correlated and it is therefore difficult to disentangle their complex interrelationships. As an example, in our post-hoc analyses we found evidence of education having a dual confounding and mediating role in the association between television watching with breast and colorectal cancers.

The main strength of the current study is the use of large-scale summary genetic data from consortia and the UK Biobank that allowed us to investigate the role of leisure sedentary behaviours on risk of developing breast, colorectal, and prostate cancer. A limitation of our study is that leisure sedentary behaviours were derived from self-reported questionnaires that are prone to measurement error (43, 44). An alternative approach is to use genetic instruments derived from objectively measured levels of physical activity using accelerometer data from the UK Biobank. However, a current limitation is that the number of genetic instruments is small as the GWAS on accelerometer data was analysed in a subset of 90,000 participants.

Analysing two highly correlated phenotypes together, like sedentary behaviours and years of education may have introduced collinearity which leads to greater imprecision and possible bias. Furthermore, caution is needed regarding the results from the analyses for leisure computer use as the genetic instruments explained a very small proportion of the phenotypic variance resulting in a low powered analysis. Also, our analyses focused solely on leisure sedentary behaviours. The genetic correlation between television watching and objectively measured sedentary behaviour in UK Biobank was weak while, the correlation for computer use was higher (r^TV^= 0.14 and r^PC^= 0.46) (16). This can be at least partially explained from the fact that accelerometers measure total but not domain-specific sedentary time (e.g., television watching) and that has been observed in previous observational studies (3, 45). Therefore, our results cannot be generalised to overall sedentary behaviour. Finally, the results cannot be generalised to diverse populations due to the lack of ancestral diversity in UK Biobank.

In conclusion, we found that higher genetically predicted television watching time increased risks of breast and colorectal cancer in univariable models. When we adjusted for years of education in multivariable MR models, these positive effect estimates were no longer present. However, these multivariable results should be interpreted cautiously as we detected evidence of education having a dual confounding and mediating role in the associations between television watching with risks of breast and colorectal cancer. Future analyses utilising objective measures of exposure (e.g., accelerometers) and novel analytic frameworks (e.g., target trial emulation) are required to provide new insights into the possible role of sedentary behaviour in cancer development.

## Data availability

The summary statistics used in this study are outlined in the supplementary materials.

## Ethics approval

All analyses were conducted using summary-level data generated by previous studies that have described their relevant ethical approvals.

## Author contributions

NP contributed to the analysis and interpretation of data and draft of the manuscript. NK contributed to the analysis and to the data acquisition. NM contributed to the conceptualization, interpretation of data and methodology. KKT, RMM, SJL, BML contributed to interpretation of data and methodology. UP contributed to the data acquisition. NP, NK, ND, KKT, RMM, SJL, BML, MH, SSK, LL, RLM, LCS, RES, AIP, JCF, UP, SCDS, MJG, and NM reviewed, contributed to, and approved the final version of the manuscript.

## Funding

This work is supported by a WCRF grant (WCRF_2020_019). RMM is supported by a Cancer Research UK Programme Grant, the Integrative Cancer Epidemiology Programme (C18281/A29019). RMM is a member of the MRC IEU which is supported by the Medical Research Council and the University of Bristol (MC_UU_12013/1-9). RMM is supported by the National Institute for Health Research (NIHR) Bristol Biomedical Research Centre which is funded by the National Institute for Health Research and is a partnership between University Hospitals Bristol NHS Trust, Weston NHS Foundation Trust and the University of Bristol. The views expressed in this publication are those of the author(s) and not necessarily those of the NIHR or the UK Department of Health and Social Care.

BML is supported by the Victorian Cancer Agency (MCRF-18005).

SJL is supported by a WCRF grant (WCRF_2020_019).

GECCO: Genetics and Epidemiology of Colorectal Cancer Consortium: National Cancer Institute, National Institutes of Health, U.S. Department of Health and Human Services (U01 CA164930, U01 CA137088, R01 CA059045, R21 CA191312, R01201407).

Genotyping/Sequencing services were provided by the Center for Inherited Disease Research (CIDR) contract number HHSN268201700006I and HHSN268201200008I . This research was funded in part through the NIH/NCI Cancer Center Support Grant P30 CA015704.

Scientific Computing Infrastructure at Fred Hutch funded by ORIP grant S10OD028685 ASTERISK: a Hospital Clinical Research Program (PHRC-BRD09/C) from the University Hospital Center of Nantes (CHU de Nantes) and supported by the Regional Council of Pays de la Loire, the Groupement des Entreprises Françaises dans la Lutte contre le Cancer (GEFLUC), the Association Anne de Bretagne Génétique and the Ligue Régionale Contre le Cancer (LRCC).

The ATBC Study is supported by the Intramural Research Program of the U.S. National Cancer Institute, National Institutes of Health.

CLUE II funding was from the National Cancer Institute (U01 CA86308, Early Detection Research Network; P30 CA006973), National Institute on Aging (U01 AG18033), and the American Institute for Cancer Research. The content of this publication does not necessarily reflect the views or policies of the Department of Health and Human Services, nor does mention of trade names, commercial products, or organizations imply endorsement by the US government. Maryland Cancer Registry (MCR) Cancer data was provided by the Maryland Cancer Registry, Center for Cancer Prevention and Control, Maryland Department of Health, with funding from the State of Maryland and the Maryland Cigarette Restitution Fund. The collection and availability of cancer registry data is also supported by the Cooperative Agreement NU58DP006333, funded by the Centers for Disease Control and Prevention. Its contents are solely the responsibility of the authors and do not necessarily represent the official views of the Centers for Disease Control and Prevention or the Department of Health and Human Services.

ColoCare: This work was supported by the National Institutes of Health (grant numbers R01 CA189184 (Li/Ulrich), U01 CA206110 (Ulrich/Li/Siegel/Figueireido/Colditz, 2P30CA015704-40 (Gilliland), R01 CA207371 (Ulrich/Li)), the Matthias Lackas-Foundation, the German Consortium for Translational Cancer Research, and the EU TRANSCAN initiative.

The Colon Cancer Family Registry (CCFR, www.coloncfr.org) is supported in part by funding from the National Cancer Institute (NCI), National Institutes of Health (NIH) (award U01 CA167551). Support for case ascertainment was provided in part from the Surveillance, Epidemiology, and End Results (SEER) Program and the following U.S. state cancer registries: AZ, CO, MN, NC, NH; and by the Victoria Cancer Registry (Australia) and Ontario Cancer Registry (Canada). The CCFR Set-1 (Illumina 1M/1M-Duo) was supported by NIH awards U01 CA122839 and R01 CA143247 (to GC). The CCFR Set-3 (Affymetrix Axiom CORECT Set array) was supported by NIH award U19 CA148107 and R01 CA81488 (to SBG). The CCFR Set-4 (Illumina OncoArray 600K SNP array) was supported by NIH award U19 CA148107 (to SBG) and by the Center for Inherited Disease Research (CIDR), which is funded by the NIH to the Johns Hopkins University, contract number HHSN268201200008I. Additional funding for the The content of this manuscript does not necessarily reflect the views or policies of the NCI, NIH or any of the collaborating centers in the Colon Cancer Family Registry (CCFR), nor does mention of trade names, commercial products, or organizations imply endorsement by the US Government, any cancer registry, or the CCFR.

COLON: The COLON study is sponsored by Wereld Kanker Onderzoek Fonds, including funds from grant 2014/1179 as part of the World Cancer Research Fund International Regular Grant Programme, by Alpe d’Huzes and the Dutch Cancer Society (UM 2012–5653, UW 2013-5927, UW2015-7946), and by TRANSCAN (JTC2012-MetaboCCC, JTC2013-

FOCUS). The Nqplus study is sponsored by a ZonMW investment grant (98–10030); by PREVIEW, the project PREVention of diabetes through lifestyle intervention and population studies in Europe and around the World (PREVIEW) project which received funding from the European Union Seventh Framework Programme (FP7/2007–2013) under grant no. 312057; by funds from TI Food and Nutrition (cardiovascular health theme), a public–private partnership on precompetitive research in food and nutrition; and by FOODBALL, the Food Biomarker Alliance, a project from JPI Healthy Diet for a Healthy Life.

Colorectal Cancer Transdisciplinary (CORECT) Study: The CORECT Study was supported by the National Cancer Institute, National Institutes of Health (NCI/NIH), U.S. Department of Health and Human Services (grant numbers U19 CA148107, R01 CA81488, P30 CA014089, R01 CA197350,; P01 CA196569; R01 CA201407) and National Institutes of Environmental Health Sciences, National Institutes of Health (grant number T32 ES013678).

CORSA: “Österreichische Nationalbank Jubiläumsfondsprojekt” (12511) and Austrian Research Funding Agency (FFG) grant 829675.

CPS-II: The American Cancer Society funds the creation, maintenance, and updating of the Cancer Prevention Study-II (CPS-II) cohort. This study was conducted with Institutional Review Board approval.

CRCGEN: Colorectal Cancer Genetics & Genomics, Spanish study was supported by Instituto de Salud Carlos III, co-funded by FEDER funds –a way to build Europe– (grants PI14-613 and PI09-1286), Agency for Management of University and Research Grants (AGAUR) of the Catalan Government (grant 2017SGR723), and Junta de Castilla y León (grant LE22A10-2). Sample collection of this work was supported by the Xarxa de Bancs de Tumors de Catalunya sponsored by Pla Director d’Oncología de Catalunya (XBTC), Plataforma Biobancos PT13/0010/0013 and ICOBIOBANC, sponsored by the Catalan Institute of Oncology.

Czech Republic CCS: This work was supported by the Czech Science Foundation (20-03997S) and by the Grant Agency of the Ministry of Health of the Czech Republic (grants NV18/03/00199 and NU21-07-00247).

DACHS: This work was supported by the German Research Council (BR 1704/6-1, BR 1704/6-3, BR 1704/6-4, CH 117/1-1, HO 5117/2-1, HE 5998/2-1, KL 2354/3-1, RO 2270/8-1

and BR 1704/17-1), the Interdisciplinary Research Program of the National Center for Tumor Diseases (NCT), Germany, and the German Federal Ministry of Education and Research (01KH0404, 01ER0814, 01ER0815, 01ER1505A and 01ER1505B).

DALS: National Institutes of Health (R01 CA48998 to M. L. Slattery).

EDRN: This work is funded and supported by the NCI, EDRN Grant (U01CA152753). EPIC: The coordination of EPIC is financially supported by the European Commission (DGSANCO) and the International Agency for Research on Cancer. The national cohorts are supported by Danish Cancer Society (Denmark); Ligue Contre le Cancer, Institut Gustave Roussy, Mutuelle Générale de l’Education Nationale, Institut National de la Santé et de la Recherche Médicale (INSERM) (France); German Cancer Aid, German Cancer Research Center (DKFZ), Federal Ministry of Education and Research (BMBF), Deutsche Krebshilfe, Deutsches Krebsforschungszentrum and Federal Ministry of Education and Research (Germany); the Hellenic Health Foundation (Greece); Associazione Italiana per la Ricerca sul Cancro-AIRCItaly and National Research Council (Italy); Dutch Ministry of Public Health, Welfare and Sports (VWS), Netherlands Cancer Registry (NKR), LK Research Funds, Dutch Prevention Funds, Dutch ZON (Zorg Onderzoek Nederland), World Cancer Research Fund (WCRF), Statistics Netherlands (The Netherlands); ERC-2009-AdG 232997 and Nordforsk, Nordic Centre of Excellence programme on Food, Nutrition and Health (Norway); Health Research Fund (FIS), PI13/00061 to Granada, PI13/01162 to EPIC-Murcia, Regional Governments of Andalucía, Asturias, Basque Country, Murcia and Navarra, ISCIII RETIC (RD06/0020) (Spain); Swedish Cancer Society, Swedish Research Council and County Councils of Skåne and Västerbotten (Sweden); Cancer Research UK (14136 to EPIC-Norfolk; C570/A16491 and C8221/A19170 to EPIC-Oxford), Medical Research Council (1000143 to EPIC-Norfolk, MR/M012190/1 to EPICOxford) (United Kingdom).

The EPIC-Norfolk study (https://doi.org/10.22025/2019.10.105.00004) has received funding from the Medical Research Council (MR/N003284/1 and MC-UU_12015/1) and Cancer Research UK (C864/A14136). The genetics work in the EPIC-Norfolk study was funded by the Medical Research Council (MC_PC_13048). Metabolite measurements in the EPIC-Norfolk study were supported by the MRC Cambridge Initiative in Metabolic Science (MR/L00002/1) and the Innovative Medicines Initiative Joint Undertaking under EMIF grant agreement no. 115372.

EPICOLON: This work was supported by grants from Fondo de Investigación Sanitaria/FEDER (PI08/0024, PI08/1276, PS09/02368, PI11/00219, PI11/00681, PI14/00173,

PI14/00230, PI17/00509, 17/00878, PI20/00113, PI20/00226, Acción Transversal de Cáncer), Xunta de Galicia (PGIDIT07PXIB9101209PR), Ministerio de Economia y Competitividad (SAF07-64873, SAF 2010-19273, SAF2014-54453R), Fundación Científica de la Asociación Española contra el Cáncer (GCB13131592CAST), Beca Grupo de Trabajo “Oncología” AEG (Asociación Española de Gastroenterología), Fundación Privada Olga Torres, FP7 CHIBCHA Consortium, Agència de Gestió d’Ajuts Universitaris i de Recerca (AGAUR, Generalitat de Catalunya, 2014SGR135, 2014SGR255, 2017SGR21, 2017SGR653), Catalan Tumour Bank Network (Pla Director d’Oncologia, Generalitat de Catalunya), PERIS (SLT002/16/00398, Generalitat de Catalunya), CERCA Programme (Generalitat de Catalunya) and COST Actions BM1206 and CA17118. CIBERehd is funded by the Instituto de Salud Carlos III. ESTHER/VERDI. This work was supported by grants from the Baden-Württemberg Ministry of Science, Research and Arts and the German Cancer Aid.

Harvard cohorts (HPFS, NHS, PHS): HPFS is supported by the National Institutes of Health (P01 CA055075, UM1 CA167552, U01 CA167552, R01 CA137178, R01 CA151993, R35

CA197735, K07 CA190673, and P50 CA127003), NHS by the National Institutes of Health (R01 CA137178, P01 CA087969, UM1 CA186107, R01 CA151993, R35 CA197735,

K07CA190673, and P50 CA127003) and PHS by the National Institutes of Health (R01 CA042182).

Hawaii Adenoma Study: NCI grants R01 CA72520.

HCES-CRC: the Hwasun Cancer Epidemiology Study–Colon and Rectum Cancer (HCES-CRC; grants from Chonnam National University Hwasun Hospital, HCRI21019).

Kentucky: This work was supported by the following grant support: Clinical Investigator Award from Damon Runyon Cancer Research Foundation (CI-8); NCI R01CA136726.

LCCS: The Leeds Colorectal Cancer Study was funded by the Food Standards Agency and Cancer Research UK Programme Award (C588/A19167).

Melbourne Collaborative Cohort Study (MCCS) cohort recruitment was funded by VicHealth and Cancer Council Victoria. The MCCS was further augmented by Australian National Health and Medical Research Council grants 209057, 396414 and 1074383 and by infrastructure provided by Cancer Council Victoria. Cases and their vital status were ascertained through the Victorian Cancer Registry and the Australian Institute of Health and Welfare, including the National Death Index and the Australian Cancer Database.

Multiethnic Cohort (MEC) Study: National Institutes of Health (R37 CA54281, P01 CA033619, R01 CA063464 and U01 CA164973).

MECC: This work was supported by the National Institutes of Health, U.S. Department of Health and Human Services (R01 CA81488 to SBG and GR).

MSKCC: The work at Sloan Kettering in New York was supported by the Robert and Kate Niehaus Center for Inherited Cancer Genomics and the Romeo Milio Foundation. Moffitt: This work was supported by funding from the National Institutes of Health (grant numbers R01 CA189184, P30 CA076292), Florida Department of Health Bankhead-Coley Grant 09BN-13, and the University of South Florida Oehler Foundation. Moffitt contributions were supported in part by the Total Cancer Care Initiative, Collaborative Data Services Core, and Tissue Core at the H. Lee Moffitt Cancer Center & Research Institute, a National Cancer Institute-designated Comprehensive Cancer Center (grant number P30 CA076292).

NCCCS I & II: We acknowledge funding support for this project from the National Institutes of Health, R01 CA66635 and P30 DK034987.

NFCCR: This work was supported by an Interdisciplinary Health Research Team award from the Canadian Institutes of Health Research (CRT 43821); the National Institutes of Health,

U.S. Department of Health and Human Serivces (U01 CA74783); and National Cancer

Institute of Canada grants (18223 and 18226). The authors wish to acknowledge the contribution of Alexandre Belisle and the genotyping team of the McGill University and Génome Québec Innovation Centre, Montréal, Canada, for genotyping the Sequenom panel in the NFCCR samples. Funding was provided to Michael O. Woods by the Canadian Cancer Society Research Institute.

NSHDS: Swedish Research Council; Swedish Cancer Society; Cutting-Edge Research Grant and other grants from Region Västerbotten; Knut and Alice Wallenberg Foundation; Lion’s Cancer Research Foundation at Umeå University; the Cancer Research Foundation in Northern Sweden; and the Faculty of Medicine, Umeå University, Umeå, Sweden.

OSUMC: OCCPI funding was provided by Pelotonia and HNPCC funding was provided by the NCI (CA16058 and CA67941).

PLCO: Intramural Research Program of the Division of Cancer Epidemiology and Genetics and supported by contracts from the Division of Cancer Prevention, National Cancer Institute, NIH, DHHS. Funding was provided by National Institutes of Health (NIH), Genes, Environment and Health Initiative (GEI) Z01 CP 010200, NIH U01 HG004446, and NIH GEI U01 HG 004438.

SEARCH: The University of Cambridge has received salary support in respect of PDPP from the NHS in the East of England through the Clinical Academic Reserve. Cancer Research UK (C490/A16561); the UK National Institute for Health Research Biomedical Research Centres at the University of Cambridge.

SELECT: Research reported in this publication was supported in part by the National Cancer Institute of the National Institutes of Health under Award Numbers U10 CA37429 (CD Blanke), and UM1 CA182883 (CM Tangen/IM Thompson). The content is solely the responsibility of the authors and does not necessarily represent the official views of the National Institutes of Health.

SMS and REACH: This work was supported by the National Cancer Institute (grant P01 CA074184 to J.D.P. and P.A.N., grants R01 CA097325, R03 CA153323, and K05 CA152715 to P.A.N., and the National Center for Advancing Translational Sciences at the National Institutes of Health (grant KL2 TR000421 to A.N.B.-H.)

The Swedish Low-risk Colorectal Cancer Study: The study was supported by grants from the Swedish research council; K2015-55X-22674-01-4, K2008-55X-20157-03-3, K2006-72X-20157-01-2 and the Stockholm County Council (ALF project).

Swedish Mammography Cohort and Cohort of Swedish Men: This work is supported by the Swedish Research Council /Infrastructure grant, the Swedish Cancer Foundation, and the Karolinska Institutés Distinguished Professor Award to Alicja Wolk.

UK Biobank: This research has been conducted using the UK Biobank Resource under Application Number 8614

VITAL: National Institutes of Health (K05 CA154337).

WHI: The WHI program is funded by the National Heart, Lung, and Blood Institute, National Institutes of Health, U.S. Department of Health and Human Services through contracts HHSN268201100046C, HHSN268201100001C, HHSN268201100002C, HHSN268201100003C, HHSN268201100004C, and HHSN271201100004C.

## Data Availability

The summary statistics used in this study are outlined in the supplementary materials

## Abbreviations

BMI: body mass index
IVW: inverse-variance weighted
GWAS: genome-wide association study
LD: linkage disequilibrium
MR: Mendelian randomization
PSA: prostate-specific antigen
OR: odds ratio
RCT: randomised control trial
SNP: single nucleotide polymorphism
SD: Standard deviation

## Acknowledgements

The breast cancer genome-wide association analyses for BCAC and CIMBA were supported by Cancer Research UK (PPRPGM-Nov20\100002, C1287/A10118, C1287/A16563, C1287/A10710, C12292/A20861, C12292/A11174, C1281/A12014, C5047/A8384, C5047/A15007, C5047/A10692, C8197/A16565) and the Gray Foundation, The National Institutes of Health (CA128978, X01HG007492-the DRIVE consortium), the PERSPECTIVE project supported by the Government of Canada through Genome Canada and the Canadian Institutes of Health Research (grant GPH-129344) and the Ministère de l’Économie, Science et Innovation du Québec through Genome Québec and the PSRSIIRI-701 grant, the Quebec Breast Cancer Foundation, the European Community’s Seventh Framework Programme under grant agreement n° 223175 (HEALTH-F2-2009-223175) (COGS), the European Union’s Horizon 2020 Research and Innovation Programme (634935 and 633784), the Post-Cancer GWAS initiative (U19 CA148537, CA148065 and CA148112 - the GAME-ON initiative), the Department of Defence (W81XWH-10-1-0341), the Canadian Institutes of Health Research (CIHR) for the CIHR Team in Familial Risks of Breast Cancer (CRN-87521), the Komen Foundation for the Cure, the Breast Cancer Research Foundation and the Ovarian Cancer Research Fund. All studies and funders are listed in Zhang H et al (Nat Genet, 2020).

ASTERISK: We are very grateful to Dr. Bruno Buecher without whom this project would not have existed. We also thank all those who agreed to participate in this study, including the patients and the healthy control persons, as well as all the physicians, technicians and students.

CCFR: The Colon CFR graciously thanks the generous contributions of their study participants, dedication of study staff, and the financial support from the U.S. National Cancer Institute, without which this important registry would not exist. The authors would like to thank the study participants and staff of the Seattle Colon Cancer Family Registry and the Hormones and Colon Cancer study (CORE Studies).

CLUE II: We thank the participants of Clue II and appreciate the continued efforts of the staff at the Johns Hopkins George W. Comstock Center for Public Health Research and Prevention in the conduct of the Clue II Cohort Study.

COLON and NQplus: the authors would like to thank the COLON and NQplus investigators at Wageningen University & Research and the involved clinicians in the participating hospitals.

CORSA: We kindly thank all those who contributed to the screening project Burgenland against CRC. Furthermore, we are grateful to Doris Mejri and Monika Hunjadi for laboratory assistance.

CPS-II: The authors thank the CPS-II participants and Study Management Group for their invaluable contributions to this research. The authors would also like to acknowledge the contribution to this study from central cancer registries supported through the Centers for Disease Control and Prevention National Program of Cancer Registries, and cancer registries supported by the National Cancer Institute Surveillance Epidemiology and End Results program.

Czech Republic CCS: We are thankful to all clinicians in major hospitals in the Czech Republic, without whom the study would not be practicable. We are also sincerely grateful to all patients participating in this study.

DACHS: We thank all participants and cooperating clinicians, and Ute Handte-Daub, Utz Benscheid, Muhabbet Celik and Ursula Eilber for excellent technical assistance.

EDRN: We acknowledge all the following contributors to the development of the resource: University of Pittsburgh School of Medicine, Division of Gastroenterology, Hepatology and Nutrition: Lynda Dzubinski; University of Pittsburgh School of Medicine, Department of Pathology: Pittsburgh Biospecimen Core; and University of Pittsburgh School of Medicine, Department of Biomedical Informatics.

EPIC: Where authors are identified as personnel of the International Agency for Research on Cancer/World Health Organization, the authors alone are responsible for the views expressed in this article and they do not necessarily represent the decisions, policy or views of the International Agency for Research on Cancer/World Health Organization.

The EPIC-Norfolk study: we are grateful to all the participants who have been part of the project and to the many members of the study teams at the University of Cambridge who have enabled this research.

EPICOLON: We are sincerely grateful to all patients participating in this study who were recruited as part of the EPICOLON project. We acknowledge the Spanish National DNA Bank, Biobank of Hospital Clínic–IDIBAPS and Biobanco Vasco for the availability of the samples. The work was carried out (in part) at the Esther Koplowitz Centre, Barcelona.

Harvard cohorts (HPFS, NHS, PHS): The study protocol was approved by the institutional review boards of the Brigham and Women’s Hospital and Harvard T.H. Chan School of Public Health, and those of participating registries as required. We acknowledge Channing Division of Network Medicine, Department of Medicine, Brigham and Women’s Hospital as home of the NHS We would like to thank the participants and staff of the HPFS, NHS and PHS for their valuable contributions as well as the following state cancer registries for their help: AL, AZ, AR, CA, CO, CT, DE, FL, GA, ID, IL, IN, IA, KY, LA, ME, MD, MA, MI, NE, NH, NJ, NY, NC, ND, OH, OK, OR, PA, RI, SC, TN, TX, VA, WA, WY. The authors assume full responsibility for analyses and interpretation of these data.

Interval: A complete list of the investigators and contributors to the INTERVAL trial is provided in reference (32). The academic coordinating centre would like to thank blood donor centre staff and blood donors for participating in the INTERVAL trial.

Kentucky: We would like to acknowledge the staff at the Kentucky Cancer Registry.

LCCS: We acknowledge the contributions of Jennifer Barrett, Robin Waxman, Gillian Smith and Emma Northwood in conducting this study.

NCCCS I & II: We would like to thank the study participants, and the NC Colorectal Cancer Study staff.

NSHDS investigators thank the Biobank Research Unit at Umeå University, the Västerbotten Intervention Programme, the Northern Sweden MONICA study and Region Västerbotten for providing data and samples and acknowledge the contribution from Biobank Sweden, supported by the Swedish Research Council (VR 2017-00650).

PLCO: The authors thank the PLCO Cancer Screening Trial screening center investigators and the staff from Information Management Services Inc and Westat Inc. Most importantly, we thank the study participants for their contributions that made this study possible.

SEARCH: We thank the SEARCH team.

SELECT: We thank the research and clinical staff at the sites that participated on SELECT study, without whom the trial would not have been successful. We are also grateful to the 35,533 dedicated men who participated in SELECT.

UK Biobank: We would like to thank the participants and researchers UK Biobank for their participation and acquisition of data.

WHI: The authors thank the WHI investigators and staff for their dedication, and the study participants for making the program possible. A full listing of WHI investigators can be found at: http://www.whi.org/researchers/Documents%20%20Write%20a%20Paper/WHI%20Investigator%20Short%20List.pdf

