## supplemental tables and figures for "Leisure time sedentary behaviour and risks of breast, colorectal, and prostate cancer: A Mendelian randomization analysis" (from another version of this preprint: version 1, 2023-03-03)

Supplemental Table 1: Summary information on leisure television watching SNPs used as genetic instruments used for the Mendelian randomisation analyses

Supplemental Table 2: Summary information on computer use SNPs used as genetic instruments used for the Mendelian randomisation analyses

Supplemental Table 3: Summary information on breast cancer risk for the leisure television watching SNPs used for the Mendelian randomisation analyses

Supplemental Table 4: Summary information on breast cancer risk for the computer use SNPs used for the Mendelian randomisation analyses

Supplemental Table 5: Summary information on colorectal cancer risk for the leisure television watching SNPs used for the Mendelian randomisation analyses

Supplemental Table 6: Summary information on colorectal cancer risk for the leisure computer use SNPs used for the Mendelian randomisation analyses

Supplemental Table 7: Summary information on prostate cancer risk for the leisure television watching SNPs used for the Mendelian randomisation analyses

Supplemental Table 8: Summary information on prostate cancer risk for the leisure computer use SNPs used for the Mendelian randomisation analyses

Supplemental Table 9: Sample size and power calculations in Mendelian randomization study of sedentary behaviour and cancer risk

Supplemental Table 22: Multivariable Mendelian randomization estimates between leisure television watching, computer use and cancer risk

Supplemental Table 23: Outlying SNPs identified by MR-PRESSO

Supplemental Table 24: Evidence of association (p<5*10-8) with secondary traits of the SNPs (or their proxies) used as genetic instruments for Mendelian randomization analyses of sedentary behaviours and risk of breast and colorectal cancer

Supplemental Table 25: Mendelian Randomization estimates for the effects of sedentary behaviour on the secondary traits

Supplemental Table 26: Mendelian Randomization estimates for the effects of the secondary traits on sedentary behaviour

Supplemental Table 27: Multivariable Mendelian randomization estimates for years of education and cancer risk

**Supplemental figures**

Supplemental Figure 1: Scatter plots showing the correlation of genetic associations of leisure television watching with genetic associations with breast cancer. Coloured lines represent the slopes of the different regression analyses.

Supplemental Figure 2: Scatter plots showing the correlation of genetic associations of leisure television watching with genetic associations with colorectal cancer. Coloured lines represent the slopes of the different regression analyses.

Supplemental Figure 3: Scatter plots showing the correlation of genetic associations of leisure television watching with genetic associations with prostate cancer. Coloured lines represent the slopes of the different regression analyses.

Supplemental Figure 4: Funnel plots of risk estimates of leisure television watching and breast cancer against instrumental strength

Supplemental Figure 5: Funnel plots of risk estimates of leisure television watching and colorectal cancer against instrumental strength

Supplemental Figure 6: Funnel plots of risk estimates of leisure television watching and prostate cancer against instrumental strength

Supplemental Figure 7: Scatter plots showing the correlation of genetic associations of leisure computer use with genetic associations with breast cancer. Coloured lines represent the slopes of the different regression analyses.

Supplemental Figure 8: Scatter plots showing the correlation of genetic associations of leisure computer use with genetic associations with colorectal cancer. Coloured lines represent the slopes of the different regression analyses.

Supplemental Figure 9: Scatter plots showing the correlation of genetic associations of leisure computer use with genetic associations with prostate cancer. Coloured lines represent the slopes of the different regression analyses

Supplemental Figure 10: Funnel plots of risk estimates of leisure computer use and breast cancer against instrumental strength

Supplemental Figure 11: Funnel plots of risk estimates of leisure computer use and colorectal cancer against instrumental strength

Supplemental Figure 12: Funnel plots of risk estimates of leisure computer use and prostate cancer against instrumental strength

| **Supplemental Table 1: Summary information on leisure television watching SNPs used as genetic instruments used for the Mendelian randomisation analyses** | | | | | | | | | | |
| --- | --- | --- | --- | --- | --- | --- | --- | --- | --- | --- |
| SNP | CHR | Position^a^ | EA | OA | EAF | beta | se | Pvalue | R^2^ | F-statistic |
| rs11810109 | 1 | 44186812 | A | T | 0.701 | 0.016 | 0.002 | 5.40E-12 | 1.31E-04 | 54.8 |
| rs112632642 | 1 | 51115634 | A | G | 0.065 | 0.024 | 0.004 | 3.60E-08 | 6.78E-05 | 28.3 |
| rs6670915 | 1 | 61109968 | C | T | 0.533 | 0.014 | 0.002 | 2.40E-10 | 9.41E-05 | 39.3 |
| rs12725114 | 1 | 62432641 | G | A | 0.802 | 0.015 | 0.003 | 1.60E-08 | 7.79E-05 | 32.5 |
| rs984409 | 1 | 67020440 | A | G | 0.637 | 0.015 | 0.002 | 4.10E-11 | 1.06E-04 | 44.2 |
| rs10890123 | 1 | 74802123 | C | T | 0.770 | 0.014 | 0.003 | 3.30E-08 | 7.86E-05 | 32.8 |
| rs71658797 | 1 | 77967507 | A | T | 0.123 | 0.020 | 0.003 | 6.30E-10 | 9.05E-05 | 37.8 |
| rs114328297 | 1 | 91190854 | T | G | 0.778 | 0.014 | 0.003 | 4.40E-08 | 8.17E-05 | 34.1 |
| rs631130 | 1 | 96043010 | A | T | 0.631 | 0.014 | 0.002 | 5.00E-09 | 9.42E-05 | 39.4 |
| rs148648822 | 1 | 97815048 | T | G | 0.157 | 0.018 | 0.003 | 4.20E-09 | 8.29E-05 | 34.6 |
| rs17379561 | 1 | 98340139 | T | A | 0.144 | 0.025 | 0.003 | 1.10E-16 | 1.65E-04 | 68.9 |
| rs55909997 | 1 | 107581039 | A | G | 0.350 | 0.014 | 0.002 | 2.40E-09 | 8.31E-05 | 34.7 |
| rs1278847 | 1 | 110037483 | C | A | 0.694 | 0.016 | 0.002 | 1.20E-11 | 1.14E-04 | 47.4 |
| rs141425667 | 1 | 114930465 | A | G | 0.065 | 0.024 | 0.004 | 4.00E-08 | 7.94E-05 | 33.1 |
| rs4845364 | 1 | 154141908 | G | A | 0.505 | 0.015 | 0.002 | 1.30E-12 | 1.29E-04 | 53.9 |
| rs6673341 | 1 | 184655649 | G | T | 0.535 | 0.015 | 0.002 | 2.20E-11 | 1.13E-04 | 47.3 |
| rs72725224 | 1 | 188115858 | C | A | 0.020 | 0.046 | 0.008 | 3.70E-09 | 7.37E-05 | 30.8 |
| rs10737620 | 1 | 193057971 | T | A | 0.274 | 0.014 | 0.002 | 2.60E-09 | 9.94E-05 | 41.5 |
| rs6721975 | 2 | 5832667 | C | T | 0.768 | 0.017 | 0.003 | 1.70E-10 | 1.07E-04 | 44.7 |
| rs11896615 | 2 | 17407744 | G | A | 0.852 | 0.017 | 0.003 | 3.10E-08 | 8.10E-05 | 33.8 |
| rs72781699 | 2 | 24277709 | A | G | 0.203 | 0.019 | 0.003 | 3.00E-12 | 1.42E-04 | 59.5 |
| rs12105701 | 2 | 41737200 | T | C | 0.604 | 0.013 | 0.002 | 5.40E-09 | 7.30E-05 | 30.5 |
| rs7564130 | 2 | 50606642 | C | T | 0.359 | 0.015 | 0.002 | 2.80E-11 | 1.12E-04 | 46.6 |
| rs11125515 | 2 | 53886799 | A | C | 0.271 | 0.014 | 0.002 | 2.20E-08 | 8.16E-05 | 34.1 |
| rs12475168 | 2 | 58992704 | T | G | 0.131 | 0.018 | 0.003 | 1.70E-08 | 6.98E-05 | 29.1 |
| rs183566425 | 2 | 59325356 | A | T | 0.982 | 0.046 | 0.008 | 1.90E-08 | 9.09E-05 | 38.0 |
| rs10189857 | 2 | 60713235 | G | A | 0.432 | 0.020 | 0.002 | 6.20E-21 | 2.28E-04 | 95.3 |
| rs62641636 | 2 | 68425427 | A | G | 0.692 | 0.014 | 0.002 | 7.00E-10 | 1.02E-04 | 42.6 |
| rs59210467 | 2 | 97097291 | T | C | 0.307 | 0.013 | 0.002 | 1.00E-08 | 7.86E-05 | 32.8 |
| rs11689199 | 2 | 100820421 | A | G | 0.598 | 0.018 | 0.002 | 5.50E-17 | 1.81E-04 | 75.6 |
| rs2916251 | 2 | 104447052 | T | G | 0.510 | 0.017 | 0.002 | 1.50E-13 | 1.35E-04 | 56.5 |
| rs1451533 | 2 | 105466005 | A | G | 0.275 | 0.016 | 0.002 | 1.10E-10 | 1.04E-04 | 43.3 |
| rs2971640 | 2 | 107624618 | A | G | 0.495 | 0.013 | 0.002 | 4.20E-09 | 9.05E-05 | 37.8 |
| rs12476388 | 2 | 116330983 | C | T | 0.709 | 0.013 | 0.002 | 2.10E-08 | 9.10E-05 | 38.0 |
| rs374722 | 2 | 147839830 | G | A | 0.150 | 0.024 | 0.003 | 5.50E-16 | 1.79E-04 | 74.7 |
| rs3754970 | 2 | 162091836 | C | T | 0.504 | 0.015 | 0.002 | 4.80E-12 | 1.46E-04 | 61.1 |
| rs1913808 | 2 | 162804678 | G | A | 0.713 | 0.015 | 0.002 | 3.90E-10 | 8.72E-05 | 36.4 |
| rs2060199 | 2 | 166196681 | T | A | 0.446 | 0.012 | 0.002 | 2.40E-08 | 7.46E-05 | 31.2 |
| rs263771 | 2 | 185921692 | A | C | 0.233 | 0.017 | 0.003 | 3.10E-11 | 1.24E-04 | 51.7 |
| rs4577309 | 2 | 191288833 | A | G | 0.469 | 0.016 | 0.002 | 1.60E-13 | 1.24E-04 | 51.7 |
| rs34224261 | 2 | 193855452 | C | G | 0.531 | 0.018 | 0.002 | 4.60E-16 | 1.60E-04 | 66.8 |
| rs4675246 | 2 | 202864487 | T | G | 0.203 | 0.015 | 0.003 | 2.90E-08 | 7.37E-05 | 30.8 |
| rs13029509 | 2 | 215374209 | A | G | 0.480 | 0.018 | 0.002 | 2.00E-17 | 1.83E-04 | 76.3 |
| rs10932837 | 2 | 221044785 | T | C | 0.506 | 0.013 | 0.002 | 1.30E-09 | 9.41E-05 | 39.3 |
| rs4973576 | 2 | 233806771 | A | C | 0.702 | 0.015 | 0.002 | 1.00E-09 | 8.93E-05 | 37.3 |
| rs138256022 | 3 | 11603090 | T | C | 0.041 | 0.031 | 0.006 | 2.90E-08 | 7.63E-05 | 31.9 |
| rs9834970 | 3 | 36856030 | T | C | 0.501 | 0.013 | 0.002 | 3.30E-09 | 8.92E-05 | 37.3 |
| rs1352890 | 3 | 49652156 | T | C | 0.525 | 0.012 | 0.002 | 1.30E-08 | 8.87E-05 | 37.1 |
| rs3796386 | 3 | 49899795 | A | G | 0.428 | 0.026 | 0.002 | 3.20E-33 | 3.93E-04 | 164.3 |
| rs182355396 | 3 | 50920457 | A | C | 0.010 | 0.080 | 0.013 | 2.10E-10 | 0.00012 | 51.6 |
| rs11917871 | 3 | 54233476 | C | T | 0.575 | 0.017 | 0.002 | 4.50E-14 | 1.20E-04 | 50.3 |
| rs11130793 | 3 | 60884659 | C | T | 0.602 | 0.013 | 0.002 | 5.00E-09 | 9.51E-05 | 39.7 |
| rs11714337 | 3 | 71582521 | G | A | 0.569 | 0.014 | 0.002 | 4.70E-11 | 1.27E-04 | 53.1 |
| rs12496084 | 3 | 75174777 | G | A | 0.247 | 0.014 | 0.003 | 4.60E-08 | 7.10E-05 | 29.7 |
| rs6797840 | 3 | 85656569 | C | A | 0.544 | 0.016 | 0.002 | 1.70E-13 | 1.35E-04 | 56.4 |
| rs56398417 | 3 | 88024986 | C | T | 0.687 | 0.014 | 0.002 | 5.90E-10 | 1.04E-04 | 43.6 |
| rs2034768 | 3 | 93986371 | A | G | 0.487 | 0.015 | 0.002 | 8.60E-12 | 1.20E-04 | 50.0 |
| rs12632778 | 3 | 106452359 | G | T | 0.046 | 0.028 | 0.005 | 3.90E-08 | 7.06E-05 | 29.5 |
| rs9867121 | 3 | 114631548 | C | A | 0.818 | 0.019 | 0.003 | 3.90E-12 | 1.15E-04 | 47.9 |
| rs12491503 | 3 | 165706855 | A | G | 0.330 | 0.014 | 0.002 | 5.50E-10 | 9.14E-05 | 38.2 |
| rs114600294 | 3 | 181419367 | C | G | 0.211 | 0.016 | 0.003 | 7.90E-10 | 9.69E-05 | 40.5 |
| rs66852340 | 4 | 3311070 | T | C | 0.224 | 0.018 | 0.003 | 7.90E-12 | 1.24E-04 | 51.8 |
| rs7700107 | 4 | 17880416 | C | A | 0.137 | 0.021 | 0.003 | 1.90E-11 | 1.15E-04 | 47.9 |
| rs34811474 | 4 | 25408838 | G | A | 0.768 | 0.015 | 0.003 | 2.30E-09 | 9.89E-05 | 41.3 |
| rs77215114 | 4 | 45187658 | A | T | 0.930 | 0.024 | 0.004 | 1.50E-08 | 9.42E-05 | 39.3 |
| rs28971796 | 4 | 49151982 | A | G | 0.378 | 0.015 | 0.002 | 2.10E-09 | 8.60E-05 | 35.9 |
| rs6850494 | 4 | 82291771 | C | A | 0.384 | 0.014 | 0.002 | 1.10E-10 | 1.08E-04 | 45.0 |
| rs13107325 | 4 | 103188709 | T | C | 0.074 | 0.029 | 0.004 | 1.50E-12 | 1.36E-04 | 56.7 |
| rs62332760 | 4 | 106135316 | T | G | 0.387 | 0.014 | 0.002 | 8.00E-11 | 1.02E-04 | 42.8 |
| rs7693703 | 4 | 118357405 | G | A | 0.910 | 0.023 | 0.004 | 2.70E-09 | 8.32E-05 | 34.8 |
| rs4334769 | 4 | 130245233 | G | T | 0.474 | 0.012 | 0.002 | 2.10E-08 | 9.13E-05 | 38.1 |
| rs6825241 | 4 | 152543783 | A | C | 0.464 | 0.017 | 0.002 | 4.90E-15 | 1.52E-04 | 63.4 |
| rs7693082 | 4 | 159857819 | G | C | 0.300 | 0.015 | 0.002 | 1.70E-10 | 1.10E-04 | 46.1 |
| rs10940659 | 5 | 59360341 | G | A | 0.469 | 0.013 | 0.002 | 1.50E-08 | 9.00E-05 | 37.6 |
| rs2409838 | 5 | 60204410 | A | T | 0.555 | 0.017 | 0.002 | 3.40E-15 | 1.60E-04 | 67.0 |
| rs262890 | 5 | 62930015 | G | A | 0.301 | 0.019 | 0.002 | 3.20E-15 | 1.54E-04 | 64.4 |
| rs7716447 | 5 | 88800331 | G | A | 0.361 | 0.013 | 0.002 | 3.60E-09 | 9.29E-05 | 38.8 |
| rs1031423 | 5 | 93276883 | C | T | 0.785 | 0.019 | 0.003 | 1.80E-12 | 1.22E-04 | 50.9 |
| rs57585211 | 5 | 107197725 | G | T | 0.173 | 0.017 | 0.003 | 4.30E-09 | 8.21E-05 | 34.3 |
| rs9326862 | 5 | 112054547 | A | T | 0.468 | 0.012 | 0.002 | 2.60E-08 | 5.59E-05 | 23.3 |
| rs10041724 | 5 | 124273520 | T | C | 0.808 | 0.018 | 0.003 | 3.90E-11 | 1.18E-04 | 49.4 |
| rs10057545 | 5 | 138431008 | A | G | 0.445 | 0.014 | 0.002 | 2.00E-10 | 8.89E-05 | 37.1 |
| rs62379379 | 5 | 141082015 | T | G | 0.071 | 0.026 | 0.004 | 7.80E-10 | 8.77E-05 | 36.6 |
| rs10054327 | 5 | 147871137 | G | A | 0.576 | 0.017 | 0.002 | 3.40E-15 | 1.57E-04 | 65.5 |
| rs42210 | 5 | 166408788 | C | G | 0.711 | 0.014 | 0.002 | 7.30E-09 | 8.67E-05 | 36.2 |
| rs2336895 | 5 | 166990695 | G | A | 0.360 | 0.012 | 0.002 | 4.60E-08 | 6.39E-05 | 26.7 |
| rs72828890 | 5 | 167806836 | C | T | 0.869 | 0.019 | 0.003 | 4.90E-09 | 9.51E-05 | 39.7 |
| rs255292 | 5 | 172580866 | A | C | 0.598 | 0.012 | 0.002 | 1.70E-08 | 7.47E-05 | 31.2 |
| rs72834698 | 6 | 26176517 | G | A | 0.858 | 0.023 | 0.003 | 2.70E-13 | 1.62E-04 | 67.8 |
| rs3135044 | 6 | 31592820 | T | C | 0.360 | 0.015 | 0.002 | 8.70E-11 | 1.02E-04 | 42.8 |
| rs576432906 | 6 | 33443967 | C | A | 0.289 | 0.014 | 0.002 | 9.60E-09 | 0.00009 | 37.1 |
| rs10947452 | 6 | 33803752 | T | C | 0.349 | 0.014 | 0.002 | 6.40E-10 | 9.46E-05 | 39.5 |
| rs9471333 | 6 | 40362023 | C | T | 0.450 | 0.013 | 0.002 | 1.50E-09 | 1.05E-04 | 43.8 |
| rs4523073 | 6 | 67550288 | G | A | 0.409 | 0.014 | 0.002 | 5.00E-10 | 9.68E-05 | 40.4 |
| rs77589760 | 6 | 89827716 | C | G | 0.961 | 0.037 | 0.006 | 4.10E-10 | 8.31E-05 | 34.7 |
| rs6905544 | 6 | 98411631 | G | A | 0.601 | 0.019 | 0.002 | 8.50E-18 | 2.22E-04 | 92.8 |
| rs17789218 | 6 | 100600097 | T | C | 0.756 | 0.019 | 0.003 | 1.40E-13 | 1.36E-04 | 57.0 |
| rs78394231 | 6 | 107649123 | C | T | 0.098 | 0.021 | 0.004 | 9.70E-09 | 1.03E-04 | 42.8 |
| rs2184364 | 6 | 143183596 | A | G | 0.782 | 0.016 | 0.003 | 3.00E-09 | 8.97E-05 | 37.4 |
| rs9718104 | 6 | 166170539 | G | T | 0.058 | 0.041 | 0.005 | 9.30E-19 | 1.81E-04 | 75.4 |
| rs17568389 | 7 | 8091876 | T | A | 0.487 | 0.015 | 0.002 | 8.60E-13 | 1.43E-04 | 59.9 |
| rs10234444 | 7 | 41828527 | G | A | 0.823 | 0.016 | 0.003 | 2.90E-08 | 8.33E-05 | 34.8 |
| rs6973656 | 7 | 77422583 | G | A | 0.397 | 0.013 | 0.002 | 1.00E-09 | 9.08E-05 | 37.9 |
| rs973734 | 7 | 89387578 | C | A | 0.152 | 0.017 | 0.003 | 1.50E-08 | 8.05E-05 | 33.6 |
| rs3137 | 7 | 99130875 | G | A | 0.846 | 0.017 | 0.003 | 1.40E-08 | 7.58E-05 | 31.6 |
| rs2694151 | 7 | 101902180 | C | T | 0.058 | 0.026 | 0.005 | 2.30E-08 | 8.77E-05 | 36.6 |
| rs62471080 | 7 | 109128060 | C | G | 0.459 | 0.013 | 0.002 | 3.70E-09 | 9.08E-05 | 37.9 |
| rs7788008 | 7 | 112972483 | G | A | 0.566 | 0.013 | 0.002 | 5.90E-09 | 8.43E-05 | 35.2 |
| rs10246289 | 7 | 115544763 | A | G | 0.113 | 0.019 | 0.003 | 1.70E-08 | 7.71E-05 | 32.2 |
| rs11763734 | 7 | 126400910 | C | A | 0.490 | 0.012 | 0.002 | 8.20E-09 | 8.64E-05 | 36.1 |
| rs7834121 | 8 | 10770255 | T | G | 0.496 | 0.014 | 0.002 | 1.10E-10 | 1.03E-04 | 43.1 |
| rs1421334 | 8 | 30865733 | A | C | 0.450 | 0.017 | 0.002 | 2.20E-15 | 1.58E-04 | 66.0 |
| rs6996198 | 8 | 65463442 | T | C | 0.157 | 0.016 | 0.003 | 3.50E-08 | 8.30E-05 | 34.6 |
| rs6472942 | 8 | 76827190 | C | T | 0.432 | 0.013 | 0.002 | 1.80E-09 | 8.53E-05 | 35.6 |
| rs11985394 | 8 | 92667521 | T | A | 0.545 | 0.012 | 0.002 | 1.30E-08 | 7.39E-05 | 30.8 |
| rs72671494 | 8 | 93195457 | C | T | 0.141 | 0.017 | 0.003 | 4.50E-08 | 6.92E-05 | 28.9 |
| rs12541615 | 8 | 118870151 | C | T | 0.183 | 0.018 | 0.003 | 3.90E-10 | 9.26E-05 | 38.7 |
| rs2717559 | 8 | 143882420 | A | G | 0.559 | 0.012 | 0.002 | 1.50E-08 | 6.94E-05 | 29.0 |
| rs2616830 | 9 | 1721385 | G | A | 0.462 | 0.016 | 0.002 | 2.90E-14 | 1.27E-04 | 53.0 |
| rs2291681 | 9 | 14790841 | G | T | 0.584 | 0.013 | 0.002 | 7.90E-09 | 7.73E-05 | 32.3 |
| rs34864022 | 9 | 22609110 | G | A | 0.066 | 0.026 | 0.004 | 1.40E-09 | 9.14E-05 | 38.2 |
| rs12554512 | 9 | 23352293 | T | C | 0.584 | 0.021 | 0.002 | 3.80E-21 | 2.41E-04 | 100.7 |
| rs2791574 | 9 | 82212094 | G | T | 0.343 | 0.013 | 0.002 | 3.00E-08 | 7.55E-05 | 31.5 |
| rs141184308 | 9 | 86463339 | A | G | 0.978 | 0.043 | 0.008 | 2.60E-08 | 7.91E-05 | 33.0 |
| rs870151 | 9 | 92230747 | A | T | 0.472 | 0.016 | 0.002 | 8.80E-13 | 1.41E-04 | 58.9 |
| rs2787374 | 9 | 103054951 | T | C | 0.408 | 0.012 | 0.002 | 1.20E-08 | 7.07E-05 | 29.5 |
| rs10739499 | 9 | 120514261 | C | G | 0.337 | 0.014 | 0.002 | 1.50E-09 | 9.39E-05 | 39.2 |
| rs7043521 | 9 | 124602779 | A | T | 0.431 | 0.014 | 0.002 | 6.50E-11 | 1.15E-04 | 48.1 |
| rs12342024 | 9 | 126134232 | T | G | 0.114 | 0.021 | 0.003 | 1.70E-09 | 8.60E-05 | 35.9 |
| rs4382592 | 9 | 134870755 | T | G | 0.301 | 0.014 | 0.002 | 6.70E-09 | 9.15E-05 | 38.2 |
| rs2073869 | 9 | 135763816 | C | T | 0.834 | 0.019 | 0.003 | 1.50E-10 | 1.05E-04 | 43.7 |
| 9:139926402_T_C | 9 | 139926402 | C | T | 0.634 | 0.013 | 0.002 | 2.00E-08 | 0.00007 | 30.7 |
| rs1623025 | 10 | 11164813 | C | G | 0.471 | 0.012 | 0.002 | 1.40E-08 | 8.98E-05 | 37.5 |
| rs1243182 | 10 | 21916728 | T | C | 0.310 | 0.019 | 0.002 | 2.00E-15 | 1.61E-04 | 67.3 |
| rs2045147 | 10 | 56660437 | A | G | 0.449 | 0.013 | 0.002 | 5.90E-09 | 6.84E-05 | 28.6 |
| rs10994943 | 10 | 63591413 | T | G | 0.581 | 0.013 | 0.002 | 3.10E-09 | 9.21E-05 | 38.5 |
| rs11201422 | 10 | 86924483 | T | C | 0.669 | 0.013 | 0.002 | 3.40E-08 | 6.74E-05 | 28.1 |
| rs10786658 | 10 | 103737193 | T | A | 0.586 | 0.014 | 0.002 | 4.60E-11 | 1.11E-04 | 46.5 |
| rs7089973 | 10 | 116569565 | A | C | 0.381 | 0.013 | 0.002 | 2.40E-09 | 8.35E-05 | 34.9 |
| rs11245482 | 10 | 126733546 | C | T | 0.387 | 0.013 | 0.002 | 2.60E-09 | 8.79E-05 | 36.7 |
| rs17727474 | 10 | 127182959 | C | T | 0.832 | 0.018 | 0.003 | 3.10E-09 | 8.69E-05 | 36.3 |
| rs12289262 | 11 | 12894758 | T | C | 0.269 | 0.014 | 0.002 | 2.00E-08 | 8.63E-05 | 36.0 |
| rs801733 | 11 | 65934549 | A | C | 0.641 | 0.017 | 0.002 | 7.30E-14 | 1.44E-04 | 60.2 |
| rs116175338 | 11 | 72467875 | T | G | 0.246 | 0.014 | 0.003 | 1.40E-08 | 7.32E-05 | 30.6 |
| rs12272012 | 11 | 84861286 | C | G | 0.044 | 0.030 | 0.005 | 1.90E-08 | 7.18E-05 | 30.0 |
| rs11020045 | 11 | 92487907 | C | A | 0.331 | 0.013 | 0.002 | 1.60E-08 | 8.68E-05 | 36.2 |
| rs17207890 | 11 | 95490754 | G | A | 0.657 | 0.016 | 0.002 | 6.70E-12 | 1.26E-04 | 52.4 |
| rs10791546 | 11 | 101830126 | C | T | 0.589 | 0.012 | 0.002 | 4.10E-08 | 6.74E-05 | 28.1 |
| rs113584870 | 11 | 107243922 | G | A | 0.824 | 0.017 | 0.003 | 1.40E-08 | 0.00008 | 33.2 |
| rs328277 | 11 | 110414195 | C | A | 0.398 | 0.012 | 0.002 | 4.50E-08 | 5.70E-05 | 23.8 |
| rs648044 | 11 | 114030799 | G | A | 0.605 | 0.014 | 0.002 | 1.00E-09 | 1.08E-04 | 45.0 |
| rs11218575 | 11 | 122154576 | C | T | 0.566 | 0.015 | 0.002 | 2.10E-12 | 1.10E-04 | 46.1 |
| rs74802478 | 11 | 131966907 | G | A | 0.819 | 0.017 | 0.003 | 3.50E-09 | 7.72E-05 | 32.2 |
| rs4937842 | 11 | 133734542 | C | G | 0.366 | 0.013 | 0.002 | 1.40E-08 | 7.94E-05 | 33.2 |
| rs10772643 | 12 | 13415288 | C | T | 0.108 | 0.025 | 0.003 | 1.30E-12 | 1.17E-04 | 48.8 |
| rs74996610 | 12 | 24075007 | G | C | 0.046 | 0.031 | 0.005 | 4.10E-09 | 7.97E-05 | 33.3 |
| rs10771746 | 12 | 30791864 | T | C | 0.283 | 0.014 | 0.002 | 2.50E-09 | 9.61E-05 | 40.1 |
| rs10876864 | 12 | 56401085 | A | G | 0.573 | 0.013 | 0.002 | 1.00E-09 | 9.57E-05 | 40.0 |
| rs8756 | 12 | 66359752 | A | C | 0.515 | 0.013 | 0.002 | 5.30E-10 | 9.62E-05 | 40.2 |
| rs11179838 | 12 | 74321123 | C | G | 0.644 | 0.013 | 0.002 | 2.10E-08 | 7.70E-05 | 32.1 |
| rs2164744 | 12 | 109872039 | C | T | 0.365 | 0.013 | 0.002 | 1.10E-08 | 7.67E-05 | 32.0 |
| rs2173650 | 12 | 117525235 | G | T | 0.852 | 0.018 | 0.003 | 4.60E-09 | 8.91E-05 | 37.2 |
| rs1790135 | 12 | 123669235 | T | C | 0.756 | 0.014 | 0.003 | 2.60E-08 | 8.66E-05 | 36.2 |
| rs9563168 | 13 | 54247827 | G | A | 0.791 | 0.018 | 0.003 | 4.30E-11 | 1.11E-04 | 46.3 |
| rs9569734 | 13 | 58319476 | A | G | 0.844 | 0.019 | 0.003 | 3.40E-10 | 1.11E-04 | 46.4 |
| rs9528023 | 13 | 60226991 | C | T | 0.724 | 0.013 | 0.002 | 3.80E-08 | 8.54E-05 | 35.6 |
| rs56858768 | 13 | 86511730 | A | G | 0.299 | 0.015 | 0.002 | 3.40E-10 | 1.04E-04 | 43.4 |
| rs553397860 | 13 | 99051118 | A | G | 0.845 | 0.017 | 0.003 | 8.90E-09 | 7.82E-05 | 32.7 |
| rs7991062 | 13 | 100713194 | G | C | 0.341 | 0.018 | 0.002 | 8.80E-15 | 1.55E-04 | 64.8 |
| rs11629120 | 14 | 23458457 | C | T | 0.414 | 0.013 | 0.002 | 1.10E-08 | 8.37E-05 | 34.9 |
| rs405797 | 14 | 26972834 | A | T | 0.753 | 0.015 | 0.003 | 2.80E-09 | 9.92E-05 | 41.4 |
| rs1022785 | 14 | 29571653 | G | A | 0.142 | 0.018 | 0.003 | 7.10E-09 | 1.01E-04 | 42.4 |
| rs10145592 | 14 | 94287215 | G | C | 0.591 | 0.015 | 0.002 | 1.80E-11 | 1.13E-04 | 47.0 |
| rs7157001 | 14 | 99749484 | G | A | 0.256 | 0.014 | 0.002 | 2.30E-08 | 7.76E-05 | 32.4 |
| rs8043253 | 15 | 41503730 | T | C | 0.427 | 0.012 | 0.002 | 3.40E-08 | 7.98E-05 | 33.3 |
| rs2460 | 15 | 53073084 | A | G | 0.264 | 0.015 | 0.002 | 5.00E-10 | 1.02E-04 | 42.4 |
| rs4775373 | 15 | 61476936 | T | C | 0.361 | 0.013 | 0.002 | 4.70E-09 | 9.02E-05 | 37.7 |
| rs61331678 | 15 | 77995949 | G | C | 0.568 | 0.015 | 0.002 | 6.10E-12 | 1.13E-04 | 47.2 |
| rs35574015 | 16 | 15146061 | C | T | 0.294 | 0.013 | 0.002 | 3.00E-08 | 7.84E-05 | 32.7 |
| rs7189927 | 16 | 28913787 | T | C | 0.356 | 0.015 | 0.002 | 3.40E-11 | 1.17E-04 | 48.7 |
| rs749671 | 16 | 31088347 | G | A | 0.629 | 0.016 | 0.002 | 2.70E-12 | 1.26E-04 | 52.5 |
| rs7184800 | 16 | 53509131 | G | A | 0.697 | 0.017 | 0.002 | 8.20E-13 | 1.38E-04 | 57.7 |
| rs142710267 | 16 | 72169262 | T | G | 0.650 | 0.016 | 0.002 | 6.60E-12 | 1.08E-04 | 45.1 |
| rs2447098 | 17 | 2277720 | A | C | 0.525 | 0.015 | 0.002 | 6.80E-12 | 1.19E-04 | 49.7 |
| rs11654952 | 17 | 4813799 | G | T | 0.139 | 0.017 | 0.003 | 4.30E-08 | 6.59E-05 | 27.5 |
| rs2584597 | 17 | 61941284 | T | C | 0.662 | 0.015 | 0.002 | 2.90E-10 | 9.49E-05 | 39.6 |
| rs9902312 | 17 | 65070304 | T | C | 0.683 | 0.015 | 0.002 | 4.50E-11 | 9.79E-05 | 40.9 |
| rs11657730 | 17 | 79372489 | C | T | 0.637 | 0.013 | 0.002 | 1.10E-08 | 9.44E-05 | 39.4 |
| rs303753 | 18 | 21074922 | A | G | 0.347 | 0.014 | 0.002 | 2.70E-10 | 1.08E-04 | 45.3 |
| rs71362609 | 18 | 23104382 | C | T | 0.486 | 0.012 | 0.002 | 2.80E-08 | 7.49E-05 | 31.3 |
| rs11081851 | 18 | 31783262 | A | G | 0.562 | 0.013 | 0.002 | 1.00E-08 | 7.61E-05 | 31.8 |
| rs9964724 | 18 | 35159124 | C | T | 0.318 | 0.018 | 0.002 | 3.30E-14 | 1.70E-04 | 71.2 |
| rs1156541 | 18 | 39952989 | C | T | 0.223 | 0.014 | 0.003 | 2.80E-08 | 6.15E-05 | 25.7 |
| rs148544378 | 18 | 40323567 | C | T | 0.979 | 0.053 | 0.008 | 2.20E-12 | 1.30E-04 | 54.4 |
| rs17512836 | 18 | 53194961 | T | C | 0.974 | 0.042 | 0.007 | 7.00E-10 | 9.30E-05 | 38.8 |
| rs7248205 | 19 | 10770305 | C | T | 0.398 | 0.014 | 0.002 | 3.40E-10 | 1.09E-04 | 45.4 |
| rs111901094 | 19 | 19513570 | T | G | 0.183 | 0.017 | 0.003 | 2.00E-09 | 1.00E-04 | 41.7 |
| rs6131281 | 20 | 11891724 | C | T | 0.597 | 0.016 | 0.002 | 3.10E-13 | 1.33E-04 | 55.4 |
| rs6141814 | 20 | 31368960 | A | C | 0.387 | 0.014 | 0.002 | 1.30E-09 | 9.34E-05 | 39.0 |
| rs4810312 | 20 | 39968707 | G | C | 0.421 | 0.013 | 0.002 | 1.20E-08 | 8.45E-05 | 35.3 |
| rs6124476 | 20 | 41202830 | T | C | 0.589 | 0.012 | 0.002 | 1.80E-08 | 8.06E-05 | 33.6 |
| rs6031440 | 20 | 42809317 | A | T | 0.398 | 0.012 | 0.002 | 1.80E-08 | 7.75E-05 | 32.4 |
| rs55700114 | 20 | 43717080 | A | G | 0.287 | 0.014 | 0.002 | 2.10E-09 | 1.00E-04 | 41.9 |
| rs6125906 | 20 | 48729803 | A | C | 0.092 | 0.021 | 0.004 | 3.90E-08 | 7.16E-05 | 29.9 |
| rs56103247 | 20 | 62483184 | C | T | 0.944 | 0.030 | 0.005 | 3.80E-10 | 9.35E-05 | 39.1 |
| rs10427502 | 21 | 40654840 | G | A | 0.622 | 0.014 | 0.002 | 8.60E-10 | 8.34E-05 | 34.8 |
| rs7609 | 22 | 29951650 | A | T | 0.231 | 0.015 | 0.003 | 3.60E-09 | 8.58E-05 | 35.8 |
| rs2092829 | 22 | 31780514 | G | A | 0.706 | 0.014 | 0.002 | 1.30E-08 | 7.24E-05 | 30.2 |
| rs150399729 | 22 | 42969453 | C | G | 0.969 | 0.039 | 0.007 | 3.60E-08 | 6.98E-05 | 29.2 |
| Chr chromosome, EA effect allele, EAF effect allele frequency, OA other allele, se standard error, SNP single nucleotide polymorphism  ^a^ Position based on GRCh37 | | | | | | | | | | |

| **Supplemental Table 2: Summary information on computer use SNPs used as genetic instruments used for the Mendelian randomisation analyses** | | | | | | | | | | |
| --- | --- | --- | --- | --- | --- | --- | --- | --- | --- | --- |
| SNP | CHR | POS | EA | OA | EAF | beta | se | P_value | R^2^ | F_statistic |
| rs2748985 | 1 | 1853184 | C | T | 0.545 | 0.015 | 0.002 | 4.10E-12 | 1.22E-04 | 50.7 |
| rs12145677 | 1 | 110023610 | A | G | 0.298 | 0.017 | 0.002 | 1.20E-12 | 1.27E-04 | 52.8 |
| rs7526456 | 1 | 197773734 | C | T | 0.587 | 0.013 | 0.002 | 1.20E-08 | 8.36E-05 | 34.7 |
| rs10754920 | 2 | 146456888 | C | G | 0.112 | 0.020 | 0.003 | 6.70E-09 | 7.59E-05 | 31.5 |
| rs6744254 | 2 | 155986195 | T | C | 0.529 | 0.016 | 0.002 | 5.40E-13 | 1.27E-04 | 52.5 |
| rs11274218 | 2 | 215035001 | T | C | 0.389 | 0.014 | 0.002 | 2.50E-09 | 8.63E-05 | 35.8 |
| rs4592851 | 2 | 215364232 | T | G | 0.250 | 0.014 | 0.003 | 1.40E-08 | 8.99E-05 | 37.3 |
| rs10208088 | 2 | 221055873 | C | T | 0.419 | 0.013 | 0.002 | 4.40E-09 | 9.21E-05 | 38.2 |
| rs11708955 | 3 | 49540114 | C | T | 0.305 | 0.016 | 0.002 | 4.20E-11 | 1.14E-04 | 47.2 |
| rs6774533 | 3 | 62471086 | T | C | 0.706 | 0.015 | 0.002 | 1.20E-09 | 9.14E-05 | 37.9 |
| rs78082503 | 3 | 116556913 | C | G | 0.214 | 0.015 | 0.003 | 1.10E-08 | 7.38E-05 | 30.6 |
| rs6857629 | 4 | 39684977 | G | A | 0.755 | 0.015 | 0.003 | 1.90E-09 | 9.25E-05 | 38.4 |
| rs2068625 | 4 | 159856739 | C | T | 0.698 | 0.016 | 0.002 | 4.00E-11 | 1.08E-04 | 44.8 |
| rs2220599 | 5 | 7378854 | G | C | 0.367 | 0.016 | 0.002 | 2.40E-12 | 1.15E-04 | 47.6 |
| rs11749912 | 5 | 88065628 | A | G | 0.423 | 0.014 | 0.002 | 4.90E-10 | 9.48E-05 | 39.3 |
| rs1469249 | 5 | 113837198 | G | A | 0.789 | 0.016 | 0.003 | 9.50E-09 | 8.09E-05 | 33.6 |
| rs162894 | 5 | 131611872 | T | G | 0.332 | 0.013 | 0.002 | 1.10E-08 | 7.66E-05 | 31.8 |
| rs56158102 | 5 | 144027803 | C | T | 0.344 | 0.013 | 0.002 | 3.10E-08 | 7.49E-05 | 31.1 |
| rs9477970 | 6 | 19082663 | A | T | 0.192 | 0.016 | 0.003 | 1.00E-08 | 8.15E-05 | 33.8 |
| rs2179517 | 6 | 26198845 | C | G | 0.449 | 0.012 | 0.002 | 1.90E-08 | 9.25E-05 | 38.4 |
| rs9372625 | 6 | 98344031 | A | G | 0.381 | 0.018 | 0.002 | 5.70E-16 | 1.65E-04 | 68.3 |
| rs17789218 | 6 | 100600097 | C | T | 0.244 | 0.015 | 0.003 | 4.20E-09 | 8.57E-05 | 35.6 |
| rs6935828 | 6 | 140811367 | T | C | 0.554 | 0.012 | 0.002 | 4.30E-08 | 7.02E-05 | 29.1 |
| rs55772938 | 7 | 70025113 | G | A | 0.295 | 0.015 | 0.002 | 3.90E-10 | 1.02E-04 | 42.4 |
| rs12706626 | 7 | 124531370 | A | G | 0.383 | 0.013 | 0.002 | 1.40E-08 | 8.13E-05 | 33.7 |
| rs3944151 | 7 | 126970005 | A | G | 0.275 | 0.014 | 0.002 | 3.10E-08 | 7.20E-05 | 29.9 |
| rs2345941 | 7 | 133337635 | A | G | 0.550 | 0.015 | 0.002 | 4.00E-11 | 1.16E-04 | 48.3 |
| rs13262595 | 8 | 143316970 | G | A | 0.560 | 0.016 | 0.002 | 1.20E-12 | 1.23E-04 | 51.1 |
| rs4977839 | 9 | 23355310 | A | G | 0.416 | 0.020 | 0.002 | 4.70E-19 | 2.01E-04 | 83.5 |
| rs113851275 | 9 | 98297220 | A | G | 0.108 | 0.021 | 0.004 | 3.20E-09 | 8.60E-05 | 35.7 |
| rs7020477 | 9 | 116827760 | A | G | 0.735 | 0.014 | 0.002 | 1.20E-08 | 7.81E-05 | 32.4 |
| rs73578186 | 9 | 126334485 | C | T | 0.675 | 0.015 | 0.002 | 2.30E-10 | 9.66E-05 | 40.1 |
| rs2734849 | 11 | 113270160 | G | A | 0.509 | 0.014 | 0.002 | 8.20E-10 | 9.70E-05 | 40.3 |
| rs1448355 | 11 | 131286685 | T | C | 0.619 | 0.015 | 0.002 | 1.30E-10 | 9.83E-05 | 40.8 |
| rs35933007 | 12 | 38929341 | A | G | 0.228 | 0.015 | 0.003 | 9.20E-09 | 8.79E-05 | 36.5 |
| rs1319734 | 12 | 40367517 | A | G | 0.601 | 0.013 | 0.002 | 1.90E-08 | 7.72E-05 | 32.1 |
| rs206965 | 12 | 120856332 | T | C | 0.207 | 0.016 | 0.003 | 9.10E-09 | 8.19E-05 | 34.0 |
| rs12874776 | 13 | 57794345 | A | C | 0.107 | 0.023 | 0.004 | 8.00E-11 | 1.01E-04 | 41.8 |
| rs198262 | 14 | 57283283 | C | T | 0.044 | 0.030 | 0.005 | 3.90E-08 | 7.19E-05 | 29.8 |
| rs166835 | 15 | 47716037 | C | T | 0.443 | 0.013 | 0.002 | 3.40E-09 | 8.96E-05 | 37.2 |
| rs4702 | 15 | 91426560 | G | A | 0.444 | 0.012 | 0.002 | 3.80E-08 | 7.30E-05 | 30.3 |
| rs6498759 | 16 | 18020295 | T | C | 0.310 | 0.013 | 0.002 | 2.70E-08 | 8.10E-05 | 33.6 |
| rs3730399 | 16 | 67229019 | A | G | 0.935 | 0.025 | 0.004 | 3.30E-08 | 7.72E-05 | 32.0 |
| rs4073003 | 17 | 19148305 | A | G | 0.874 | 0.020 | 0.003 | 1.20E-09 | 9.54E-05 | 39.6 |
| rs7209653 | 17 | 19882084 | T | C | 0.704 | 0.016 | 0.002 | 9.10E-11 | 9.97E-05 | 41.4 |
| rs12603813 | 17 | 43196584 | T | C | 0.746 | 0.014 | 0.003 | 1.30E-08 | 7.98E-05 | 33.1 |
| rs66643547 | 17 | 79375315 | T | C | 0.353 | 0.015 | 0.002 | 3.20E-11 | 1.09E-04 | 45.3 |
| rs784256 | 18 | 53398626 | G | A | 0.188 | 0.019 | 0.003 | 6.20E-11 | 1.04E-04 | 43.3 |
| rs6129084 | 20 | 59828857 | T | A | 0.641 | 0.014 | 0.002 | 9.30E-10 | 8.92E-05 | 37.0 |
| rs9608249 | 22 | 24613952 | G | A | 0.851 | 0.017 | 0.003 | 2.20E-08 | 7.49E-05 | 31.1 |
| rs136553 | 22 | 27255675 | T | C | 0.376 | 0.015 | 0.002 | 2.30E-11 | 1.11E-04 | 46.1 |
| rs7288455 | 22 | 39966547 | A | G | 0.436 | 0.013 | 0.002 | 4.00E-09 | 8.55E-05 | 35.5 |
| Chr chromosome, EA effect allele, EAF effect allele frequency, OA other allele, se standard error, SNP single nucleotide polymorphism  ^a^ Position based on GRCh37 | | | | | | | | | | |

| **Supplemental Table 3: Summary information on breast cancer risk for the leisure television watching SNPs used for the Mendelian randomisation analyses** | | | | | | | | | | | | | | |
| --- | --- | --- | --- | --- | --- | --- | --- | --- | --- | --- | --- | --- | --- | --- |
| SNP | EA | OA | brst_beta | brst_se | Lum_A_beta | Lum_A_se | Lum_B_beta | Lum_B_se | HER2Neg_beta | HER2Neg_se | HER2_Enriched_beta | HER2_Enriched_se | 3N_beta | 3N_se |
| rs11810109 | A | T | 0.005 | 0.007 | -0.001 | 0.009 | 0.020 | 0.019 | 0.016 | 0.016 | 0.000 | 0.028 | 0.026 | 0.017 |
| rs112632642 | A | G | 0.007 | 0.012 | 0.011 | 0.016 | -0.002 | 0.035 | -0.052 | 0.031 | 0.076 | 0.049 | 0.005 | 0.030 |
| rs6670915 | C | T | 0.005 | 0.006 | -0.002 | 0.008 | 0.004 | 0.018 | 0.014 | 0.015 | -0.027 | 0.026 | 0.011 | 0.015 |
| rs12725114 | G | A | 0.007 | 0.007 | 0.016 | 0.010 | 0.029 | 0.022 | 0.002 | 0.018 | 0.040 | 0.032 | -0.004 | 0.019 |
| rs984409 | A | G | 0.015 | 0.006 | 0.019 | 0.008 | 0.006 | 0.019 | 0.032 | 0.016 | -0.022 | 0.027 | 0.025 | 0.016 |
| rs10890123 | C | T | 0.002 | 0.008 | 0.019 | 0.010 | -0.012 | 0.023 | 0.012 | 0.019 | -0.019 | 0.033 | -0.016 | 0.020 |
| rs71658797 | A | T | 0.004 | 0.010 | 0.003 | 0.013 | 0.034 | 0.029 | -0.059 | 0.025 | 0.038 | 0.042 | -0.002 | 0.025 |
| rs114328297 | T | G | -0.006 | 0.008 | 0.001 | 0.010 | -0.021 | 0.023 | -0.019 | 0.020 | 0.053 | 0.034 | -0.033 | 0.020 |
| rs631130 | A | T | 0.006 | 0.007 | 0.013 | 0.009 | 0.007 | 0.021 | -0.006 | 0.018 | -0.042 | 0.030 | 0.016 | 0.018 |
| rs148648822 | T | G | 0.005 | 0.009 | -0.004 | 0.011 | 0.006 | 0.025 | 0.011 | 0.021 | -0.022 | 0.037 | 0.008 | 0.021 |
| rs17379561 | T | A | 0.002 | 0.009 | 0.001 | 0.011 | -0.052 | 0.025 | 0.029 | 0.021 | -0.010 | 0.036 | 0.008 | 0.021 |
| rs55909997 | A | G | 0.002 | 0.006 | -0.002 | 0.008 | -0.016 | 0.019 | 0.024 | 0.016 | 0.005 | 0.027 | 0.024 | 0.016 |
| rs1278847 | C | A | -0.012 | 0.007 | -0.016 | 0.009 | -0.008 | 0.019 | -0.004 | 0.017 | -0.017 | 0.028 | 0.011 | 0.017 |
| rs141425667 | A | G | -0.016 | 0.014 | -0.017 | 0.018 | -0.013 | 0.038 | -0.047 | 0.034 | -0.029 | 0.057 | 0.032 | 0.033 |
| rs4845364 | G | A | -0.005 | 0.006 | -0.007 | 0.008 | -0.002 | 0.018 | 0.002 | 0.015 | -0.018 | 0.026 | -0.020 | 0.015 |
| rs6673341 | G | T | 0.000 | 0.006 | 0.006 | 0.008 | 0.004 | 0.018 | -0.007 | 0.015 | 0.005 | 0.026 | 0.026 | 0.015 |
| rs72725224 | C | A | -0.034 | 0.025 | -0.023 | 0.032 | -0.056 | 0.070 | -0.038 | 0.061 | -0.046 | 0.101 | -0.042 | 0.062 |
| rs10737620 | T | A | 0.011 | 0.007 | 0.022 | 0.009 | 0.017 | 0.020 | 0.012 | 0.017 | 0.012 | 0.029 | -0.017 | 0.017 |
| rs6721975 | C | T | 0.012 | 0.008 | 0.008 | 0.010 | -0.018 | 0.022 | -0.008 | 0.019 | 0.031 | 0.033 | 0.025 | 0.020 |
| rs11896615 | G | A | -0.002 | 0.009 | 0.004 | 0.011 | 0.020 | 0.025 | -0.022 | 0.021 | -0.019 | 0.036 | -0.024 | 0.021 |
| rs72781699 | A | G | -0.011 | 0.008 | -0.006 | 0.010 | -0.016 | 0.022 | 0.015 | 0.019 | 0.007 | 0.032 | -0.043 | 0.019 |
| rs12105701 | T | C | 0.003 | 0.006 | 0.004 | 0.008 | 0.020 | 0.018 | -0.043 | 0.016 | -0.026 | 0.026 | 0.027 | 0.016 |
| rs7564130 | C | T | 0.002 | 0.006 | 0.010 | 0.008 | 0.016 | 0.018 | 0.005 | 0.016 | -0.005 | 0.026 | -0.006 | 0.016 |
| rs11125515 | A | C | 0.005 | 0.007 | -0.001 | 0.009 | 0.016 | 0.020 | 0.017 | 0.017 | 0.057 | 0.028 | 0.013 | 0.017 |
| rs12475168 | T | G | 0.008 | 0.009 | 0.002 | 0.012 | 0.052 | 0.025 | 0.017 | 0.022 | 0.036 | 0.037 | -0.001 | 0.022 |
| rs183566425 | A | T | -0.023 | 0.031 | -0.076 | 0.039 | 0.088 | 0.094 | 0.066 | 0.080 | 0.324 | 0.156 | -0.082 | 0.075 |
| rs10189857 | G | A | 0.017 | 0.006 | 0.019 | 0.008 | 0.011 | 0.018 | 0.022 | 0.015 | 0.008 | 0.026 | 0.015 | 0.015 |
| rs62641636 | A | G | 0.001 | 0.007 | -0.003 | 0.009 | 0.039 | 0.019 | 0.009 | 0.016 | -0.042 | 0.028 | 0.016 | 0.017 |
| rs59210467 | T | C | 0.000 | 0.007 | 0.011 | 0.009 | -0.025 | 0.020 | -0.007 | 0.017 | 0.049 | 0.028 | 0.012 | 0.017 |
| rs11689199 | A | G | -0.010 | 0.006 | -0.008 | 0.008 | -0.016 | 0.018 | -0.007 | 0.016 | -0.014 | 0.027 | -0.017 | 0.016 |
| rs2916251 | T | G | 0.010 | 0.006 | 0.008 | 0.008 | -0.018 | 0.018 | 0.003 | 0.016 | 0.004 | 0.027 | 0.018 | 0.016 |
| rs1451533 | A | G | 0.004 | 0.007 | 0.015 | 0.009 | -0.032 | 0.021 | 0.001 | 0.018 | -0.018 | 0.031 | 0.005 | 0.018 |
| rs2971640 | A | G | -0.010 | 0.006 | -0.008 | 0.008 | -0.016 | 0.018 | -0.018 | 0.015 | -0.013 | 0.026 | -0.002 | 0.015 |
| rs12476388 | C | T | -0.002 | 0.007 | 0.002 | 0.009 | 0.009 | 0.019 | -0.004 | 0.017 | -0.035 | 0.028 | -0.014 | 0.017 |
| rs374722 | G | A | 0.011 | 0.009 | 0.018 | 0.012 | 0.036 | 0.025 | -0.012 | 0.022 | -0.080 | 0.039 | 0.001 | 0.022 |
| rs3754970 | C | T | -0.007 | 0.006 | 0.000 | 0.008 | 0.003 | 0.018 | -0.006 | 0.015 | 0.017 | 0.026 | -0.023 | 0.016 |
| rs1913808 | G | A | 0.010 | 0.007 | 0.003 | 0.009 | 0.052 | 0.019 | -0.012 | 0.016 | 0.001 | 0.028 | 0.001 | 0.016 |
| rs2060199 | T | A | -0.007 | 0.006 | -0.007 | 0.008 | 0.010 | 0.018 | 0.019 | 0.016 | 0.011 | 0.026 | -0.021 | 0.016 |
| rs263771 | A | C | -0.003 | 0.007 | -0.001 | 0.010 | -0.016 | 0.021 | -0.007 | 0.018 | -0.008 | 0.031 | -0.028 | 0.018 |
| rs4577309 | A | G | -0.001 | 0.006 | 0.004 | 0.008 | -0.014 | 0.018 | 0.008 | 0.015 | -0.006 | 0.026 | -0.015 | 0.015 |
| rs34224261 | C | G | 0.000 | 0.006 | 0.006 | 0.008 | -0.004 | 0.017 | 0.026 | 0.015 | 0.060 | 0.026 | 0.001 | 0.015 |
| rs4675246 | T | G | -0.006 | 0.008 | -0.013 | 0.010 | 0.005 | 0.022 | -0.038 | 0.019 | 0.077 | 0.032 | 0.040 | 0.019 |
| rs13029509 | A | G | 0.009 | 0.006 | 0.008 | 0.008 | -0.008 | 0.018 | 0.019 | 0.015 | -0.017 | 0.026 | 0.017 | 0.015 |
| rs10932837 | T | C | -0.001 | 0.006 | -0.003 | 0.008 | 0.011 | 0.018 | -0.009 | 0.015 | 0.023 | 0.026 | 0.000 | 0.015 |
| rs4973576 | A | C | -0.012 | 0.007 | -0.022 | 0.009 | -0.031 | 0.020 | 0.015 | 0.017 | -0.018 | 0.029 | -0.026 | 0.017 |
| rs138256022 | T | C | 0.016 | 0.017 | 0.025 | 0.023 | 0.024 | 0.050 | 0.043 | 0.043 | 0.061 | 0.072 | 0.005 | 0.044 |
| rs9834970 | T | C | 0.001 | 0.006 | 0.001 | 0.008 | 0.005 | 0.018 | 0.004 | 0.016 | -0.035 | 0.027 | 0.011 | 0.016 |
| rs1352890 | T | C | 0.010 | 0.006 | 0.009 | 0.008 | 0.004 | 0.018 | 0.019 | 0.015 | 0.034 | 0.026 | 0.014 | 0.015 |
| rs3796386 | A | G | 0.018 | 0.006 | 0.014 | 0.008 | 0.038 | 0.018 | 0.012 | 0.015 | 0.063 | 0.026 | 0.043 | 0.015 |
| rs11917871 | C | T | 0.005 | 0.006 | 0.017 | 0.008 | 0.009 | 0.019 | -0.015 | 0.016 | -0.047 | 0.027 | 0.025 | 0.016 |
| rs11130793 | C | T | 0.001 | 0.006 | 0.006 | 0.008 | 0.012 | 0.018 | -0.021 | 0.016 | 0.004 | 0.027 | 0.002 | 0.016 |
| rs11714337 | G | A | 0.032 | 0.006 | 0.045 | 0.008 | 0.036 | 0.018 | 0.029 | 0.015 | 0.000 | 0.026 | 0.014 | 0.015 |
| rs12496084 | G | A | -0.002 | 0.007 | -0.008 | 0.010 | -0.018 | 0.021 | 0.015 | 0.018 | -0.069 | 0.031 | 0.016 | 0.018 |
| rs6797840 | C | A | 0.004 | 0.006 | 0.007 | 0.008 | 0.015 | 0.018 | 0.022 | 0.015 | -0.024 | 0.026 | -0.005 | 0.015 |
| rs56398417 | C | T | -0.018 | 0.007 | -0.019 | 0.009 | -0.040 | 0.019 | -0.033 | 0.016 | -0.014 | 0.028 | -0.026 | 0.016 |
| rs2034768 | A | G | -0.003 | 0.006 | -0.006 | 0.008 | 0.010 | 0.018 | 0.016 | 0.015 | 0.016 | 0.025 | -0.025 | 0.015 |
| rs12632778 | G | T | 0.023 | 0.015 | 0.044 | 0.019 | -0.025 | 0.044 | 0.012 | 0.037 | -0.100 | 0.066 | 0.026 | 0.037 |
| rs9867121 | C | A | 0.013 | 0.008 | 0.015 | 0.010 | 0.020 | 0.023 | 0.001 | 0.020 | 0.052 | 0.034 | 0.009 | 0.020 |
| rs12491503 | A | G | 0.001 | 0.007 | -0.008 | 0.009 | 0.018 | 0.019 | 0.029 | 0.016 | 0.025 | 0.028 | -0.009 | 0.017 |
| rs114600294 | C | G | -0.017 | 0.010 | -0.002 | 0.013 | -0.029 | 0.028 | 0.015 | 0.024 | -0.035 | 0.042 | -0.007 | 0.025 |
| rs66852340 | T | C | 0.016 | 0.007 | 0.019 | 0.009 | -0.027 | 0.021 | 0.046 | 0.018 | 0.011 | 0.031 | 0.022 | 0.018 |
| rs7700107 | C | A | -0.005 | 0.009 | -0.033 | 0.012 | 0.019 | 0.025 | 0.026 | 0.022 | 0.066 | 0.036 | 0.009 | 0.022 |
| rs34811474 | G | A | 0.006 | 0.008 | -0.002 | 0.011 | 0.015 | 0.025 | 0.012 | 0.021 | 0.045 | 0.036 | 0.005 | 0.021 |
| rs77215114 | A | T | -0.016 | 0.013 | -0.022 | 0.018 | -0.019 | 0.039 | 0.021 | 0.034 | 0.022 | 0.057 | -0.051 | 0.033 |
| rs28971796 | A | G | 0.010 | 0.009 | 0.017 | 0.012 | 0.042 | 0.026 | 0.005 | 0.022 | 0.049 | 0.037 | 0.017 | 0.022 |
| rs6850494 | C | A | -0.009 | 0.006 | -0.004 | 0.008 | -0.030 | 0.018 | -0.002 | 0.015 | 0.045 | 0.026 | -0.020 | 0.016 |
| rs13107325 | T | C | 0.019 | 0.012 | 0.021 | 0.016 | 0.051 | 0.034 | 0.007 | 0.030 | -0.051 | 0.051 | 0.025 | 0.030 |
| rs62332760 | T | G | -0.017 | 0.006 | -0.018 | 0.008 | -0.016 | 0.018 | -0.024 | 0.016 | 0.039 | 0.026 | 0.021 | 0.016 |
| rs7693703 | G | A | -0.007 | 0.010 | -0.015 | 0.014 | -0.012 | 0.030 | 0.009 | 0.026 | 0.052 | 0.046 | 0.005 | 0.026 |
| rs4334769 | G | T | -0.008 | 0.006 | -0.007 | 0.008 | -0.016 | 0.018 | -0.032 | 0.015 | 0.012 | 0.026 | 0.015 | 0.015 |
| rs6825241 | A | C | -0.002 | 0.006 | -0.009 | 0.008 | 0.008 | 0.018 | 0.018 | 0.015 | -0.008 | 0.026 | -0.006 | 0.016 |
| rs7693082 | G | C | -0.004 | 0.007 | -0.013 | 0.009 | -0.023 | 0.020 | -0.001 | 0.017 | 0.044 | 0.028 | -0.013 | 0.017 |
| rs10940659 | G | A | -0.008 | 0.006 | -0.020 | 0.008 | 0.061 | 0.018 | -0.014 | 0.015 | 0.028 | 0.026 | -0.003 | 0.015 |
| rs2409838 | A | T | -0.002 | 0.006 | -0.010 | 0.008 | 0.007 | 0.018 | 0.009 | 0.015 | 0.015 | 0.026 | -0.011 | 0.016 |
| rs262890 | G | A | 0.003 | 0.007 | 0.002 | 0.009 | -0.032 | 0.019 | 0.006 | 0.016 | 0.021 | 0.028 | 0.008 | 0.017 |
| rs7716447 | G | A | -0.007 | 0.007 | -0.009 | 0.009 | 0.013 | 0.019 | -0.011 | 0.016 | -0.034 | 0.028 | 0.011 | 0.017 |
| rs1031423 | C | T | 0.002 | 0.008 | 0.007 | 0.010 | -0.013 | 0.022 | -0.013 | 0.019 | 0.010 | 0.032 | -0.021 | 0.019 |
| rs57585211 | G | T | -0.001 | 0.008 | 0.000 | 0.011 | -0.021 | 0.024 | 0.036 | 0.020 | -0.044 | 0.035 | 0.026 | 0.020 |
| rs9326862 | A | T | 0.005 | 0.006 | 0.008 | 0.008 | 0.008 | 0.018 | -0.002 | 0.015 | 0.025 | 0.026 | 0.005 | 0.015 |
| rs10041724 | T | C | 0.008 | 0.008 | 0.006 | 0.010 | 0.020 | 0.022 | -0.011 | 0.019 | 0.065 | 0.033 | -0.021 | 0.019 |
| rs10057545 | A | G | 0.006 | 0.006 | -0.002 | 0.008 | 0.030 | 0.018 | 0.007 | 0.016 | 0.003 | 0.027 | 0.014 | 0.016 |
| rs62379379 | T | G | 0.001 | 0.012 | 0.014 | 0.016 | -0.074 | 0.037 | -0.019 | 0.031 | -0.122 | 0.055 | 0.031 | 0.030 |
| rs10054327 | G | A | -0.010 | 0.006 | -0.013 | 0.008 | 0.000 | 0.018 | 0.004 | 0.015 | 0.003 | 0.026 | -0.022 | 0.016 |
| rs42210 | C | G | 0.020 | 0.007 | 0.008 | 0.010 | 0.046 | 0.021 | 0.046 | 0.018 | 0.083 | 0.031 | 0.019 | 0.018 |
| rs2336895 | G | A | 0.007 | 0.007 | 0.013 | 0.009 | -0.008 | 0.020 | 0.020 | 0.017 | -0.019 | 0.029 | 0.001 | 0.017 |
| rs72828890 | C | T | -0.005 | 0.011 | 0.006 | 0.014 | -0.010 | 0.032 | 0.027 | 0.028 | 0.002 | 0.047 | 0.012 | 0.028 |
| rs255292 | A | C | -0.003 | 0.006 | 0.001 | 0.008 | 0.001 | 0.018 | -0.003 | 0.016 | 0.018 | 0.026 | -0.020 | 0.016 |
| rs72834698 | G | A | 0.007 | 0.009 | 0.019 | 0.012 | -0.001 | 0.026 | -0.022 | 0.022 | 0.030 | 0.038 | 0.019 | 0.023 |
| rs3135044 | T | C | 0.004 | 0.006 | -0.003 | 0.008 | 0.018 | 0.018 | -0.002 | 0.015 | 0.010 | 0.026 | -0.021 | 0.016 |
| rs576432906 | C | A | 0.007 | 0.007 | -0.001 | 0.009 | 0.023 | 0.020 | 0.011 | 0.017 | 0.027 | 0.029 | 0.025 | 0.017 |
| rs10947452 | T | C | 0.011 | 0.007 | 0.011 | 0.009 | -0.016 | 0.019 | 0.032 | 0.016 | -0.005 | 0.028 | 0.014 | 0.016 |
| rs9471333 | C | T | 0.000 | 0.006 | 0.012 | 0.009 | -0.008 | 0.019 | -0.039 | 0.016 | -0.044 | 0.027 | 0.011 | 0.016 |
| rs4523073 | G | A | 0.002 | 0.006 | 0.004 | 0.008 | -0.025 | 0.018 | 0.006 | 0.016 | 0.003 | 0.027 | -0.008 | 0.016 |
| rs77589760 | C | G | 0.057 | 0.024 | 0.060 | 0.031 | 0.124 | 0.071 | 0.011 | 0.058 | 0.029 | 0.101 | 0.124 | 0.062 |
| rs6905544 | G | A | 0.011 | 0.006 | 0.016 | 0.008 | 0.007 | 0.018 | 0.012 | 0.015 | -0.005 | 0.026 | 0.012 | 0.016 |
| rs17789218 | T | C | -0.013 | 0.008 | -0.025 | 0.011 | -0.002 | 0.023 | 0.030 | 0.020 | 0.068 | 0.034 | 0.014 | 0.020 |
| rs78394231 | C | T | 0.003 | 0.010 | 0.001 | 0.013 | -0.011 | 0.030 | 0.018 | 0.025 | 0.017 | 0.043 | 0.004 | 0.026 |
| rs2184364 | A | G | 0.007 | 0.007 | 0.009 | 0.010 | 0.009 | 0.022 | -0.004 | 0.018 | -0.035 | 0.031 | 0.023 | 0.019 |
| rs9718104 | G | T | 0.010 | 0.014 | 0.009 | 0.018 | -0.009 | 0.041 | -0.004 | 0.034 | -0.041 | 0.060 | 0.054 | 0.034 |
| rs17568389 | T | A | -0.003 | 0.006 | -0.008 | 0.008 | -0.012 | 0.018 | -0.014 | 0.015 | 0.038 | 0.026 | 0.010 | 0.016 |
| rs10234444 | G | A | 0.011 | 0.008 | 0.001 | 0.011 | 0.031 | 0.025 | 0.005 | 0.021 | -0.069 | 0.035 | 0.021 | 0.022 |
| rs6973656 | G | A | 0.000 | 0.006 | -0.006 | 0.009 | 0.013 | 0.019 | 0.019 | 0.016 | 0.013 | 0.027 | -0.003 | 0.016 |
| rs973734 | C | A | 0.013 | 0.009 | 0.015 | 0.011 | 0.032 | 0.025 | 0.015 | 0.021 | 0.017 | 0.036 | 0.023 | 0.022 |
| rs3137 | G | A | 0.010 | 0.008 | 0.002 | 0.011 | 0.018 | 0.024 | 0.004 | 0.021 | 0.037 | 0.035 | 0.019 | 0.021 |
| rs2694151 | C | T | -0.003 | 0.013 | -0.008 | 0.017 | 0.039 | 0.036 | 0.028 | 0.031 | -0.046 | 0.055 | -0.046 | 0.033 |
| rs62471080 | C | G | -0.005 | 0.006 | -0.006 | 0.008 | -0.027 | 0.018 | -0.008 | 0.015 | -0.004 | 0.026 | 0.004 | 0.015 |
| rs7788008 | G | A | 0.000 | 0.006 | 0.002 | 0.008 | -0.027 | 0.018 | 0.013 | 0.015 | -0.019 | 0.026 | 0.005 | 0.015 |
| rs10246289 | A | G | 0.021 | 0.009 | 0.018 | 0.012 | 0.021 | 0.027 | 0.014 | 0.024 | 0.032 | 0.040 | 0.048 | 0.024 |
| rs11763734 | C | A | -0.003 | 0.006 | 0.003 | 0.008 | -0.011 | 0.018 | -0.009 | 0.015 | 0.006 | 0.026 | -0.024 | 0.015 |
| rs7834121 | T | G | 0.008 | 0.007 | 0.009 | 0.009 | -0.008 | 0.021 | -0.005 | 0.017 | 0.013 | 0.028 | 0.008 | 0.018 |
| rs1421334 | A | C | 0.005 | 0.006 | 0.002 | 0.008 | 0.031 | 0.018 | -0.006 | 0.015 | 0.018 | 0.026 | 0.009 | 0.016 |
| rs6996198 | T | C | 0.001 | 0.008 | 0.001 | 0.011 | 0.002 | 0.024 | 0.002 | 0.020 | -0.065 | 0.036 | 0.036 | 0.020 |
| rs6472942 | C | T | 0.012 | 0.006 | 0.020 | 0.008 | -0.023 | 0.018 | 0.013 | 0.015 | 0.031 | 0.025 | 0.006 | 0.015 |
| rs11985394 | T | A | 0.005 | 0.006 | 0.010 | 0.008 | 0.002 | 0.018 | 0.005 | 0.015 | -0.012 | 0.026 | 0.001 | 0.016 |
| rs72671494 | C | T | -0.001 | 0.009 | 0.006 | 0.012 | 0.034 | 0.027 | -0.030 | 0.024 | 0.007 | 0.040 | -0.025 | 0.024 |
| rs12541615 | C | T | -0.029 | 0.008 | -0.037 | 0.011 | -0.035 | 0.023 | -0.048 | 0.020 | -0.039 | 0.034 | -0.004 | 0.020 |
| rs2717559 | A | G | 0.000 | 0.007 | -0.009 | 0.009 | 0.018 | 0.019 | -0.022 | 0.016 | 0.030 | 0.028 | -0.003 | 0.017 |
| rs2616830 | G | A | 0.003 | 0.006 | 0.006 | 0.008 | -0.001 | 0.018 | 0.012 | 0.015 | -0.052 | 0.026 | 0.001 | 0.015 |
| rs2291681 | G | T | 0.009 | 0.006 | 0.007 | 0.008 | 0.010 | 0.019 | 0.019 | 0.016 | -0.023 | 0.027 | 0.027 | 0.016 |
| rs34864022 | G | A | -0.008 | 0.013 | -0.002 | 0.017 | -0.001 | 0.038 | -0.043 | 0.032 | 0.046 | 0.054 | -0.011 | 0.033 |
| rs12554512 | T | C | -0.006 | 0.006 | -0.001 | 0.008 | -0.038 | 0.019 | -0.006 | 0.016 | 0.017 | 0.027 | -0.018 | 0.016 |
| rs2791574 | G | T | -0.003 | 0.006 | -0.011 | 0.008 | -0.009 | 0.018 | 0.004 | 0.016 | -0.026 | 0.027 | 0.029 | 0.016 |
| rs141184308 | A | G | 0.012 | 0.033 | 0.058 | 0.044 | 0.147 | 0.102 | -0.082 | 0.080 | -0.132 | 0.135 | -0.049 | 0.082 |
| rs870151 | A | T | -0.005 | 0.006 | 0.008 | 0.008 | -0.014 | 0.018 | 0.005 | 0.015 | -0.043 | 0.026 | -0.021 | 0.015 |
| rs2787374 | T | C | -0.004 | 0.006 | -0.010 | 0.008 | 0.001 | 0.018 | 0.042 | 0.015 | -0.015 | 0.026 | 0.000 | 0.015 |
| rs10739499 | C | G | 0.001 | 0.007 | 0.014 | 0.009 | -0.003 | 0.019 | -0.010 | 0.016 | -0.017 | 0.028 | -0.014 | 0.017 |
| rs7043521 | A | T | 0.003 | 0.006 | 0.003 | 0.008 | -0.019 | 0.018 | 0.016 | 0.015 | 0.037 | 0.026 | -0.019 | 0.015 |
| rs12342024 | T | G | 0.002 | 0.010 | -0.001 | 0.013 | 0.000 | 0.028 | -0.015 | 0.025 | 0.012 | 0.041 | -0.013 | 0.025 |
| rs4382592 | T | G | 0.011 | 0.007 | 0.004 | 0.009 | 0.030 | 0.019 | 0.002 | 0.016 | 0.045 | 0.028 | 0.006 | 0.017 |
| rs2073869 | C | T | 0.011 | 0.008 | 0.001 | 0.011 | 0.012 | 0.023 | 0.029 | 0.020 | 0.019 | 0.034 | -0.013 | 0.020 |
| 9:139926402_T_C | C | T | -0.002 | 0.006 | 0.002 | 0.008 | -0.006 | 0.018 | -0.029 | 0.016 | -0.026 | 0.027 | 0.015 | 0.016 |
| rs1623025 | C | G | 0.009 | 0.006 | 0.012 | 0.008 | 0.011 | 0.018 | 0.013 | 0.015 | 0.044 | 0.026 | 0.000 | 0.015 |
| rs1243182 | T | C | 0.056 | 0.006 | 0.081 | 0.008 | 0.059 | 0.019 | 0.051 | 0.016 | 0.016 | 0.027 | -0.042 | 0.016 |
| rs2045147 | A | G | 0.002 | 0.006 | -0.002 | 0.008 | 0.023 | 0.018 | 0.032 | 0.015 | 0.045 | 0.026 | 0.001 | 0.015 |
| rs10994943 | T | G | 0.011 | 0.006 | 0.010 | 0.008 | -0.009 | 0.018 | 0.015 | 0.015 | 0.028 | 0.026 | 0.032 | 0.015 |
| rs11201422 | T | C | -0.015 | 0.007 | -0.007 | 0.009 | -0.025 | 0.019 | -0.027 | 0.016 | -0.059 | 0.028 | 0.007 | 0.017 |
| rs10786658 | T | A | -0.001 | 0.006 | -0.002 | 0.008 | 0.032 | 0.018 | -0.022 | 0.015 | 0.011 | 0.026 | -0.014 | 0.016 |
| rs7089973 | A | C | 0.005 | 0.006 | 0.008 | 0.009 | -0.025 | 0.019 | 0.007 | 0.016 | 0.045 | 0.027 | -0.002 | 0.016 |
| rs11245482 | C | T | 0.005 | 0.006 | 0.003 | 0.008 | 0.037 | 0.018 | -0.004 | 0.015 | 0.016 | 0.026 | -0.022 | 0.016 |
| rs17727474 | C | T | -0.013 | 0.008 | -0.020 | 0.011 | 0.021 | 0.025 | -0.048 | 0.021 | -0.078 | 0.035 | 0.025 | 0.022 |
| rs12289262 | T | C | -0.002 | 0.007 | -0.008 | 0.009 | 0.064 | 0.021 | 0.013 | 0.018 | 0.001 | 0.030 | -0.012 | 0.018 |
| rs801733 | A | C | 0.023 | 0.006 | 0.031 | 0.008 | 0.016 | 0.019 | 0.001 | 0.016 | 0.057 | 0.027 | -0.002 | 0.016 |
| rs116175338 | T | G | 0.017 | 0.007 | 0.028 | 0.009 | -0.007 | 0.021 | 0.004 | 0.018 | 0.016 | 0.030 | 0.025 | 0.018 |
| rs12272012 | C | G | 0.023 | 0.015 | 0.017 | 0.019 | -0.098 | 0.046 | 0.003 | 0.037 | 0.045 | 0.063 | 0.040 | 0.037 |
| rs11020045 | C | A | 0.009 | 0.006 | 0.003 | 0.008 | 0.000 | 0.019 | 0.017 | 0.016 | 0.019 | 0.027 | 0.022 | 0.016 |
| rs17207890 | G | A | 0.017 | 0.006 | 0.033 | 0.008 | 0.000 | 0.019 | 0.039 | 0.016 | -0.011 | 0.027 | 0.019 | 0.016 |
| rs10791546 | C | T | -0.001 | 0.006 | 0.007 | 0.008 | 0.005 | 0.018 | -0.014 | 0.015 | 0.000 | 0.026 | -0.016 | 0.015 |
| rs113584870 | G | A | 0.009 | 0.009 | 0.004 | 0.011 | 0.022 | 0.025 | 0.011 | 0.022 | 0.068 | 0.038 | 0.011 | 0.022 |
| rs328277 | C | A | -0.007 | 0.006 | -0.013 | 0.008 | 0.005 | 0.018 | 0.001 | 0.015 | 0.012 | 0.026 | 0.008 | 0.015 |
| rs648044 | G | A | 0.005 | 0.007 | 0.004 | 0.009 | 0.024 | 0.020 | 0.009 | 0.017 | 0.004 | 0.030 | -0.003 | 0.018 |
| rs11218575 | C | T | 0.009 | 0.006 | 0.000 | 0.009 | 0.024 | 0.019 | 0.001 | 0.016 | 0.063 | 0.027 | 0.008 | 0.016 |
| rs74802478 | G | A | -0.007 | 0.008 | -0.002 | 0.011 | 0.011 | 0.024 | -0.007 | 0.020 | -0.012 | 0.034 | -0.030 | 0.020 |
| rs4937842 | C | G | -0.009 | 0.006 | -0.012 | 0.008 | 0.005 | 0.018 | -0.017 | 0.016 | -0.020 | 0.027 | -0.010 | 0.016 |
| rs10772643 | C | T | 0.008 | 0.010 | 0.022 | 0.013 | -0.012 | 0.030 | -0.005 | 0.025 | -0.044 | 0.044 | -0.017 | 0.026 |
| rs74996610 | G | C | 0.023 | 0.015 | 0.032 | 0.019 | -0.067 | 0.046 | 0.038 | 0.037 | 0.070 | 0.062 | -0.005 | 0.038 |
| rs10771746 | T | C | 0.024 | 0.007 | 0.015 | 0.009 | 0.037 | 0.019 | 0.053 | 0.016 | 0.018 | 0.028 | -0.013 | 0.017 |
| rs10876864 | A | G | -0.004 | 0.006 | -0.008 | 0.008 | -0.040 | 0.018 | 0.011 | 0.015 | -0.008 | 0.026 | 0.024 | 0.015 |
| rs8756 | A | C | 0.003 | 0.006 | -0.004 | 0.008 | -0.009 | 0.018 | 0.007 | 0.015 | -0.019 | 0.025 | 0.023 | 0.015 |
| rs11179838 | C | G | -0.004 | 0.006 | -0.009 | 0.008 | -0.009 | 0.018 | 0.003 | 0.016 | -0.013 | 0.026 | 0.000 | 0.016 |
| rs2164744 | C | T | -0.019 | 0.006 | -0.009 | 0.008 | -0.024 | 0.018 | -0.042 | 0.016 | -0.072 | 0.026 | -0.011 | 0.016 |
| rs2173650 | G | T | 0.016 | 0.009 | 0.014 | 0.014 | 0.012 | 0.028 | 0.009 | 0.025 | 0.036 | 0.042 | 0.043 | 0.026 |
| rs1790135 | T | C | 0.003 | 0.007 | 0.000 | 0.009 | 0.006 | 0.020 | 0.019 | 0.017 | 0.021 | 0.029 | -0.003 | 0.017 |
| rs9563168 | G | A | 0.003 | 0.008 | 0.011 | 0.010 | 0.010 | 0.022 | 0.003 | 0.019 | -0.004 | 0.032 | -0.013 | 0.019 |
| rs9569734 | A | G | 0.002 | 0.008 | 0.010 | 0.011 | -0.034 | 0.024 | 0.014 | 0.021 | 0.002 | 0.036 | 0.023 | 0.021 |
| rs9528023 | C | T | 0.002 | 0.007 | -0.004 | 0.009 | -0.003 | 0.021 | 0.015 | 0.018 | -0.003 | 0.030 | -0.020 | 0.018 |
| rs56858768 | A | G | -0.012 | 0.007 | -0.008 | 0.009 | -0.012 | 0.020 | -0.017 | 0.017 | 0.002 | 0.029 | -0.004 | 0.017 |
| rs553397860 | A | G | -0.013 | 0.009 | -0.023 | 0.011 | -0.004 | 0.025 | 0.006 | 0.021 | 0.045 | 0.037 | -0.005 | 0.022 |
| rs7991062 | G | C | 0.004 | 0.006 | 0.012 | 0.008 | 0.001 | 0.019 | 0.001 | 0.016 | 0.036 | 0.027 | 0.005 | 0.016 |
| rs11629120 | C | T | 0.012 | 0.006 | 0.016 | 0.008 | 0.007 | 0.018 | 0.016 | 0.015 | 0.028 | 0.026 | 0.012 | 0.015 |
| rs405797 | A | T | -0.015 | 0.008 | -0.013 | 0.010 | -0.017 | 0.022 | -0.036 | 0.019 | -0.027 | 0.032 | -0.004 | 0.019 |
| rs1022785 | G | A | 0.008 | 0.009 | 0.009 | 0.011 | 0.007 | 0.025 | -0.014 | 0.022 | -0.026 | 0.037 | 0.032 | 0.022 |
| rs10145592 | G | C | -0.003 | 0.006 | 0.015 | 0.009 | 0.009 | 0.019 | -0.006 | 0.016 | -0.030 | 0.027 | -0.020 | 0.016 |
| rs7157001 | G | A | 0.003 | 0.008 | 0.007 | 0.011 | 0.020 | 0.024 | -0.027 | 0.021 | -0.018 | 0.035 | -0.013 | 0.021 |
| rs8043253 | T | C | 0.004 | 0.006 | 0.018 | 0.008 | -0.016 | 0.018 | -0.012 | 0.015 | 0.004 | 0.026 | 0.008 | 0.015 |
| rs2460 | A | G | 0.000 | 0.007 | 0.006 | 0.010 | 0.001 | 0.021 | 0.007 | 0.018 | -0.024 | 0.031 | -0.024 | 0.019 |
| rs4775373 | T | C | 0.002 | 0.006 | 0.000 | 0.008 | -0.001 | 0.019 | 0.004 | 0.016 | 0.034 | 0.027 | -0.026 | 0.016 |
| rs61331678 | G | C | 0.002 | 0.006 | 0.002 | 0.008 | 0.027 | 0.018 | -0.034 | 0.015 | -0.027 | 0.026 | 0.017 | 0.016 |
| rs35574015 | C | T | 0.007 | 0.007 | 0.000 | 0.009 | 0.023 | 0.020 | 0.008 | 0.017 | -0.026 | 0.030 | 0.015 | 0.018 |
| rs7189927 | T | C | 0.001 | 0.006 | 0.006 | 0.008 | -0.010 | 0.019 | -0.001 | 0.016 | 0.014 | 0.027 | -0.026 | 0.016 |
| rs749671 | G | A | -0.017 | 0.006 | -0.014 | 0.008 | -0.010 | 0.018 | -0.010 | 0.015 | -0.002 | 0.026 | -0.015 | 0.016 |
| rs7184800 | G | A | -0.005 | 0.006 | -0.010 | 0.008 | -0.014 | 0.019 | 0.005 | 0.016 | 0.026 | 0.028 | -0.011 | 0.016 |
| rs142710267 | T | G | 0.003 | 0.007 | 0.007 | 0.009 | 0.026 | 0.019 | -0.014 | 0.016 | 0.030 | 0.028 | 0.019 | 0.017 |
| rs2447098 | A | C | -0.008 | 0.006 | -0.009 | 0.009 | -0.012 | 0.019 | -0.036 | 0.016 | 0.005 | 0.027 | -0.020 | 0.016 |
| rs11654952 | G | T | -0.007 | 0.009 | -0.009 | 0.012 | -0.031 | 0.026 | -0.037 | 0.022 | -0.017 | 0.038 | 0.027 | 0.022 |
| rs2584597 | T | C | -0.006 | 0.007 | -0.008 | 0.009 | 0.007 | 0.020 | -0.013 | 0.017 | -0.016 | 0.028 | 0.010 | 0.017 |
| rs9902312 | T | C | -0.009 | 0.006 | -0.024 | 0.008 | -0.026 | 0.019 | 0.016 | 0.016 | -0.021 | 0.027 | 0.009 | 0.016 |
| rs11657730 | C | T | 0.021 | 0.007 | 0.021 | 0.009 | 0.007 | 0.019 | 0.024 | 0.017 | 0.062 | 0.029 | 0.023 | 0.017 |
| rs303753 | A | G | 0.004 | 0.006 | 0.010 | 0.009 | 0.033 | 0.019 | -0.005 | 0.016 | -0.026 | 0.028 | 0.051 | 0.016 |
| rs71362609 | C | T | 0.014 | 0.006 | 0.011 | 0.008 | 0.019 | 0.018 | 0.045 | 0.015 | -0.009 | 0.026 | -0.001 | 0.015 |
| rs11081851 | A | G | 0.003 | 0.006 | -0.001 | 0.008 | -0.004 | 0.018 | 0.003 | 0.015 | -0.009 | 0.026 | 0.029 | 0.015 |
| rs9964724 | C | T | 0.007 | 0.007 | 0.002 | 0.009 | 0.029 | 0.019 | 0.007 | 0.017 | -0.043 | 0.029 | 0.009 | 0.017 |
| rs1156541 | C | T | -0.018 | 0.007 | -0.012 | 0.009 | -0.043 | 0.021 | -0.029 | 0.018 | -0.010 | 0.030 | -0.044 | 0.018 |
| rs17512836 | T | C | -0.008 | 0.019 | -0.034 | 0.025 | 0.048 | 0.056 | -0.022 | 0.047 | -0.007 | 0.079 | 0.035 | 0.049 |
| rs7248205 | C | T | 0.012 | 0.006 | 0.012 | 0.008 | 0.030 | 0.019 | 0.021 | 0.016 | -0.012 | 0.027 | -0.022 | 0.016 |
| rs111901094 | T | G | 0.032 | 0.009 | 0.038 | 0.012 | 0.011 | 0.027 | 0.024 | 0.023 | 0.092 | 0.039 | 0.064 | 0.023 |
| rs6131281 | C | T | 0.002 | 0.007 | -0.006 | 0.009 | -0.007 | 0.019 | 0.008 | 0.016 | 0.016 | 0.027 | 0.010 | 0.016 |
| rs6141814 | A | C | -0.004 | 0.006 | -0.005 | 0.008 | -0.029 | 0.018 | 0.000 | 0.015 | -0.006 | 0.026 | 0.006 | 0.016 |
| rs4810312 | G | C | 0.009 | 0.006 | 0.014 | 0.008 | 0.007 | 0.018 | 0.006 | 0.015 | -0.005 | 0.026 | -0.017 | 0.015 |
| rs6124476 | T | C | -0.004 | 0.006 | 0.007 | 0.008 | -0.022 | 0.018 | 0.004 | 0.016 | 0.011 | 0.027 | -0.013 | 0.016 |
| rs6031440 | A | T | 0.005 | 0.006 | 0.013 | 0.008 | 0.006 | 0.019 | -0.007 | 0.016 | 0.011 | 0.027 | -0.012 | 0.016 |
| rs55700114 | A | G | 0.002 | 0.006 | 0.007 | 0.009 | -0.035 | 0.019 | 0.019 | 0.016 | -0.018 | 0.027 | 0.008 | 0.016 |
| rs6125906 | A | C | 0.004 | 0.011 | -0.002 | 0.014 | 0.029 | 0.031 | -0.040 | 0.027 | -0.041 | 0.047 | 0.023 | 0.027 |
| rs56103247 | C | T | -0.006 | 0.015 | -0.007 | 0.020 | 0.020 | 0.045 | -0.013 | 0.037 | -0.068 | 0.061 | 0.014 | 0.038 |
| rs10427502 | G | A | -0.012 | 0.006 | -0.013 | 0.008 | -0.003 | 0.018 | -0.021 | 0.016 | -0.067 | 0.027 | -0.014 | 0.016 |
| rs7609 | A | T | -0.003 | 0.007 | -0.001 | 0.010 | -0.014 | 0.021 | -0.024 | 0.018 | -0.029 | 0.031 | -0.010 | 0.019 |
| rs2092829 | G | A | -0.002 | 0.007 | -0.010 | 0.009 | -0.005 | 0.019 | -0.014 | 0.017 | -0.001 | 0.028 | 0.010 | 0.017 |
| rs150399729 | C | G | -0.011 | 0.017 | 0.012 | 0.022 | -0.044 | 0.049 | -0.052 | 0.041 | -0.069 | 0.069 | -0.032 | 0.042 |
| brst breast, EA effect allele, Lum luminal, Neg negative OA other allele, se standard error, SNP single nucleotide polymorphism, 3N triple negative | | | | | | | | | | | | | | |

| **Supplemental Table 4: Summary information on breast cancer risk for the computer use SNPs used for the Mendelian randomisation analyses** | | | | | | | | | | | | | | |
| --- | --- | --- | --- | --- | --- | --- | --- | --- | --- | --- | --- | --- | --- | --- |
| SNP | EA | OA | brst_beta | brst_se | Lum_A_beta | Lum_A_se | Lum_B_beta | Lum_B_se | HER2Neg_beta | HER2Neg_se | HER2_Enriched_beta | HER2_Enriched_se | 3N_beta | 3N_se |
| rs2748985 | C | T | -0.003 | 0.007 | -0.007 | 0.009 | -0.017 | 0.019 | 0.002 | 0.017 | 0.011 | 0.028 | -0.014 | 0.017 |
| rs12145677 | A | G | 0.011 | 0.007 | 0.016 | 0.009 | 0.012 | 0.020 | 0.003 | 0.017 | 0.009 | 0.028 | -0.013 | 0.017 |
| rs7526456 | C | T | -0.005 | 0.006 | -0.005 | 0.009 | -0.012 | 0.019 | -0.002 | 0.016 | 0.039 | 0.028 | -0.014 | 0.016 |
| rs10754920 | C | G | 0.004 | 0.010 | 0.016 | 0.013 | 0.005 | 0.028 | 0.015 | 0.024 | 0.014 | 0.040 | -0.064 | 0.025 |
| rs6744254 | T | C | -0.007 | 0.006 | -0.002 | 0.008 | 0.014 | 0.018 | -0.005 | 0.015 | -0.024 | 0.026 | -0.004 | 0.015 |
| rs11274218 | T | C | -0.014 | 0.006 | -0.008 | 0.008 | -0.030 | 0.018 | -0.003 | 0.016 | -0.006 | 0.027 | -0.021 | 0.016 |
| rs4592851 | T | G | -0.010 | 0.007 | -0.001 | 0.009 | 0.031 | 0.020 | -0.008 | 0.017 | 0.007 | 0.030 | -0.044 | 0.018 |
| rs10208088 | C | T | 0.001 | 0.006 | 0.002 | 0.008 | -0.021 | 0.018 | 0.012 | 0.015 | 0.000 | 0.026 | 0.006 | 0.015 |
| rs11708955 | C | T | -0.023 | 0.007 | -0.018 | 0.009 | -0.016 | 0.019 | -0.025 | 0.016 | -0.062 | 0.028 | -0.053 | 0.016 |
| rs6774533 | T | C | 0.012 | 0.007 | 0.022 | 0.009 | 0.004 | 0.021 | 0.004 | 0.018 | 0.047 | 0.031 | -0.018 | 0.018 |
| rs78082503 | C | G | -0.004 | 0.008 | 0.001 | 0.010 | -0.022 | 0.022 | -0.013 | 0.019 | -0.054 | 0.032 | -0.012 | 0.019 |
| rs6857629 | G | A | 0.009 | 0.007 | 0.014 | 0.009 | 0.003 | 0.021 | 0.023 | 0.018 | 0.005 | 0.030 | 0.015 | 0.018 |
| rs2068625 | C | T | 0.005 | 0.007 | 0.013 | 0.009 | 0.022 | 0.020 | 0.002 | 0.017 | -0.041 | 0.028 | 0.014 | 0.017 |
| rs2220599 | G | C | -0.008 | 0.007 | -0.014 | 0.009 | -0.018 | 0.019 | -0.005 | 0.016 | 0.005 | 0.027 | 0.015 | 0.016 |
| rs11749912 | A | G | 0.002 | 0.006 | -0.001 | 0.008 | 0.010 | 0.018 | 0.019 | 0.015 | -0.015 | 0.026 | 0.006 | 0.015 |
| rs1469249 | G | A | -0.011 | 0.007 | -0.008 | 0.010 | -0.023 | 0.021 | -0.006 | 0.018 | -0.013 | 0.031 | -0.019 | 0.019 |
| rs162894 | T | G | -0.024 | 0.006 | -0.034 | 0.008 | -0.048 | 0.019 | -0.031 | 0.016 | 0.013 | 0.027 | -0.030 | 0.016 |
| rs56158102 | C | T | -0.015 | 0.006 | -0.012 | 0.008 | -0.042 | 0.019 | -0.016 | 0.016 | -0.054 | 0.027 | -0.015 | 0.016 |
| rs9477970 | A | T | 0.002 | 0.008 | -0.001 | 0.010 | 0.001 | 0.023 | -0.017 | 0.020 | -0.007 | 0.033 | 0.002 | 0.020 |
| rs2179517 | C | G | 0.013 | 0.006 | 0.006 | 0.008 | 0.000 | 0.018 | 0.019 | 0.015 | 0.044 | 0.025 | 0.006 | 0.015 |
| rs9372625 | A | G | -0.005 | 0.006 | -0.009 | 0.008 | -0.003 | 0.019 | -0.003 | 0.016 | 0.008 | 0.027 | -0.006 | 0.016 |
| rs17789218 | C | T | 0.013 | 0.008 | 0.025 | 0.011 | 0.002 | 0.023 | -0.030 | 0.020 | -0.068 | 0.034 | -0.014 | 0.020 |
| rs6935828 | T | C | 0.004 | 0.006 | 0.003 | 0.008 | -0.018 | 0.018 | -0.007 | 0.015 | 0.028 | 0.026 | -0.001 | 0.015 |
| rs55772938 | G | A | -0.003 | 0.007 | 0.003 | 0.009 | -0.020 | 0.019 | 0.024 | 0.016 | -0.042 | 0.028 | -0.015 | 0.017 |
| rs12706626 | A | G | 0.009 | 0.006 | 0.016 | 0.008 | -0.012 | 0.018 | 0.015 | 0.015 | -0.006 | 0.026 | -0.004 | 0.016 |
| rs3944151 | A | G | 0.016 | 0.007 | 0.013 | 0.009 | 0.003 | 0.020 | 0.015 | 0.017 | 0.037 | 0.028 | 0.010 | 0.017 |
| rs2345941 | A | G | 0.007 | 0.006 | 0.002 | 0.008 | 0.036 | 0.018 | 0.007 | 0.015 | 0.019 | 0.026 | -0.002 | 0.015 |
| rs13262595 | G | A | 0.001 | 0.006 | -0.005 | 0.008 | 0.016 | 0.018 | -0.013 | 0.015 | -0.043 | 0.026 | 0.005 | 0.015 |
| rs4977839 | A | G | 0.006 | 0.006 | 0.001 | 0.008 | 0.037 | 0.019 | 0.006 | 0.016 | -0.019 | 0.027 | 0.022 | 0.016 |
| rs113851275 | A | G | 0.028 | 0.010 | 0.044 | 0.012 | 0.025 | 0.028 | 0.009 | 0.024 | -0.010 | 0.041 | -0.008 | 0.024 |
| rs7020477 | A | G | -0.005 | 0.008 | -0.011 | 0.010 | -0.012 | 0.022 | 0.007 | 0.019 | 0.007 | 0.033 | 0.037 | 0.020 |
| rs73578186 | C | T | -0.003 | 0.006 | -0.011 | 0.008 | 0.011 | 0.019 | -0.021 | 0.016 | -0.002 | 0.027 | -0.028 | 0.016 |
| rs2734849 | G | A | -0.003 | 0.006 | -0.001 | 0.008 | -0.023 | 0.018 | 0.002 | 0.015 | -0.004 | 0.025 | 0.005 | 0.015 |
| rs1448355 | T | C | 0.001 | 0.006 | 0.001 | 0.008 | -0.011 | 0.018 | 0.002 | 0.015 | -0.007 | 0.026 | 0.006 | 0.016 |
| rs35933007 | A | G | -0.003 | 0.008 | -0.017 | 0.011 | 0.000 | 0.024 | -0.018 | 0.020 | 0.017 | 0.034 | 0.030 | 0.021 |
| rs1319734 | A | G | 0.017 | 0.006 | 0.014 | 0.008 | 0.031 | 0.018 | 0.012 | 0.016 | -0.007 | 0.026 | 0.011 | 0.016 |
| rs206965 | T | C | 0.010 | 0.007 | 0.008 | 0.010 | 0.014 | 0.022 | 0.032 | 0.019 | -0.008 | 0.032 | 0.014 | 0.019 |
| rs12874776 | A | C | -0.001 | 0.010 | -0.007 | 0.013 | -0.036 | 0.030 | -0.007 | 0.026 | 0.005 | 0.044 | -0.018 | 0.026 |
| rs198262 | C | T | 0.002 | 0.015 | 0.011 | 0.020 | 0.041 | 0.044 | -0.020 | 0.038 | -0.019 | 0.065 | -0.010 | 0.039 |
| rs166835 | C | T | -0.013 | 0.006 | -0.023 | 0.008 | -0.038 | 0.018 | 0.002 | 0.015 | 0.001 | 0.026 | -0.015 | 0.015 |
| rs4702 | G | A | 0.029 | 0.006 | 0.016 | 0.008 | 0.057 | 0.018 | 0.071 | 0.015 | -0.002 | 0.026 | 0.007 | 0.015 |
| rs6498759 | T | C | 0.012 | 0.007 | 0.021 | 0.009 | 0.034 | 0.020 | -0.028 | 0.017 | 0.021 | 0.029 | 0.017 | 0.018 |
| rs3730399 | A | G | -0.015 | 0.012 | 0.005 | 0.016 | -0.038 | 0.035 | 0.021 | 0.030 | -0.032 | 0.051 | -0.015 | 0.030 |
| rs4073003 | A | G | -0.007 | 0.009 | -0.005 | 0.012 | -0.015 | 0.027 | -0.025 | 0.023 | -0.047 | 0.039 | -0.006 | 0.023 |
| rs7209653 | T | C | -0.007 | 0.007 | 0.000 | 0.009 | -0.035 | 0.020 | -0.012 | 0.017 | 0.012 | 0.029 | -0.023 | 0.017 |
| rs12603813 | T | C | 0.005 | 0.007 | 0.017 | 0.009 | 0.036 | 0.020 | -0.003 | 0.017 | 0.033 | 0.030 | -0.032 | 0.017 |
| rs66643547 | T | C | -0.020 | 0.007 | -0.021 | 0.009 | -0.005 | 0.019 | -0.024 | 0.017 | -0.061 | 0.029 | -0.019 | 0.017 |
| rs784256 | G | A | -0.005 | 0.008 | 0.006 | 0.010 | -0.020 | 0.023 | 0.009 | 0.020 | -0.015 | 0.033 | -0.034 | 0.020 |
| rs6129084 | T | A | 0.001 | 0.006 | -0.005 | 0.008 | 0.004 | 0.019 | 0.010 | 0.016 | -0.015 | 0.027 | 0.001 | 0.016 |
| rs9608249 | G | A | 0.008 | 0.009 | 0.019 | 0.012 | -0.015 | 0.025 | -0.011 | 0.022 | 0.027 | 0.037 | 0.024 | 0.022 |
| rs136553 | T | C | 0.004 | 0.006 | 0.001 | 0.008 | 0.000 | 0.018 | 0.005 | 0.015 | -0.014 | 0.027 | -0.002 | 0.016 |
| rs7288455 | A | G | 0.001 | 0.006 | 0.005 | 0.008 | -0.018 | 0.019 | 0.004 | 0.016 | -0.025 | 0.027 | -0.007 | 0.016 |
| EA effect allele, ER estrogen receptor, OA other allele, se standard error, SNP single nucleotide polymorphism | | | | | | | | | | | | | | |

| **Supplemental Table 5: Summary information on colorectal cancer risk for the leisure television watching SNPs used for the Mendelian randomisation analyses** | | | | | | | | | | | | |
| --- | --- | --- | --- | --- | --- | --- | --- | --- | --- | --- | --- | --- |
| SNP | ΕΑ | ΟΑ | beta_crc | se_crc | beta_crc_men | se_crc_men | beta_crc_women | se_crc_women | beta_colon | se_colon | beta_re | se_re |
| rs11810109 | A | T | 0.011 | 0.011 | 0.000 | 0.015 | 0.022 | 0.015 | 0.004 | 0.013 | 0.016 | 0.017 |
| rs112632642 | A | G | -0.003 | 0.020 | 0.008 | 0.029 | -0.011 | 0.028 | -0.009 | 0.024 | 0.065 | 0.032 |
| rs6670915 | C | T | 0.005 | 0.010 | 0.011 | 0.014 | 0.001 | 0.014 | 0.000 | 0.012 | 0.012 | 0.015 |
| rs12725114 | G | A | -0.016 | 0.012 | -0.023 | 0.017 | -0.007 | 0.017 | 0.003 | 0.014 | -0.060 | 0.018 |
| rs984409 | A | G | 0.019 | 0.010 | 0.010 | 0.015 | 0.028 | 0.015 | 0.011 | 0.012 | 0.030 | 0.016 |
| rs10890123 | C | T | 0.007 | 0.012 | -0.004 | 0.017 | 0.015 | 0.017 | -0.004 | 0.014 | 0.022 | 0.019 |
| rs71658797 | A | T | 0.005 | 0.016 | 0.002 | 0.023 | 0.009 | 0.023 | 0.009 | 0.019 | 0.009 | 0.025 |
| rs114328297 | T | G | 0.002 | 0.012 | 0.018 | 0.017 | -0.018 | 0.018 | 0.016 | 0.015 | -0.056 | 0.019 |
| rs631130 | A | T | 0.000 | 0.011 | -0.020 | 0.015 | 0.020 | 0.015 | 0.011 | 0.013 | -0.014 | 0.017 |
| rs148648822 | T | G | 0.050 | 0.013 | 0.046 | 0.019 | 0.054 | 0.019 | 0.049 | 0.017 | 0.042 | 0.022 |
| rs17379561 | T | A | -0.016 | 0.014 | -0.004 | 0.020 | -0.024 | 0.020 | -0.016 | 0.017 | -0.016 | 0.022 |
| rs55909997 | A | G | 0.027 | 0.010 | 0.037 | 0.014 | 0.017 | 0.014 | 0.011 | 0.012 | 0.033 | 0.016 |
| rs1278847 | C | A | -0.006 | 0.011 | 0.008 | 0.015 | -0.018 | 0.015 | -0.006 | 0.013 | -0.004 | 0.017 |
| rs141425667 | A | G | 0.018 | 0.021 | 0.020 | 0.029 | 0.020 | 0.030 | 0.037 | 0.025 | 0.000 | 0.032 |
| rs4845364 | G | A | 0.006 | 0.010 | 0.014 | 0.014 | -0.001 | 0.014 | 0.008 | 0.012 | 0.013 | 0.015 |
| rs6673341 | G | T | 0.000 | 0.010 | 0.012 | 0.014 | -0.012 | 0.014 | -0.002 | 0.012 | 0.014 | 0.015 |
| rs72725224 | C | A | 0.075 | 0.038 | 0.044 | 0.053 | 0.098 | 0.055 | 0.056 | 0.045 | 0.106 | 0.057 |
| rs10737620 | T | A | -0.010 | 0.011 | -0.008 | 0.015 | -0.011 | 0.015 | 0.000 | 0.013 | -0.024 | 0.017 |
| rs11896615 | G | A | 0.020 | 0.014 | 0.011 | 0.020 | 0.029 | 0.020 | 0.005 | 0.016 | 0.037 | 0.022 |
| rs72781699 | A | G | -0.009 | 0.012 | -0.008 | 0.017 | -0.008 | 0.017 | 0.012 | 0.014 | -0.042 | 0.019 |
| rs12105701 | T | C | 0.009 | 0.010 | -0.013 | 0.014 | 0.030 | 0.014 | 0.007 | 0.012 | 0.017 | 0.015 |
| rs7564130 | C | T | 0.021 | 0.010 | 0.019 | 0.014 | 0.022 | 0.014 | 0.032 | 0.012 | 0.002 | 0.016 |
| rs11125515 | A | C | 0.005 | 0.011 | 0.001 | 0.015 | 0.008 | 0.015 | 0.006 | 0.013 | 0.019 | 0.017 |
| rs12475168 | T | G | -0.012 | 0.013 | -0.021 | 0.019 | -0.003 | 0.019 | -0.012 | 0.016 | 0.006 | 0.021 |
| rs183566425 | A | T | -0.031 | 0.043 | -0.043 | 0.062 | -0.015 | 0.061 | -0.060 | 0.050 | 0.153 | 0.071 |
| rs10189857 | G | A | 0.022 | 0.010 | 0.022 | 0.014 | 0.021 | 0.014 | 0.020 | 0.012 | 0.019 | 0.015 |
| rs62641636 | A | G | 0.010 | 0.010 | 0.000 | 0.015 | 0.018 | 0.015 | 0.022 | 0.012 | 0.015 | 0.016 |
| rs59210467 | T | C | -0.006 | 0.011 | -0.001 | 0.015 | -0.012 | 0.015 | -0.007 | 0.013 | 0.010 | 0.017 |
| rs11689199 | A | G | 0.025 | 0.010 | 0.039 | 0.014 | 0.012 | 0.014 | 0.020 | 0.012 | 0.034 | 0.016 |
| rs2916251 | T | G | 0.006 | 0.010 | 0.016 | 0.014 | -0.004 | 0.014 | 0.008 | 0.012 | 0.018 | 0.015 |
| rs1451533 | A | G | -0.009 | 0.011 | -0.009 | 0.015 | -0.011 | 0.015 | -0.002 | 0.013 | -0.013 | 0.017 |
| rs2971640 | A | G | 0.013 | 0.010 | 0.014 | 0.014 | 0.013 | 0.014 | 0.007 | 0.012 | 0.022 | 0.015 |
| rs12476388 | C | T | -0.002 | 0.010 | -0.005 | 0.015 | 0.000 | 0.015 | -0.012 | 0.013 | 0.001 | 0.017 |
| rs374722 | G | A | 0.026 | 0.013 | 0.002 | 0.019 | 0.049 | 0.019 | 0.038 | 0.016 | 0.017 | 0.021 |
| rs3754970 | C | T | -0.006 | 0.010 | -0.003 | 0.014 | -0.007 | 0.014 | -0.011 | 0.012 | 0.011 | 0.015 |
| rs1913808 | G | A | 0.014 | 0.010 | 0.025 | 0.015 | 0.005 | 0.015 | 0.003 | 0.013 | 0.022 | 0.017 |
| rs2060199 | T | A | 0.016 | 0.010 | 0.039 | 0.014 | -0.002 | 0.014 | 0.012 | 0.012 | 0.028 | 0.015 |
| rs263771 | A | C | 0.006 | 0.012 | 0.001 | 0.017 | 0.016 | 0.017 | 0.001 | 0.014 | 0.009 | 0.019 |
| rs4577309 | A | G | 0.001 | 0.010 | 0.015 | 0.014 | -0.012 | 0.014 | 0.004 | 0.012 | 0.009 | 0.015 |
| rs34224261 | C | G | -0.001 | 0.010 | 0.002 | 0.014 | -0.005 | 0.014 | 0.002 | 0.012 | 0.000 | 0.015 |
| rs4675246 | T | G | -0.037 | 0.013 | -0.043 | 0.018 | -0.031 | 0.018 | -0.028 | 0.015 | -0.047 | 0.020 |
| rs13029509 | A | G | 0.005 | 0.010 | 0.007 | 0.014 | 0.004 | 0.014 | 0.001 | 0.012 | 0.011 | 0.015 |
| rs10932837 | T | C | 0.003 | 0.010 | -0.013 | 0.014 | 0.020 | 0.014 | 0.005 | 0.012 | 0.016 | 0.015 |
| rs4973576 | A | C | -0.013 | 0.011 | 0.005 | 0.015 | -0.034 | 0.015 | -0.013 | 0.013 | 0.001 | 0.017 |
| rs138256022 | T | C | -0.043 | 0.031 | -0.025 | 0.045 | -0.065 | 0.044 | -0.031 | 0.037 | -0.046 | 0.049 |
| rs9834970 | T | C | 0.013 | 0.010 | 0.002 | 0.014 | 0.024 | 0.014 | 0.010 | 0.012 | 0.017 | 0.015 |
| rs1352890 | T | C | -0.006 | 0.010 | 0.007 | 0.014 | -0.014 | 0.014 | -0.016 | 0.012 | 0.029 | 0.015 |
| rs3796386 | A | G | 0.015 | 0.010 | 0.014 | 0.014 | 0.018 | 0.014 | 0.012 | 0.012 | 0.040 | 0.015 |
| rs182355396 | A | C | -0.002 | 0.061 | 0.094 | 0.087 | -0.098 | 0.084 | 0.047 | 0.072 | -0.052 | 0.097 |
| rs11917871 | C | T | -0.012 | 0.010 | -0.013 | 0.014 | -0.006 | 0.014 | -0.012 | 0.012 | 0.005 | 0.016 |
| rs11130793 | C | T | 0.008 | 0.010 | 0.018 | 0.014 | -0.003 | 0.014 | -0.004 | 0.012 | 0.011 | 0.016 |
| rs11714337 | G | A | -0.021 | 0.010 | -0.010 | 0.014 | -0.029 | 0.014 | -0.029 | 0.012 | -0.011 | 0.015 |
| rs12496084 | G | A | 0.007 | 0.011 | 0.010 | 0.016 | 0.006 | 0.016 | 0.003 | 0.014 | 0.006 | 0.018 |
| rs6797840 | C | A | -0.009 | 0.010 | 0.004 | 0.014 | -0.018 | 0.014 | -0.016 | 0.012 | -0.015 | 0.015 |
| rs56398417 | C | T | 0.015 | 0.011 | 0.028 | 0.015 | 0.001 | 0.015 | 0.011 | 0.013 | 0.013 | 0.017 |
| rs2034768 | A | G | -0.013 | 0.010 | -0.017 | 0.014 | -0.007 | 0.014 | -0.014 | 0.012 | -0.007 | 0.015 |
| rs12632778 | G | T | -0.029 | 0.021 | -0.045 | 0.030 | -0.021 | 0.030 | -0.035 | 0.028 | 0.026 | 0.036 |
| rs9867121 | C | A | 0.020 | 0.012 | 0.041 | 0.017 | -0.004 | 0.017 | 0.018 | 0.015 | 0.014 | 0.019 |
| rs12491503 | A | G | 0.001 | 0.010 | 0.011 | 0.014 | -0.008 | 0.014 | 0.005 | 0.012 | 0.006 | 0.016 |
| rs114600294 | C | G | -0.013 | 0.014 | -0.022 | 0.020 | -0.005 | 0.020 | -0.021 | 0.017 | -0.007 | 0.022 |
| rs66852340 | T | C | 0.004 | 0.011 | 0.013 | 0.016 | -0.002 | 0.016 | 0.000 | 0.014 | 0.030 | 0.018 |
| rs7700107 | C | A | 0.003 | 0.014 | 0.025 | 0.019 | -0.017 | 0.019 | -0.018 | 0.017 | 0.011 | 0.022 |
| rs34811474 | G | A | 0.022 | 0.013 | 0.009 | 0.019 | 0.035 | 0.019 | 0.038 | 0.016 | 0.016 | 0.021 |
| rs77215114 | A | T | 0.011 | 0.019 | -0.017 | 0.027 | 0.036 | 0.027 | 0.031 | 0.024 | 0.017 | 0.032 |
| rs28971796 | A | G | 0.010 | 0.012 | 0.004 | 0.017 | 0.018 | 0.017 | 0.003 | 0.015 | 0.037 | 0.019 |
| rs6850494 | C | A | 0.011 | 0.010 | 0.019 | 0.014 | 0.002 | 0.014 | 0.029 | 0.012 | 0.008 | 0.016 |
| rs13107325 | T | C | -0.008 | 0.019 | 0.009 | 0.026 | -0.028 | 0.027 | -0.012 | 0.022 | 0.006 | 0.029 |
| rs62332760 | T | G | 0.031 | 0.010 | 0.032 | 0.014 | 0.029 | 0.014 | 0.031 | 0.012 | 0.026 | 0.016 |
| rs7693703 | G | A | 0.022 | 0.017 | -0.004 | 0.024 | 0.050 | 0.024 | 0.018 | 0.020 | 0.070 | 0.027 |
| rs4334769 | G | T | -0.015 | 0.010 | -0.010 | 0.014 | -0.016 | 0.014 | -0.023 | 0.012 | -0.015 | 0.015 |
| rs6825241 | A | C | 0.008 | 0.010 | 0.003 | 0.014 | 0.012 | 0.014 | 0.005 | 0.012 | 0.016 | 0.015 |
| rs7693082 | G | C | 0.017 | 0.011 | 0.012 | 0.015 | 0.020 | 0.015 | 0.026 | 0.013 | 0.020 | 0.017 |
| rs10940659 | G | A | 0.011 | 0.010 | 0.027 | 0.014 | -0.003 | 0.014 | 0.011 | 0.012 | 0.020 | 0.015 |
| rs2409838 | A | T | 0.002 | 0.010 | -0.007 | 0.014 | 0.010 | 0.014 | 0.006 | 0.012 | -0.010 | 0.015 |
| rs262890 | G | A | -0.031 | 0.011 | -0.036 | 0.015 | -0.028 | 0.015 | -0.028 | 0.013 | -0.043 | 0.017 |
| rs7716447 | G | A | 0.016 | 0.010 | 0.018 | 0.014 | 0.009 | 0.014 | 0.007 | 0.012 | 0.021 | 0.016 |
| rs1031423 | C | T | -0.020 | 0.012 | -0.023 | 0.017 | -0.017 | 0.017 | -0.022 | 0.014 | -0.017 | 0.019 |
| rs57585211 | G | T | 0.003 | 0.013 | 0.020 | 0.018 | -0.012 | 0.018 | 0.008 | 0.016 | 0.009 | 0.021 |
| rs9326862 | A | T | 0.001 | 0.010 | -0.011 | 0.014 | 0.014 | 0.014 | 0.006 | 0.012 | -0.010 | 0.015 |
| rs10041724 | T | C | 0.015 | 0.012 | 0.021 | 0.017 | 0.010 | 0.017 | 0.016 | 0.014 | 0.012 | 0.019 |
| rs10057545 | A | G | 0.013 | 0.010 | 0.021 | 0.014 | 0.005 | 0.014 | 0.013 | 0.012 | 0.003 | 0.015 |
| rs62379379 | T | G | 0.032 | 0.019 | 0.025 | 0.027 | 0.038 | 0.026 | 0.029 | 0.023 | 0.022 | 0.031 |
| rs10054327 | G | A | -0.002 | 0.010 | 0.003 | 0.014 | -0.005 | 0.014 | -0.013 | 0.012 | -0.002 | 0.016 |
| rs42210 | C | G | 0.007 | 0.011 | 0.016 | 0.015 | -0.004 | 0.015 | -0.004 | 0.013 | 0.024 | 0.017 |
| rs2336895 | G | A | 0.008 | 0.010 | 0.020 | 0.014 | -0.003 | 0.014 | 0.016 | 0.012 | 0.020 | 0.016 |
| rs72828890 | C | T | 0.012 | 0.016 | 0.026 | 0.023 | -0.002 | 0.023 | 0.017 | 0.019 | 0.003 | 0.025 |
| rs255292 | A | C | 0.006 | 0.010 | 0.015 | 0.014 | -0.002 | 0.014 | 0.012 | 0.012 | -0.004 | 0.015 |
| rs72834698 | G | A | 0.044 | 0.014 | 0.060 | 0.020 | 0.031 | 0.020 | 0.059 | 0.017 | 0.069 | 0.022 |
| rs3135044 | T | C | 0.014 | 0.010 | 0.018 | 0.014 | 0.009 | 0.014 | 0.020 | 0.012 | 0.011 | 0.016 |
| rs10947452 | T | C | 0.019 | 0.010 | 0.034 | 0.014 | 0.005 | 0.014 | 0.011 | 0.012 | 0.042 | 0.016 |
| rs9471333 | C | T | 0.010 | 0.010 | 0.020 | 0.014 | 0.001 | 0.014 | 0.007 | 0.012 | 0.018 | 0.015 |
| rs4523073 | G | A | -0.006 | 0.010 | -0.002 | 0.014 | -0.009 | 0.014 | -0.002 | 0.012 | -0.022 | 0.016 |
| rs6905544 | G | A | 0.029 | 0.010 | 0.037 | 0.014 | 0.023 | 0.014 | 0.024 | 0.012 | 0.035 | 0.015 |
| rs17789218 | T | C | 0.004 | 0.012 | 0.016 | 0.016 | -0.006 | 0.016 | -0.006 | 0.014 | 0.011 | 0.018 |
| rs78394231 | C | T | 0.024 | 0.016 | 0.010 | 0.023 | 0.040 | 0.023 | 0.034 | 0.019 | 0.056 | 0.025 |
| rs2184364 | A | G | 0.023 | 0.012 | 0.025 | 0.017 | 0.022 | 0.017 | 0.024 | 0.015 | 0.017 | 0.019 |
| rs9718104 | G | T | 0.002 | 0.022 | -0.002 | 0.032 | 0.013 | 0.031 | -0.009 | 0.027 | -0.005 | 0.036 |
| rs17568389 | T | A | 0.001 | 0.010 | 0.012 | 0.014 | -0.012 | 0.014 | 0.000 | 0.012 | 0.012 | 0.015 |
| rs10234444 | G | A | -0.017 | 0.013 | -0.018 | 0.018 | -0.018 | 0.018 | -0.023 | 0.016 | -0.004 | 0.021 |
| rs6973656 | G | A | -0.029 | 0.010 | -0.025 | 0.014 | -0.031 | 0.014 | -0.028 | 0.012 | -0.036 | 0.015 |
| rs973734 | C | A | 0.002 | 0.014 | -0.022 | 0.020 | 0.031 | 0.020 | 0.013 | 0.016 | -0.009 | 0.021 |
| rs3137 | G | A | 0.013 | 0.013 | -0.005 | 0.018 | 0.034 | 0.018 | 0.013 | 0.016 | -0.003 | 0.021 |
| rs2694151 | C | T | 0.011 | 0.020 | 0.008 | 0.028 | 0.016 | 0.028 | 0.003 | 0.023 | 0.047 | 0.030 |
| rs62471080 | C | G | 0.005 | 0.010 | 0.007 | 0.014 | 0.004 | 0.014 | -0.010 | 0.012 | 0.012 | 0.015 |
| rs7788008 | G | A | 0.001 | 0.010 | 0.001 | 0.014 | 0.002 | 0.014 | 0.005 | 0.012 | -0.027 | 0.015 |
| rs10246289 | A | G | 0.041 | 0.015 | 0.039 | 0.022 | 0.042 | 0.022 | 0.033 | 0.019 | 0.033 | 0.024 |
| rs11763734 | C | A | 0.001 | 0.010 | -0.001 | 0.014 | 0.005 | 0.014 | 0.002 | 0.012 | -0.005 | 0.015 |
| rs7834121 | T | G | -0.007 | 0.010 | -0.015 | 0.015 | 0.000 | 0.015 | 0.007 | 0.012 | -0.028 | 0.016 |
| rs1421334 | A | C | 0.017 | 0.010 | 0.017 | 0.014 | 0.018 | 0.014 | 0.022 | 0.012 | -0.005 | 0.015 |
| rs6996198 | T | C | 0.016 | 0.013 | 0.009 | 0.018 | 0.025 | 0.018 | 0.011 | 0.016 | 0.017 | 0.020 |
| rs6472942 | C | T | 0.000 | 0.010 | 0.006 | 0.014 | -0.004 | 0.014 | 0.001 | 0.012 | -0.015 | 0.015 |
| rs11985394 | T | A | 0.010 | 0.010 | 0.017 | 0.014 | -0.001 | 0.014 | 0.020 | 0.012 | 0.000 | 0.015 |
| rs72671494 | C | T | 0.006 | 0.015 | -0.007 | 0.022 | 0.018 | 0.022 | 0.011 | 0.018 | -0.011 | 0.024 |
| rs12541615 | C | T | 0.003 | 0.012 | -0.001 | 0.017 | 0.008 | 0.018 | 0.013 | 0.015 | -0.006 | 0.019 |
| rs2717559 | A | G | 0.000 | 0.010 | -0.018 | 0.014 | 0.018 | 0.014 | -0.006 | 0.012 | 0.003 | 0.015 |
| rs2616830 | G | A | 0.013 | 0.010 | 0.014 | 0.014 | 0.008 | 0.014 | 0.016 | 0.012 | 0.031 | 0.015 |
| rs2291681 | G | T | -0.003 | 0.010 | -0.004 | 0.014 | -0.003 | 0.014 | -0.002 | 0.012 | -0.010 | 0.015 |
| rs34864022 | G | A | -0.013 | 0.021 | -0.033 | 0.030 | 0.010 | 0.030 | -0.015 | 0.025 | 0.026 | 0.033 |
| rs12554512 | T | C | -0.002 | 0.010 | -0.001 | 0.014 | -0.002 | 0.014 | 0.011 | 0.012 | -0.005 | 0.016 |
| rs2791574 | G | T | 0.020 | 0.010 | 0.017 | 0.014 | 0.019 | 0.014 | 0.021 | 0.012 | 0.014 | 0.016 |
| rs870151 | A | T | -0.010 | 0.010 | -0.005 | 0.014 | -0.018 | 0.014 | -0.011 | 0.012 | -0.005 | 0.015 |
| rs2787374 | T | C | 0.012 | 0.010 | -0.002 | 0.014 | 0.025 | 0.014 | 0.005 | 0.012 | 0.028 | 0.015 |
| rs10739499 | C | G | -0.006 | 0.010 | 0.002 | 0.014 | -0.016 | 0.015 | 0.001 | 0.012 | 0.024 | 0.016 |
| rs7043521 | A | T | -0.021 | 0.010 | -0.014 | 0.014 | -0.027 | 0.014 | -0.021 | 0.012 | -0.043 | 0.015 |
| rs12342024 | T | G | 0.033 | 0.015 | 0.045 | 0.022 | 0.019 | 0.022 | 0.048 | 0.018 | 0.015 | 0.024 |
| rs4382592 | T | G | 0.009 | 0.010 | 0.013 | 0.015 | 0.005 | 0.015 | 0.006 | 0.013 | 0.000 | 0.016 |
| rs2073869 | C | T | -0.011 | 0.013 | -0.002 | 0.018 | -0.018 | 0.018 | -0.018 | 0.015 | -0.019 | 0.020 |
| rs1623025 | C | G | -0.018 | 0.010 | -0.019 | 0.014 | -0.017 | 0.014 | -0.019 | 0.012 | -0.006 | 0.015 |
| rs1243182 | T | C | 0.029 | 0.011 | 0.039 | 0.015 | 0.020 | 0.015 | 0.025 | 0.013 | 0.054 | 0.016 |
| rs2045147 | A | G | 0.009 | 0.010 | 0.018 | 0.014 | -0.001 | 0.014 | 0.006 | 0.012 | 0.019 | 0.015 |
| rs10994943 | T | G | 0.007 | 0.010 | 0.017 | 0.014 | -0.006 | 0.014 | 0.000 | 0.012 | 0.014 | 0.015 |
| rs11201422 | T | C | 0.019 | 0.011 | 0.005 | 0.015 | 0.036 | 0.015 | 0.022 | 0.012 | 0.027 | 0.016 |
| rs10786658 | T | A | -0.003 | 0.010 | -0.011 | 0.014 | 0.007 | 0.014 | -0.002 | 0.012 | 0.001 | 0.015 |
| rs7089973 | A | C | 0.021 | 0.010 | 0.022 | 0.015 | 0.020 | 0.014 | 0.028 | 0.012 | 0.036 | 0.016 |
| rs11245482 | C | T | 0.004 | 0.010 | -0.006 | 0.014 | 0.016 | 0.014 | 0.002 | 0.012 | 0.008 | 0.015 |
| rs17727474 | C | T | 0.001 | 0.013 | -0.001 | 0.019 | 0.005 | 0.019 | -0.009 | 0.016 | 0.010 | 0.021 |
| rs12289262 | T | C | 0.028 | 0.011 | 0.029 | 0.016 | 0.029 | 0.016 | 0.030 | 0.014 | 0.010 | 0.018 |
| rs801733 | A | C | 0.027 | 0.010 | 0.028 | 0.014 | 0.026 | 0.014 | 0.023 | 0.012 | 0.048 | 0.016 |
| rs116175338 | T | G | 0.001 | 0.011 | -0.011 | 0.016 | 0.011 | 0.016 | 0.002 | 0.014 | -0.020 | 0.018 |
| rs12272012 | C | G | -0.031 | 0.024 | 0.000 | 0.034 | -0.062 | 0.033 | -0.036 | 0.028 | -0.064 | 0.037 |
| rs11020045 | C | A | -0.007 | 0.010 | -0.021 | 0.015 | 0.006 | 0.015 | -0.018 | 0.012 | 0.003 | 0.016 |
| rs17207890 | G | A | 0.000 | 0.010 | 0.012 | 0.015 | -0.010 | 0.015 | 0.016 | 0.012 | -0.021 | 0.016 |
| rs10791546 | C | T | 0.018 | 0.010 | 0.023 | 0.014 | 0.020 | 0.014 | 0.025 | 0.012 | 0.002 | 0.015 |
| rs328277 | C | A | 0.000 | 0.010 | -0.004 | 0.014 | 0.003 | 0.014 | 0.005 | 0.012 | -0.017 | 0.016 |
| rs648044 | G | A | -0.002 | 0.011 | 0.003 | 0.015 | -0.008 | 0.015 | -0.002 | 0.013 | 0.016 | 0.017 |
| rs11218575 | C | T | 0.001 | 0.010 | 0.013 | 0.014 | -0.010 | 0.014 | 0.001 | 0.012 | -0.001 | 0.015 |
| rs74802478 | G | A | -0.028 | 0.013 | -0.029 | 0.018 | -0.029 | 0.018 | -0.017 | 0.015 | -0.052 | 0.020 |
| rs4937842 | C | G | 0.012 | 0.010 | 0.018 | 0.014 | 0.008 | 0.014 | 0.018 | 0.012 | 0.023 | 0.016 |
| rs10772643 | C | T | 0.027 | 0.016 | 0.041 | 0.022 | 0.014 | 0.022 | 0.024 | 0.019 | 0.056 | 0.024 |
| rs74996610 | G | C | -0.013 | 0.024 | -0.004 | 0.034 | -0.020 | 0.034 | 0.001 | 0.030 | -0.019 | 0.039 |
| rs10771746 | T | C | -0.006 | 0.010 | 0.008 | 0.015 | -0.017 | 0.015 | -0.001 | 0.013 | -0.031 | 0.017 |
| rs10876864 | A | G | 0.010 | 0.010 | 0.005 | 0.014 | 0.018 | 0.014 | 0.029 | 0.012 | -0.021 | 0.015 |
| rs8756 | A | C | 0.009 | 0.010 | 0.008 | 0.014 | 0.009 | 0.014 | 0.005 | 0.012 | 0.016 | 0.015 |
| rs11179838 | C | G | 0.006 | 0.010 | -0.012 | 0.014 | 0.023 | 0.014 | 0.019 | 0.012 | 0.006 | 0.016 |
| rs2164744 | C | T | -0.014 | 0.010 | -0.013 | 0.014 | -0.015 | 0.014 | -0.007 | 0.012 | -0.027 | 0.015 |
| rs2173650 | G | T | -0.006 | 0.013 | 0.016 | 0.019 | -0.028 | 0.019 | -0.002 | 0.016 | 0.010 | 0.021 |
| rs1790135 | T | C | 0.030 | 0.011 | 0.022 | 0.016 | 0.037 | 0.016 | 0.039 | 0.013 | 0.034 | 0.017 |
| rs9563168 | G | A | 0.006 | 0.012 | -0.004 | 0.017 | 0.016 | 0.017 | 0.010 | 0.014 | -0.013 | 0.018 |
| rs9569734 | A | G | 0.010 | 0.013 | 0.024 | 0.019 | -0.011 | 0.019 | 0.012 | 0.016 | 0.037 | 0.021 |
| rs9528023 | C | T | 0.005 | 0.011 | 0.013 | 0.016 | -0.005 | 0.016 | 0.018 | 0.013 | 0.000 | 0.017 |
| rs56858768 | A | G | 0.010 | 0.011 | 0.006 | 0.015 | 0.015 | 0.015 | 0.012 | 0.013 | 0.019 | 0.017 |
| rs553397860 | A | G | 0.004 | 0.014 | 0.004 | 0.019 | 0.008 | 0.019 | 0.002 | 0.016 | 0.020 | 0.021 |
| rs7991062 | G | C | 0.013 | 0.010 | 0.011 | 0.014 | 0.015 | 0.014 | 0.012 | 0.012 | 0.017 | 0.016 |
| rs11629120 | C | T | 0.027 | 0.012 | 0.027 | 0.014 | 0.044 | 0.014 | 0.038 | 0.012 | 0.037 | 0.015 |
| rs405797 | A | T | 0.014 | 0.013 | 0.009 | 0.016 | 0.023 | 0.016 | 0.017 | 0.013 | 0.006 | 0.017 |
| rs1022785 | G | A | -0.003 | 0.016 | 0.027 | 0.019 | -0.024 | 0.019 | -0.005 | 0.016 | 0.035 | 0.021 |
| rs10145592 | G | C | 0.023 | 0.012 | 0.017 | 0.014 | 0.014 | 0.014 | 0.019 | 0.012 | 0.029 | 0.015 |
| rs7157001 | G | A | 0.007 | 0.014 | 0.005 | 0.017 | 0.013 | 0.017 | 0.003 | 0.014 | 0.051 | 0.019 |
| rs8043253 | T | C | -0.017 | 0.010 | -0.013 | 0.014 | -0.019 | 0.014 | -0.028 | 0.012 | 0.002 | 0.015 |
| rs2460 | A | G | 0.015 | 0.011 | -0.004 | 0.016 | 0.032 | 0.016 | 0.021 | 0.013 | -0.015 | 0.018 |
| rs4775373 | T | C | 0.006 | 0.010 | 0.002 | 0.014 | 0.009 | 0.014 | 0.007 | 0.012 | 0.017 | 0.016 |
| rs61331678 | G | C | -0.011 | 0.010 | 0.004 | 0.014 | -0.025 | 0.014 | -0.015 | 0.012 | -0.011 | 0.015 |
| rs35574015 | C | T | -0.020 | 0.010 | -0.028 | 0.015 | -0.011 | 0.015 | -0.018 | 0.013 | 0.001 | 0.016 |
| rs7189927 | T | C | 0.009 | 0.010 | 0.006 | 0.015 | 0.012 | 0.015 | 0.016 | 0.012 | 0.009 | 0.016 |
| rs749671 | G | A | 0.023 | 0.010 | 0.018 | 0.014 | 0.028 | 0.014 | 0.023 | 0.012 | 0.015 | 0.016 |
| rs7184800 | G | A | 0.005 | 0.011 | -0.003 | 0.015 | 0.011 | 0.015 | 0.003 | 0.013 | 0.024 | 0.016 |
| rs2447098 | A | C | 0.011 | 0.010 | 0.034 | 0.014 | -0.009 | 0.014 | 0.010 | 0.012 | 0.003 | 0.015 |
| rs11654952 | G | T | -0.025 | 0.014 | -0.005 | 0.019 | -0.047 | 0.020 | -0.018 | 0.017 | -0.001 | 0.022 |
| rs9902312 | T | C | -0.001 | 0.010 | -0.005 | 0.015 | 0.004 | 0.015 | 0.000 | 0.012 | 0.010 | 0.016 |
| rs11657730 | C | T | 0.010 | 0.011 | 0.007 | 0.015 | 0.012 | 0.015 | 0.010 | 0.012 | 0.016 | 0.016 |
| rs303753 | A | G | 0.013 | 0.010 | 0.029 | 0.015 | -0.001 | 0.015 | 0.008 | 0.012 | 0.022 | 0.016 |
| rs11081851 | A | G | 0.001 | 0.010 | 0.007 | 0.014 | -0.004 | 0.014 | 0.005 | 0.012 | -0.012 | 0.015 |
| rs9964724 | C | T | 0.009 | 0.010 | 0.014 | 0.015 | 0.004 | 0.015 | 0.011 | 0.012 | 0.012 | 0.016 |
| rs1156541 | C | T | 0.017 | 0.011 | 0.016 | 0.016 | 0.019 | 0.016 | 0.015 | 0.014 | 0.043 | 0.018 |
| rs148544378 | C | T | 0.046 | 0.047 | 0.153 | 0.067 | -0.060 | 0.067 | 0.045 | 0.056 | 0.068 | 0.073 |
| rs17512836 | T | C | -0.014 | 0.028 | -0.018 | 0.040 | -0.009 | 0.040 | 0.019 | 0.033 | -0.075 | 0.043 |
| rs7248205 | C | T | -0.029 | 0.010 | -0.046 | 0.014 | -0.010 | 0.014 | -0.021 | 0.012 | -0.023 | 0.016 |
| rs111901094 | T | G | 0.011 | 0.013 | -0.022 | 0.018 | 0.044 | 0.018 | -0.003 | 0.016 | 0.022 | 0.020 |
| rs6131281 | C | T | 0.017 | 0.010 | 0.021 | 0.014 | 0.011 | 0.014 | 0.020 | 0.012 | 0.014 | 0.015 |
| rs6141814 | A | C | -0.013 | 0.010 | -0.020 | 0.014 | -0.004 | 0.014 | -0.015 | 0.012 | -0.003 | 0.016 |
| rs4810312 | G | C | -0.007 | 0.010 | -0.010 | 0.014 | -0.003 | 0.014 | -0.010 | 0.012 | 0.008 | 0.015 |
| rs6124476 | T | C | -0.005 | 0.010 | -0.015 | 0.014 | 0.003 | 0.014 | -0.013 | 0.012 | 0.024 | 0.015 |
| rs6031440 | A | T | -0.013 | 0.010 | -0.018 | 0.014 | -0.009 | 0.014 | -0.015 | 0.012 | 0.005 | 0.016 |
| rs55700114 | A | G | 0.001 | 0.011 | 0.006 | 0.015 | -0.006 | 0.015 | 0.002 | 0.013 | 0.006 | 0.016 |
| rs6125906 | A | C | -0.019 | 0.017 | -0.021 | 0.024 | -0.019 | 0.024 | -0.016 | 0.020 | -0.009 | 0.026 |
| rs56103247 | C | T | -0.077 | 0.024 | -0.075 | 0.035 | -0.080 | 0.034 | -0.074 | 0.029 | -0.074 | 0.038 |
| rs10427502 | G | A | 0.001 | 0.010 | 0.024 | 0.014 | -0.017 | 0.014 | -0.006 | 0.012 | 0.000 | 0.016 |
| rs7609 | A | T | 0.001 | 0.011 | -0.016 | 0.016 | 0.014 | 0.016 | 0.003 | 0.014 | -0.014 | 0.018 |
| rs2092829 | G | A | -0.023 | 0.011 | -0.027 | 0.015 | -0.016 | 0.015 | -0.026 | 0.013 | -0.029 | 0.017 |
| crc colorectal, EA effect allele, OA other allele, re rectal, se standard error, SNP single nucleotide polymorphism | | | | | | | | | | | | |

| **Supplemental Table 6: Summary information on colorectal cancer risk for the leisure computer use SNPs used for the Mendelian randomisation analyses** | | | | | | | | | | | | |
| --- | --- | --- | --- | --- | --- | --- | --- | --- | --- | --- | --- | --- |
| SNP | ΕΑ | ΟΑ | beta_crc | se_crc | beta_crc_men | se_crc_men | beta_crc_women | se_crc_women | beta_colon | se_colon | beta_re | se_re |
| rs2748985 | C | T | -0.006 | 0.010 | -0.017 | 0.014 | 0.003 | 0.014 | -0.016 | 0.012 | 0.003 | 0.015 |
| rs12145677 | A | G | 0.006 | 0.011 | -0.012 | 0.015 | 0.023 | 0.015 | 0.007 | 0.013 | 0.004 | 0.017 |
| rs7526456 | C | T | 0.005 | 0.010 | 0.001 | 0.014 | 0.009 | 0.014 | 0.004 | 0.012 | 0.012 | 0.015 |
| rs10754920 | C | G | 0.002 | 0.014 | -0.012 | 0.020 | 0.011 | 0.020 | 0.007 | 0.018 | 0.011 | 0.023 |
| rs6744254 | T | C | -0.014 | 0.010 | -0.041 | 0.014 | 0.012 | 0.014 | -0.020 | 0.012 | -0.021 | 0.015 |
| rs11274218 | T | C | 0.007 | 0.010 | -0.005 | 0.014 | 0.016 | 0.014 | 0.009 | 0.012 | -0.001 | 0.015 |
| rs4592851 | T | G | -0.015 | 0.011 | -0.020 | 0.016 | -0.010 | 0.016 | -0.018 | 0.013 | -0.009 | 0.017 |
| rs10208088 | C | T | 0.002 | 0.010 | 0.007 | 0.014 | -0.006 | 0.014 | 0.003 | 0.012 | -0.019 | 0.015 |
| rs11708955 | C | T | 0.001 | 0.010 | -0.007 | 0.015 | 0.006 | 0.015 | 0.007 | 0.012 | -0.029 | 0.016 |
| rs6774533 | T | C | 0.004 | 0.011 | 0.008 | 0.015 | 0.001 | 0.015 | 0.009 | 0.013 | -0.009 | 0.017 |
| rs78082503 | C | G | 0.007 | 0.011 | 0.020 | 0.016 | -0.004 | 0.016 | 0.002 | 0.014 | -0.033 | 0.018 |
| rs6857629 | G | A | 0.021 | 0.011 | 0.034 | 0.016 | 0.012 | 0.016 | 0.014 | 0.013 | 0.026 | 0.017 |
| rs2068625 | C | T | -0.017 | 0.011 | -0.011 | 0.015 | -0.020 | 0.015 | -0.025 | 0.013 | -0.020 | 0.017 |
| rs2220599 | G | C | 0.003 | 0.010 | 0.010 | 0.014 | 0.000 | 0.014 | -0.001 | 0.012 | 0.002 | 0.016 |
| rs11749912 | A | G | 0.015 | 0.010 | 0.010 | 0.014 | 0.023 | 0.014 | 0.019 | 0.012 | -0.001 | 0.015 |
| rs1469249 | G | A | 0.009 | 0.012 | 0.017 | 0.017 | 0.005 | 0.017 | -0.001 | 0.014 | 0.038 | 0.018 |
| rs162894 | T | G | -0.012 | 0.010 | -0.017 | 0.014 | -0.007 | 0.014 | -0.018 | 0.012 | -0.012 | 0.016 |
| rs56158102 | C | T | -0.006 | 0.010 | 0.003 | 0.014 | -0.014 | 0.014 | -0.001 | 0.012 | -0.001 | 0.016 |
| rs9477970 | A | T | 0.017 | 0.012 | 0.032 | 0.018 | 0.004 | 0.018 | 0.016 | 0.015 | 0.005 | 0.020 |
| rs2179517 | C | G | -0.020 | 0.010 | -0.024 | 0.014 | -0.014 | 0.014 | -0.030 | 0.012 | -0.016 | 0.015 |
| rs9372625 | A | G | -0.031 | 0.010 | -0.033 | 0.014 | -0.030 | 0.014 | -0.026 | 0.012 | -0.041 | 0.016 |
| rs17789218 | C | T | -0.004 | 0.012 | -0.016 | 0.016 | 0.006 | 0.016 | 0.006 | 0.014 | -0.011 | 0.018 |
| rs6935828 | T | C | 0.002 | 0.010 | -0.005 | 0.014 | 0.004 | 0.014 | -0.002 | 0.012 | -0.022 | 0.015 |
| rs55772938 | G | A | 0.008 | 0.011 | -0.001 | 0.015 | 0.017 | 0.015 | 0.001 | 0.013 | 0.006 | 0.016 |
| rs12706626 | A | G | 0.007 | 0.010 | 0.029 | 0.014 | -0.016 | 0.014 | 0.007 | 0.012 | -0.003 | 0.016 |
| rs3944151 | A | G | 0.021 | 0.011 | 0.020 | 0.015 | 0.019 | 0.015 | 0.029 | 0.013 | 0.016 | 0.017 |
| rs2345941 | A | G | -0.023 | 0.010 | -0.013 | 0.014 | -0.033 | 0.014 | -0.025 | 0.012 | -0.012 | 0.015 |
| rs13262595 | G | A | 0.001 | 0.010 | -0.004 | 0.014 | 0.008 | 0.014 | 0.007 | 0.012 | -0.012 | 0.015 |
| rs4977839 | A | G | 0.004 | 0.010 | 0.002 | 0.014 | 0.005 | 0.014 | -0.010 | 0.012 | 0.006 | 0.016 |
| rs113851275 | A | G | -0.022 | 0.015 | -0.001 | 0.022 | -0.043 | 0.022 | -0.010 | 0.019 | 0.001 | 0.024 |
| rs7020477 | A | G | 0.016 | 0.012 | -0.004 | 0.017 | 0.033 | 0.017 | 0.020 | 0.014 | 0.011 | 0.018 |
| rs73578186 | C | T | -0.015 | 0.010 | -0.022 | 0.014 | -0.006 | 0.015 | -0.018 | 0.012 | -0.003 | 0.016 |
| rs2734849 | G | A | 0.003 | 0.010 | -0.004 | 0.014 | 0.010 | 0.014 | 0.009 | 0.012 | -0.012 | 0.015 |
| rs1448355 | T | C | -0.012 | 0.010 | 0.008 | 0.014 | -0.030 | 0.014 | -0.004 | 0.012 | -0.013 | 0.016 |
| rs35933007 | A | G | 0.009 | 0.013 | 0.005 | 0.019 | 0.017 | 0.019 | 0.011 | 0.016 | 0.002 | 0.021 |
| rs1319734 | A | G | 0.014 | 0.010 | 0.008 | 0.014 | 0.021 | 0.014 | 0.016 | 0.012 | 0.002 | 0.016 |
| rs206965 | T | C | -0.008 | 0.012 | -0.014 | 0.017 | 0.001 | 0.017 | -0.001 | 0.014 | -0.029 | 0.019 |
| rs12874776 | A | C | 0.010 | 0.016 | 0.008 | 0.023 | 0.009 | 0.022 | 0.008 | 0.019 | 0.012 | 0.025 |
| rs198262 | C | T | -0.023 | 0.030 | 0.005 | 0.035 | -0.003 | 0.034 | 0.015 | 0.029 | -0.038 | 0.038 |
| rs166835 | C | T | -0.007 | 0.010 | -0.013 | 0.014 | 0.002 | 0.014 | -0.001 | 0.012 | -0.012 | 0.015 |
| rs4702 | G | A | 0.018 | 0.010 | 0.022 | 0.014 | 0.014 | 0.014 | 0.020 | 0.012 | 0.013 | 0.015 |
| rs6498759 | T | C | -0.003 | 0.010 | 0.001 | 0.014 | -0.009 | 0.014 | -0.004 | 0.012 | 0.000 | 0.016 |
| rs3730399 | A | G | -0.054 | 0.020 | -0.003 | 0.028 | -0.104 | 0.029 | -0.075 | 0.024 | -0.048 | 0.031 |
| rs4073003 | A | G | 0.009 | 0.015 | 0.016 | 0.021 | -0.001 | 0.021 | -0.002 | 0.018 | 0.013 | 0.023 |
| rs7209653 | T | C | -0.006 | 0.011 | -0.010 | 0.015 | 0.001 | 0.015 | -0.016 | 0.013 | 0.011 | 0.017 |
| rs12603813 | T | C | -0.003 | 0.011 | -0.024 | 0.016 | 0.018 | 0.016 | 0.012 | 0.013 | -0.019 | 0.017 |
| rs66643547 | T | C | -0.011 | 0.010 | -0.010 | 0.015 | -0.011 | 0.015 | -0.009 | 0.012 | -0.016 | 0.016 |
| rs784256 | G | A | 0.011 | 0.013 | 0.010 | 0.018 | 0.013 | 0.018 | 0.006 | 0.015 | 0.006 | 0.020 |
| rs6129084 | T | A | -0.015 | 0.010 | -0.019 | 0.014 | -0.009 | 0.014 | -0.003 | 0.012 | -0.051 | 0.016 |
| rs9608249 | G | A | 0.014 | 0.014 | -0.019 | 0.019 | 0.046 | 0.019 | 0.015 | 0.016 | 0.002 | 0.021 |
| rs136553 | T | C | -0.014 | 0.010 | -0.024 | 0.014 | -0.005 | 0.014 | -0.007 | 0.012 | -0.011 | 0.016 |
| rs7288455 | A | G | -0.002 | 0.010 | -0.004 | 0.014 | -0.004 | 0.014 | 0.006 | 0.012 | -0.010 | 0.016 |
| crc colorectal, EA effect allele, OA other allele, re rectal, se standard error, SNP single nucleotide polymorphism | | | | | | | | | | | | |

| **Supplemental Table 7: Summary information on prostate cancer risk for the leisure television watching SNPs used for the Mendelian randomisation analyses** | | | | | | |
| --- | --- | --- | --- | --- | --- | --- |
| SNP | ΕΑ | ΟΑ | beta_prst | se_prst | beta_adv | se_adv |
| rs11810109 | A | T | -0.018 | 0.009 | -0.005 | 0.015 |
| rs112632642 | A | G | 0.022 | 0.016 | -0.005 | 0.028 |
| rs6670915 | C | T | -0.016 | 0.008 | -0.017 | 0.014 |
| rs12725114 | G | A | 0.011 | 0.010 | 0.025 | 0.017 |
| rs984409 | A | G | -0.008 | 0.008 | 0.009 | 0.014 |
| rs10890123 | C | T | 0.011 | 0.010 | 0.019 | 0.018 |
| rs71658797 | A | T | 0.008 | 0.013 | -0.001 | 0.022 |
| rs114328297 | T | G | -0.007 | 0.011 | -0.021 | 0.018 |
| rs631130 | A | T | 0.000 | 0.009 | -0.001 | 0.016 |
| rs148648822 | T | G | -0.012 | 0.012 | -0.007 | 0.020 |
| rs17379561 | T | A | -0.010 | 0.011 | -0.008 | 0.019 |
| rs55909997 | A | G | 0.003 | 0.009 | -0.002 | 0.015 |
| rs1278847 | C | A | -0.002 | 0.009 | -0.010 | 0.015 |
| rs141425667 | A | G | -0.028 | 0.018 | 0.013 | 0.031 |
| rs4845364 | G | A | 0.033 | 0.008 | 0.044 | 0.014 |
| rs6673341 | G | T | 0.004 | 0.008 | -0.010 | 0.014 |
| rs72725224 | C | A | -0.067 | 0.033 | -0.061 | 0.059 |
| rs10737620 | T | A | 0.004 | 0.009 | 0.014 | 0.016 |
| rs6721975 | C | T | NA | NA | -0.010 | 0.018 |
| rs11896615 | G | A | 0.002 | 0.012 | 0.022 | 0.020 |
| rs72781699 | A | G | -0.029 | 0.010 | -0.028 | 0.018 |
| rs12105701 | T | C | 0.001 | 0.008 | -0.017 | 0.014 |
| rs7564130 | C | T | -0.005 | 0.008 | -0.010 | 0.014 |
| rs11125515 | A | C | 0.001 | 0.009 | 0.001 | 0.016 |
| rs12475168 | T | G | 0.008 | 0.012 | -0.004 | 0.020 |
| rs183566425 | A | T | -0.044 | 0.042 | -0.015 | 0.074 |
| rs10189857 | G | A | -0.001 | 0.008 | 0.033 | 0.014 |
| rs62641636 | A | G | -0.006 | 0.009 | -0.008 | 0.015 |
| rs59210467 | T | C | 0.006 | 0.009 | 0.028 | 0.015 |
| rs11689199 | A | G | -0.004 | 0.009 | 0.001 | 0.015 |
| rs1451533 | A | G | -0.008 | 0.009 | 0.004 | 0.016 |
| rs2971640 | A | G | 0.003 | 0.008 | 0.007 | 0.014 |
| rs12476388 | C | T | 0.009 | 0.009 | -0.012 | 0.015 |
| rs374722 | G | A | 0.005 | 0.012 | -0.003 | 0.020 |
| rs3754970 | C | T | 0.001 | 0.008 | -0.006 | 0.014 |
| rs1913808 | G | A | 0.003 | 0.009 | -0.009 | 0.015 |
| rs2060199 | T | A | -0.010 | 0.008 | -0.013 | 0.014 |
| rs263771 | A | C | 0.009 | 0.010 | 0.006 | 0.017 |
| rs4577309 | A | G | NA | NA | -0.006 | 0.014 |
| rs34224261 | C | G | -0.002 | 0.008 | 0.015 | 0.014 |
| rs4675246 | T | G | -0.005 | 0.010 | -0.010 | 0.017 |
| rs13029509 | A | G | 0.000 | 0.008 | -0.001 | 0.014 |
| rs10932837 | T | C | 0.007 | 0.008 | 0.007 | 0.014 |
| rs4973576 | A | C | -0.021 | 0.009 | -0.011 | 0.015 |
| rs138256022 | T | C | 0.011 | 0.023 | 0.058 | 0.039 |
| rs9834970 | T | C | -0.005 | 0.009 | -0.010 | 0.015 |
| rs1352890 | T | C | -0.006 | 0.008 | -0.021 | 0.014 |
| rs3796386 | A | G | 0.027 | 0.008 | 0.025 | 0.014 |
| rs182355396 | A | C | 0.070 | 0.083 | -0.063 | 0.149 |
| rs11917871 | C | T | 0.011 | 0.009 | 0.005 | 0.015 |
| rs11130793 | C | T | 0.009 | 0.008 | 0.008 | 0.014 |
| rs11714337 | G | A | -0.016 | 0.008 | -0.033 | 0.014 |
| rs12496084 | G | A | -0.013 | 0.010 | -0.026 | 0.017 |
| rs6797840 | C | A | -0.007 | 0.008 | -0.023 | 0.014 |
| rs56398417 | C | T | -0.005 | 0.009 | 0.005 | 0.015 |
| rs2034768 | A | G | -0.007 | 0.008 | -0.008 | 0.014 |
| rs12632778 | G | T | -0.030 | 0.020 | -0.055 | 0.033 |
| rs9867121 | C | A | NA | NA | -0.007 | 0.018 |
| rs12491503 | A | G | NA | NA | 0.004 | 0.015 |
| rs114600294 | C | G | -0.002 | 0.013 | -0.008 | 0.022 |
| rs66852340 | T | C | -0.001 | 0.010 | 0.010 | 0.017 |
| rs7700107 | C | A | -0.004 | 0.012 | -0.010 | 0.021 |
| rs34811474 | G | A | 0.018 | 0.011 | 0.005 | 0.019 |
| rs77215114 | A | T | -0.027 | 0.017 | -0.024 | 0.030 |
| rs28971796 | A | G | -0.014 | 0.012 | -0.018 | 0.020 |
| rs6850494 | C | A | 0.011 | 0.008 | 0.007 | 0.014 |
| rs13107325 | T | C | -0.015 | 0.016 | -0.009 | 0.028 |
| rs62332760 | T | G | 0.053 | 0.008 | 0.041 | 0.014 |
| rs7693703 | G | A | 0.022 | 0.014 | 0.034 | 0.024 |
| rs4334769 | G | T | 0.013 | 0.008 | 0.007 | 0.014 |
| rs6825241 | A | C | -0.011 | 0.008 | -0.021 | 0.014 |
| rs7693082 | G | C | -0.004 | 0.009 | 0.035 | 0.015 |
| rs10940659 | G | A | -0.005 | 0.008 | -0.017 | 0.014 |
| rs2409838 | A | T | -0.013 | 0.008 | -0.009 | 0.014 |
| rs262890 | G | A | -0.002 | 0.009 | -0.011 | 0.015 |
| rs7716447 | G | A | -0.007 | 0.009 | 0.008 | 0.015 |
| rs1031423 | C | T | 0.008 | 0.010 | 0.013 | 0.018 |
| rs57585211 | G | T | -0.011 | 0.011 | -0.012 | 0.018 |
| rs9326862 | A | T | -0.002 | 0.008 | -0.008 | 0.014 |
| rs10041724 | T | C | 0.004 | 0.010 | 0.028 | 0.018 |
| rs10057545 | A | G | -0.008 | 0.009 | -0.011 | 0.015 |
| rs62379379 | T | G | -0.004 | 0.016 | -0.023 | 0.028 |
| rs10054327 | G | A | 0.008 | 0.008 | -0.003 | 0.014 |
| rs42210 | C | G | -0.010 | 0.010 | 0.008 | 0.016 |
| rs2336895 | G | A | -0.003 | 0.009 | 0.003 | 0.015 |
| rs72828890 | C | T | 0.023 | 0.014 | -0.019 | 0.024 |
| rs255292 | A | C | -0.003 | 0.008 | 0.012 | 0.014 |
| rs72834698 | G | A | -0.034 | 0.012 | -0.034 | 0.020 |
| rs3135044 | T | C | 0.022 | 0.008 | 0.023 | 0.014 |
| rs10947452 | T | C | 0.000 | 0.009 | 0.017 | 0.015 |
| rs9471333 | C | T | 0.002 | 0.009 | -0.023 | 0.015 |
| rs4523073 | G | A | -0.005 | 0.008 | -0.008 | 0.015 |
| rs77589760 | C | G | NA | NA | 0.001 | 0.053 |
| rs6905544 | G | A | 0.001 | 0.008 | 0.013 | 0.014 |
| rs17789218 | T | C | -0.008 | 0.011 | -0.019 | 0.018 |
| rs78394231 | C | T | 0.042 | 0.014 | 0.006 | 0.024 |
| rs2184364 | A | G | -0.020 | 0.010 | -0.013 | 0.017 |
| rs9718104 | G | T | 0.002 | 0.018 | -0.021 | 0.032 |
| rs17568389 | T | A | 0.005 | 0.008 | -0.006 | 0.014 |
| rs10234444 | G | A | 0.010 | 0.012 | 0.022 | 0.020 |
| rs6973656 | G | A | 0.005 | 0.009 | 0.014 | 0.015 |
| rs973734 | C | A | 0.014 | 0.011 | 0.013 | 0.020 |
| rs3137 | G | A | 0.002 | 0.011 | 0.007 | 0.019 |
| rs2694151 | C | T | 0.012 | 0.018 | -0.032 | 0.031 |
| rs62471080 | C | G | -0.003 | 0.008 | -0.001 | 0.014 |
| rs7788008 | G | A | -0.006 | 0.008 | -0.003 | 0.014 |
| rs10246289 | A | G | 0.002 | 0.013 | -0.004 | 0.021 |
| rs11763734 | C | A | 0.007 | 0.008 | -0.002 | 0.014 |
| rs7834121 | T | G | -0.017 | 0.008 | -0.030 | 0.014 |
| rs1421334 | A | C | 0.010 | 0.008 | 0.028 | 0.014 |
| rs6996198 | T | C | -0.011 | 0.011 | -0.009 | 0.019 |
| rs6472942 | C | T | -0.010 | 0.008 | -0.011 | 0.014 |
| rs11985394 | T | A | 0.000 | 0.008 | 0.014 | 0.014 |
| rs72671494 | C | T | 0.010 | 0.013 | 0.031 | 0.021 |
| rs12541615 | C | T | -0.033 | 0.011 | -0.050 | 0.018 |
| rs2717559 | A | G | NA | NA | -0.016 | 0.015 |
| rs2616830 | G | A | -0.004 | 0.008 | -0.002 | 0.014 |
| rs2291681 | G | T | 0.008 | 0.009 | 0.028 | 0.014 |
| rs34864022 | G | A | -0.024 | 0.017 | 0.024 | 0.028 |
| rs12554512 | T | C | -0.014 | 0.009 | -0.008 | 0.014 |
| rs2791574 | G | T | -0.001 | 0.009 | 0.010 | 0.015 |
| rs141184308 | A | G | NA | NA | 0.065 | 0.074 |
| rs870151 | A | T | -0.013 | 0.008 | -0.011 | 0.014 |
| rs2787374 | T | C | -0.015 | 0.008 | 0.003 | 0.014 |
| rs10739499 | C | G | -0.007 | 0.009 | -0.003 | 0.015 |
| rs7043521 | A | T | -0.004 | 0.008 | -0.016 | 0.014 |
| rs12342024 | T | G | -0.037 | 0.013 | -0.045 | 0.022 |
| rs4382592 | T | G | -0.013 | 0.009 | -0.027 | 0.015 |
| rs2073869 | C | T | -0.006 | 0.011 | -0.022 | 0.018 |
| 9:139926402_T_C | C | T | 0.004 | 0.008 | NA | NA |
| rs1623025 | C | G | 0.010 | 0.008 | 0.023 | 0.014 |
| rs1243182 | T | C | -0.010 | 0.009 | -0.008 | 0.015 |
| rs2045147 | A | G | -0.002 | 0.008 | 0.003 | 0.014 |
| rs10994943 | T | G | -0.017 | 0.008 | -0.022 | 0.014 |
| rs11201422 | T | C | 0.011 | 0.009 | 0.017 | 0.015 |
| rs10786658 | T | A | -0.006 | 0.008 | -0.017 | 0.014 |
| rs7089973 | A | C | -0.007 | 0.009 | -0.048 | 0.015 |
| rs11245482 | C | T | 0.023 | 0.008 | 0.021 | 0.014 |
| rs17727474 | C | T | 0.004 | 0.011 | 0.037 | 0.020 |
| rs12289262 | T | C | -0.015 | 0.009 | -0.011 | 0.016 |
| rs801733 | A | C | 0.002 | 0.009 | -0.004 | 0.015 |
| rs116175338 | T | G | NA | NA | 0.002 | 0.016 |
| rs12272012 | C | G | 0.011 | 0.020 | 0.006 | 0.034 |
| rs11020045 | C | A | 0.010 | 0.009 | -0.015 | 0.015 |
| rs17207890 | G | A | -0.001 | 0.009 | 0.018 | 0.015 |
| rs10791546 | C | T | 0.008 | 0.008 | -0.010 | 0.015 |
| rs113584870 | G | A | -0.028 | 0.012 | NA | NA |
| rs328277 | C | A | 0.002 | 0.008 | 0.030 | 0.014 |
| rs648044 | G | A | -0.003 | 0.009 | -0.010 | 0.016 |
| rs11218575 | C | T | 0.002 | 0.009 | 0.009 | 0.015 |
| rs74802478 | G | A | 0.002 | 0.011 | -0.041 | 0.018 |
| rs4937842 | C | G | -0.004 | 0.008 | -0.011 | 0.014 |
| rs10772643 | C | T | 0.029 | 0.013 | NA | NA |
| rs74996610 | G | C | 0.017 | 0.020 | NA | NA |
| rs10771746 | T | C | 0.021 | 0.009 | NA | NA |
| rs10876864 | A | G | -0.017 | 0.008 | -0.026 | 0.014 |
| rs8756 | A | C | 0.002 | 0.008 | 0.006 | 0.014 |
| rs11179838 | C | G | 0.004 | 0.008 | NA | NA |
| rs2164744 | C | T | -0.018 | 0.009 | NA | NA |
| rs2173650 | G | T | 0.010 | 0.012 | NA | NA |
| rs1790135 | T | C | -0.005 | 0.009 | NA | NA |
| rs9563168 | G | A | 0.007 | 0.009 | 0.030 | 0.017 |
| rs9569734 | A | G | 0.007 | 0.012 | 0.026 | 0.019 |
| rs9528023 | C | T | -0.012 | 0.010 | -0.017 | 0.017 |
| rs56858768 | A | G | 0.000 | 0.009 | -0.011 | 0.015 |
| rs553397860 | A | G | -0.010 | 0.012 | -0.010 | 0.020 |
| rs7991062 | G | C | -0.010 | 0.009 | -0.010 | 0.015 |
| rs11629120 | C | T | 0.024 | 0.008 | 0.035 | 0.014 |
| rs405797 | A | T | 0.008 | 0.010 | -0.007 | 0.018 |
| rs1022785 | G | A | -0.011 | 0.012 | -0.018 | 0.020 |
| rs10145592 | G | C | 0.006 | 0.009 | 0.020 | 0.015 |
| rs7157001 | G | A | NA | NA | -0.007 | 0.019 |
| rs8043253 | T | C | 0.000 | 0.008 | -0.015 | 0.014 |
| rs2460 | A | G | -0.020 | 0.010 | -0.039 | 0.017 |
| rs4775373 | T | C | 0.006 | 0.009 | 0.010 | 0.015 |
| rs61331678 | G | C | 0.012 | 0.008 | 0.023 | 0.014 |
| rs35574015 | C | T | -0.016 | 0.009 | -0.031 | 0.015 |
| rs7189927 | T | C | -0.003 | 0.009 | -0.013 | 0.015 |
| rs749671 | G | A | -0.012 | 0.008 | -0.015 | 0.014 |
| rs7184800 | G | A | -0.015 | 0.009 | -0.013 | 0.015 |
| rs142710267 | T | G | NA | NA | 0.024 | 0.015 |
| rs2447098 | A | C | 0.005 | 0.009 | 0.015 | 0.015 |
| rs11654952 | G | T | NA | NA | 0.011 | 0.020 |
| rs2584597 | T | C | 0.008 | 0.009 | -0.025 | 0.015 |
| rs9902312 | T | C | -0.003 | 0.009 | -0.009 | 0.015 |
| rs11657730 | C | T | 0.009 | 0.009 | -0.003 | 0.016 |
| rs303753 | A | G | 0.006 | 0.009 | 0.008 | 0.015 |
| rs71362609 | C | T | -0.013 | 0.008 | -0.029 | 0.014 |
| rs11081851 | A | G | -0.012 | 0.008 | 0.000 | 0.014 |
| rs9964724 | C | T | 0.003 | 0.009 | -0.023 | 0.015 |
| rs1156541 | C | T | 0.016 | 0.010 | 0.021 | 0.017 |
| rs148544378 | C | T | -0.003 | 0.043 | 0.079 | 0.072 |
| rs17512836 | T | C | 0.058 | 0.026 | 0.060 | 0.045 |
| rs7248205 | C | T | -0.002 | 0.009 | 0.009 | 0.015 |
| rs111901094 | T | G | 0.024 | 0.012 | 0.026 | 0.021 |
| rs6131281 | C | T | 0.009 | 0.009 | 0.019 | 0.015 |
| rs6141814 | A | C | -0.035 | 0.008 | -0.038 | 0.014 |
| rs4810312 | G | C | -0.010 | 0.008 | -0.032 | 0.014 |
| rs6124476 | T | C | 0.001 | 0.009 | 0.009 | 0.014 |
| rs6031440 | A | T | 0.006 | 0.009 | 0.016 | 0.015 |
| rs55700114 | A | G | -0.001 | 0.009 | 0.009 | 0.015 |
| rs6125906 | A | C | 0.024 | 0.015 | 0.054 | 0.025 |
| rs56103247 | C | T | 0.004 | 0.020 | 0.079 | 0.035 |
| rs10427502 | G | A | -0.004 | 0.009 | 0.017 | 0.015 |
| rs7609 | A | T | -0.020 | 0.010 | 0.015 | 0.017 |
| rs2092829 | G | A | -0.012 | 0.009 | -0.008 | 0.015 |
| rs150399729 | C | G | 0.026 | 0.025 | -0.006 | 0.040 |
| adv adcanced, EA effect allele, OA other allele, NA not available, prst prostate, se standard error, SNP single nucleotide polymorphism | | | | | | |

| **Supplemental Table 8: Summary information on prostate cancer risk for the leisure computer use SNPs used for the Mendelian randomisation analyses** | | | | | | |
| --- | --- | --- | --- | --- | --- | --- |
| SNP | ΕΑ | ΟΑ | beta_prst | se_prst | beta_adv | se_adv |
| rs2748985 | C | T | -0.021 | 0.009 | -0.022 | 0.016 |
| rs12145677 | A | G | 0.000 | 0.009 | 0.007 | 0.015 |
| rs7526456 | C | T | 0.015 | 0.009 | -0.002 | 0.015 |
| rs10754920 | C | G | 0.003 | 0.013 | 0.003 | 0.022 |
| rs6744254 | T | C | -0.006 | 0.008 | -0.017 | 0.014 |
| rs11274218 | T | C | 0.002 | 0.008 | 0.000 | 0.015 |
| rs4592851 | T | G | 0.004 | 0.009 | 0.003 | 0.016 |
| rs10208088 | C | T | -0.003 | 0.008 | -0.005 | 0.014 |
| rs11708955 | C | T | -0.027 | 0.009 | -0.016 | 0.015 |
| rs6774533 | T | C | 0.000 | 0.009 | -0.008 | 0.016 |
| rs78082503 | C | G | 0.003 | 0.010 | -0.004 | 0.017 |
| rs6857629 | G | A | -0.008 | 0.010 | -0.023 | 0.016 |
| rs2068625 | C | T | -0.006 | 0.009 | -0.034 | 0.015 |
| rs2220599 | G | C | -0.011 | 0.009 | -0.017 | 0.015 |
| rs11749912 | A | G | -0.003 | 0.008 | -0.020 | 0.014 |
| rs1469249 | G | A | 0.009 | 0.010 | 0.015 | 0.017 |
| rs162894 | T | G | -0.013 | 0.008 | -0.005 | 0.014 |
| rs56158102 | C | T | NA | NA | 0.003 | 0.015 |
| rs9477970 | A | T | -0.004 | 0.010 | -0.017 | 0.018 |
| rs2179517 | C | G | 0.009 | 0.008 | 0.017 | 0.014 |
| rs9372625 | A | G | 0.003 | 0.009 | -0.012 | 0.015 |
| rs17789218 | C | T | 0.008 | 0.011 | 0.019 | 0.018 |
| rs6935828 | T | C | 0.015 | 0.008 | 0.009 | 0.014 |
| rs55772938 | G | A | 0.005 | 0.009 | 0.007 | 0.016 |
| rs12706626 | A | G | 0.009 | 0.008 | -0.015 | 0.014 |
| rs3944151 | A | G | 0.001 | 0.009 | -0.005 | 0.015 |
| rs2345941 | A | G | -0.008 | 0.008 | -0.028 | 0.014 |
| rs13262595 | G | A | 0.020 | 0.008 | 0.019 | 0.014 |
| rs4977839 | A | G | 0.014 | 0.009 | 0.010 | 0.014 |
| rs113851275 | A | G | -0.008 | 0.013 | -0.009 | 0.022 |
| rs7020477 | A | G | -0.007 | 0.010 | 0.000 | 0.017 |
| rs73578186 | C | T | 0.017 | 0.009 | 0.034 | 0.015 |
| rs2734849 | G | A | -0.001 | 0.008 | -0.001 | 0.014 |
| rs1448355 | T | C | -0.005 | 0.008 | -0.024 | 0.015 |
| rs35933007 | A | G | 0.003 | 0.011 | NA | NA |
| rs1319734 | A | G | -0.008 | 0.008 | NA | NA |
| rs206965 | T | C | -0.017 | 0.010 | -0.013 | 0.017 |
| rs12874776 | A | C | 0.009 | 0.014 | 0.021 | 0.024 |
| rs198262 | C | T | -0.020 | 0.021 | -0.030 | 0.035 |
| rs166835 | C | T | -0.002 | 0.008 | -0.012 | 0.014 |
| rs4702 | G | A | 0.018 | 0.008 | 0.025 | 0.014 |
| rs6498759 | T | C | 0.010 | 0.009 | -0.020 | 0.016 |
| rs3730399 | A | G | 0.031 | 0.016 | 0.048 | 0.027 |
| rs4073003 | A | G | 0.012 | 0.012 | 0.035 | 0.021 |
| rs7209653 | T | C | 0.002 | 0.009 | 0.014 | 0.016 |
| rs12603813 | T | C | 0.007 | 0.009 | 0.020 | 0.016 |
| rs66643547 | T | C | -0.007 | 0.009 | 0.002 | 0.016 |
| rs784256 | G | A | -0.018 | 0.011 | -0.046 | 0.018 |
| rs6129084 | T | A | 0.013 | 0.009 | 0.016 | 0.015 |
| rs9608249 | G | A | NA | NA | 0.006 | 0.020 |
| rs136553 | T | C | 0.015 | 0.008 | 0.015 | 0.014 |
| rs7288455 | A | G | 0.006 | 0.009 | 0.013 | 0.015 |
| adv adcanced, EA effect allele, OA other allele, NA not available, prst prostate, se standard error, SNP single nucleotide polymorphism | | | | | | |

| **Supplemental Table 9: Sample size and power calculations in Mendelian randomization study of sedentary behaviour and cancer risk** | | | | | | | | |
| --- | --- | --- | --- | --- | --- | --- | --- | --- |
| **Exposure** | **Outcome** | **Sample size** | **Proportion of cases** | **Selected scenarios^*^** | | | |  |
|  |  |  |  | **OR=1.05** | **OR=1.1** | **OR=1.15** | **OR=1.2** | **OR=1.25** |
| **Television use** | Breast cancer | 228,951 | 0.54 | 0.38 | 0.89 | 1 | 1 | 1 |
|  | Colorectal cancer | 98,715 | 0.53 | 0.19 | 0.56 | 0.87 | 0.98 | 1 |
|  | Prostate cancer | 140,254 | 0.56 | 0.25 | 0.7 | 0.95 | 1 | 1 |
| **Computer use** | Breast cancer | 228,951 | 0.54 | 0.13 | 0.36 | 0.65 | 0.86 | 0.96 |
|  | Colorectal cancer | 98,715 | 0.53 | 0.08 | 0.18 | 0.34 | 0.52 | 0.69 |
|  | Prostate cancer | 140,254 | 0.56 | 0.1 | 0.24 | 0.45 | 0.66 | 0.83 |
| **Driving** | Breast cancer | 228,951 | 0.54 | 0.06 | 0.08 | 0.12 | 0.16 | 0.22 |
|  | Colorectal cancer | 98,715 | 0.53 | 0.05 | 0.06 | 0.08 | 0.10 | 0.12 |
|  | Prostate cancer | 140,254 | 0.56 | 0.05 | 0.07 | 0.09 | 0.12 | 0.15 |
| Abbreviations: OR, odds ratio  ^*^ Type 1 error of 5% and a proportion of variance explained equal to 2%, 0.5%, and 0.05% are assumed for television use, computer use and driving respectively | | | | | | | |  |

| **Supplemental Table 22: Multivariable Mendelian randomization estimates between leisure television watching, computer use and cancer risk** | | | | |
| --- | --- | --- | --- | --- |
| **OR* (95% CI) adjusted for:** | | | | |
| **Traits** | **Alcohol** | **BMI** | **Years of education** | **Smoking** |
| **Cancer Type** |  |  |  |  |
| **Television watching** | |  |  |  |
| **Breast cancer** |  |  |  |  |
| Overall | 1.15 (1.04, 1.28) | 1.27 (1.13, 1.43) | 1.08 (0.92, 1.27) | 1.12 (0.99, 1.27) |
| Luminal A | 1.21 (1.05, 1.39) | 1.30 (1.12, 1.52) | 1.11 (0.90, 1.35) | 1.19 (1.01, 1.39) |
| Luminal B | 1.09 (0.86, 1.39) | 1.17 (0.91, 1.51) | 0.93 (0.66, 1.31) | 1.13 (0.87, 1.48) |
| HER2 negative | 1.22 (1.00, 1.51) | 1.32 (1.07, 1.65) | 1.12 (0.84, 1.51) | 1.17 (0.93, 1.49) |
| HER2 enriched | 1.23 (0.89, 1.70) | 1.46 (1.05, 2.05) | 1.08 (0.66, 1.79) | 1.15 (0.79, 1.67) |
| Triple negative | 1.19 (0.97, 1.04) | 1.39 (1.12, 1.72) | 1.07 (0.81, 1.40) | 1.16 (0.92, 1.46) |
| **Colorectal cancer** |  |  |  |  |
| Overall | 1.26 (1.09, 1.46) | 1.20 (1.03, 1.38) | 1.08 (0.90, 1.31) | 1.17 (1.00, 1.39) |
| Men | 1.40 (1.17, 1.70) | 1.32 (1.09, 1.58) | 1.13 (0.88, 1.45) | 1.32 (1.08, 1.62) |
| Women | 1.17 (0.97, 1.40) | 1.11 (0.92, 1.34) | 1.07 (0.83, 1.38) | 1.08 (0.89, 1.34) |
| Colon | 1.26 (1.07, 1.49) | 1.20 (1.02, 1.40) | 1.07 (0.86, 1.32) | 1.17 (0.99, 1.40) |
| Rectal | 1.49 (1.20, 1.84) | 1.43 (1.16, 1.75) | 1.07 (0.79, 1.45) | 1.35 (1.05, 1.75) |
| **Computer use** |  |  |  |  |
| **Breast cancer** |  |  |  |  |
| Overall | 1.05 (0.93, 1.19) | 1.06 (0.87, 1.30) | 1.20 (0.99, 1.45) | 1.01 (0.84, 1.21) |
| Luminal A | 1.04 (0.90, 1.20) | 1.05 (0.84, 1.32) | 1.26 (1.00, 1.60) | 1.06 (0.85, 1.31) |
| Luminal B | 0.95 (0.73, 1.23) | 1.00 (0.70, 1.45) | 0.91 (0.60, 1.40) | 0.88 (0.59, 1.30) |
| HER2 negative | 1.01 (0.83, 1.32) | 1.14 (0.83, 1.57) | 1.32 (0.94, 1.88) | 1.09 (0.80, 1.49) |
| HER2 enriched | 1.32 (0.98, 1.79) | 0.77 (0.48, 1.26) | 1.06 (0.59, 1.90) | 0.66 (0.39, 1.08) |
| Triple negative | 1.06 (0.87, 1.30) | 0.73 (0.52, 1.01) | 0.79 (0.57, 1.09) | 0.68 (0.49, 0.95) |
| **Colorectal cancer** |  |  |  |  |
| Overall | 1.14 (0.99, 1.32) | 0.81 (0.66, 0.99) | 1.01 (0.79, 1.27) | 0.90 (0.72, 1.12) |
| Men | 1.11 (0.93, 1.32) | 0.82 (0.61, 1.09) | 0.89 (0.65, 1.20) | 0.84 (0.63, 1.09) |
| Women | 1.20 (0.99, 1.43) | 0.87 (0.66, 1.14) | 1.14 (0.84, 1.54) | 0.98 (0.72, 1.34) |
| Colon | 1.19 (1.00, 1.42) | 0.84 (0.66, 1.05) | 0.99 (0.76, 1.30) | 0.90 (0.71, 1.16) |
| Rectal | 1.09 (0.89, 1.34) | 0.63 (0.48, 0.84) | 0.92 (0.65, 1.30) | 0.64 (0.46, 0.90) |
| Abbreviations: BMI Body mass index, CI Confidence intervals, HER2 Human epidermal growth factor receptor 2, OR Odds ratio, SNPs Single nucleotide polymorphism  * The estimates correspond to a standard deviation increase in leisure television watching or computer use | | | | |

| **Supplemental Table 23: Outlying SNPs identified by MR-PRESSO** | | | | | |
| --- | --- | --- | --- | --- | --- |
| **Exposure** |  | | | | |
|  | **Colorectal cancer in women** | **Overall breast cancer** | **Luminal A breast cancer** | **Luminal B breast cancer** | **Overall prostate cancer** |
| **Television watching** |  | rs1243182  rs11714337 | rs1243182  rs11714337 |  | rs4845364  rs62332760  rs6141814 |
| **Computer use** | rs3730399 | rs11708955  rs162894  rs4702 | rs162894  rs113851275 | rs4702 | rs11708955 |
| MR-PRESSO MR pleiotropy residual sum and outlier test | | | | | |

| **Supplemental Table 24: Evidence of association (p<5*10-8) with secondary traits of the SNPs (or their proxies) used as genetic instruments for Mendelian randomization analyses of sedentary behaviours and risk of breast and colorectal cancer** | | | |
| --- | --- | --- | --- |
| SNP | Chr | Gene | Diseases & traits |
| **Computer** |  |  |  |
| rs2748985 | 1 | CFAP74 | Arm fat-free mass (UK Biobank), Arm predicted mass (UK Biobank) Basal metabolic rate (UK Biobank), Body mass index (UK Biobank), Impedance of arm/whole body (UK Biobank), Trunk fat-free mass (UK Biobank), Trunk predicted mass (UK Biobank) |
| rs12145677 | 1 | SYPL2 | Impedance of arm/leg/whole body (UK Biobank), Qualifications: college or university degree (UK Biobank), Time spent watching television (UK Biobank) |
| rs7526456 | 1 |  |  |
| rs10754920 | 2 |  |  |
| rs6744254 | 2 | KCNJ3 |  |
| rs11274218 | 2 | SPAG16 |  |
| rs4592851 | 2 | VWC2L |  |
| rs10208088 | 2 | LOC105373893 |  |
| rs11708955 | 3 | DAG1 | Mean corpuscular volume (PMID: 27863252), Reticulocyte count (PMID: 27863252), Crohns disease (PMID: 26192919), Inflammatory bowel disease (PMID: 26192919), Ulcerative colitis (PMID: 26192919), Primary sclerosing cholangitis (PMID: 27992413), Years of educational attainment (PMID: 27225129), Arm fat mass/percentage (UK Biobank), Body mass index (UK Biobank), Time spent watching television (UK Biobank), Diastolic blood pressure (UK Biobank), Leg fat mass/percentage (UK Biobank), Trunk fat mass/percentage (UK Biobank), Whole body fat mass (UK Biobank), Weight (UK Biobank), Age at menarche (PMID: 25231870), Fed-up/miserableness (UK Biobank), Fluid intelligence score (UK Biobank) |
| rs6774533 | 3 | CADPS | Arm fat mass right (UK Biobank), Hip circumference (UK Biobank), Trunk fat mass (UK Biobank), Waist circumference (UK Biobank), Whole body fat mass (UK Biobank), Qualifications: college or university degree (UK Biobank) |
| rs78082503 | 3 |  |  |
| rs6857629 | 4 |  |  |
| rs2068625 | 4 | C4orf45 | Overall health rating (UK Biobank), Pulse rate (UK Biobank) |
| rs2220599 | 5 | LINC02142 | Number of days or week walked 10+ minutes (UK Biobank), Subjective well being (PMID: 29292387) |
| rs11749912 | 5 | MEF2C | Trunk fat percentage (UK Biobank), Years of educational attainment (PMID: 27225129) |
| rs1469249 | 5 | LOC101927078 | Self-reported hypertension (UK Biobank), Years of educational attainment (PMID: 27225129) |
| rs162894 | 5 |  | Eosinophil count (PMID: 27863252), Granulocyte count (PMID: 27863252), Mean platelet volume (PMID: 27863252), Monocyte count (PMID: 27863252), Myeloid white cell count (PMID: 27863252), Height (PMID: 25282103), Asthma (UK Biobank), Basal metabolic rate (UK Biobank), Hand grip strength right (UK Biobank), Leg fat-free mass (UK Biobank), Leg predicted mass (UK Biobank), Trunk fat-free mass (UK Biobank), Trunk predicted mass (UK Biobank), Whole body fat-free mass (UK Biobank) |
| rs56158102 | 5 |  |  |
| rs9477970 | 6 | LOC101928519 | Arm fat-free mass (UK Biobank), Arm predicted mass (UK Biobank)  Basal metabolic rate (UK Biobank), Height (UK Biobank), Impedance of arm/whole body (UK Biobank), Leg predicted mass (UK Biobank), Leg fat-free mass (UK Biobank), Trunk fat-free mass (UK Biobank), Trunk predicted mass (UK Biobank), Whole body fat-free mass (UK Biobank), Qualifications: college or university degree (UK Biobank) |
| rs2179517 | 6 | H3C4 | Arm fat mass (UK Biobank), Arm fat percentage (UK Biobank), Arm fat-free mass (UK Biobank), Basal metabolic rate (UK Biobank), Body fat percentage (UK Biobank), Height (UK Biobank), Hip circumference (UK Biobank), Leg fat mass (UK Biobank), Leg fat percentage (UK Biobank), Leg fat-free mass (UK Biobank), Leg predicted mass (UK Biobank), Mean corpuscular hemoglobin (PMID: 27863252), Qualifications: college or university degree (UK Biobank), Trunk fat mass (UK Biobank), Trunk fat percentage (UK Biobank), Trunk fat-free mass (UK Biobank), Trunk predicted mass (UK Biobank), Waist circumference (UK Biobank), Weight (UK Biobank)  Whole body fat mass (UK Biobank), Whole body fat-free mass (UK Biobank) |
| rs9372625 | 6 |  | Alcohol intake frequency (UK Biobank), Arm fat mass (UK Biobank), Arm fat percentage (UK Biobank), Body fat percentage (UK Biobank), Body mass index (UK Biobank), Leg fat mass (UK Biobank), Leg fat percentage (UK Biobank), Overall health rating (UK Biobank), Time spent watching television (UK Biobank), Trunk fat mass (UK Biobank), Whole body fat mass (UK Biobank), Years of educational attainment (PMID: 27225129), Number of days or week walked 10+ minutes (UK Biobank), Sodium in urine (UK Biobank), Weight (UK Biobank), Fluid intelligence score (UK Biobank) |
| rs17789218 | 6 | LOC105377911 | Arm fat mass (UK Biobank), Basal metabolic rate (UK Biobank), Comparative height size at age 10 (UK Biobank), Diastolic blood pressure (UK Biobank), Hip circumference (UK Biobank), Impedance of leg (UK Biobank), Leg fat-free mass (UK Biobank), Leg predicted mass (UK Biobank), Trunk fat mass (UK Biobank), Weight (UK Biobank), Whole body fat mass (UK Biobank) |
| rs6935828 | 6 |  | Hayfever, allergic rhinitis or eczema (UK Biobank) |
| rs55772938 | 7 | AUTS2 | Trunk fat mass (UK Biobank), Weight (UK Biobank) |
| rs12706626 | 7 | POT1 |  |
| rs3944151 | 7 |  | Pulse rate (UK Biobank) |
| rs2345941 | 7 | EXOC4 |  |
| rs13262595 | 8 | TSNARE1 | Nervous feelings (UK Biobank), Schizophrenia (PMID: 26198764)  Tense or highly strung (UK Biobank), Qualifications-degree (UK Biobank) |
| rs4977839 | 9 |  | Bipolar disorder (PMID: 27329760), Basal metabolic rate (UK Biobank), Fluid intelligence score (UK Biobank), Impedance of leg (UK Biobank), Leg fat-free mass (UK Biobank), Leg predicted mass (UK Biobank), Time spent watching television (UK Biobank), Whole body fat-free mass (UK Biobank), Years of educational attainment (PMID: 27225129), Qualifications-degree (UK Biobank) |
| rs113851275 | 9 | LOC105376157 | Arm fat-free mass (UK Biobank), Arm predicted mass (UK Biobank), Basal metabolic rate (UK Biobank), Birth weight (UK Biobank), Forced expiratory volume in 1-second (UK Biobank), Forced vital capacity (UK Biobank), Height (UK Biobank), Impedance of leg (UK Biobank), Leg fat-free mass (UK Biobank), Leg predicted mass (UK Biobank), Trunk fat-free mass (UK Biobank), Trunk predicted mass (UK Biobank), Whole body fat-free mass (UK Biobank), Whole body water mass (UK Biobank), Sensitivity or hurt feelings (UK Biobank) |
| rs7020477 | 9 | AMBP |  |
| rs73578186 | 9 | DENND1A |  |
| rs2734849 | 11 | ANKK1 | Neuroticism score (UK Biobank) |
| rs1448355 | 11 | NTM |  |
| rs35933007 | 12 |  | Morning or evening person (UK Biobank) |
| rs1319734 | 12 | SLC2A13 |  |
| rs206965 | 12 |  |  |
| rs12874776 | 13 |  |  |
| rs198262 | 14 | OTX2-AS1 |  |
| rs166835 | 15 | SEMA6D |  |
| rs4702 | 15 | FES | Schizophrenia (PMID: 26198764), Diastolic blood pressure (UK Biobank), Systolic blood pressure (UK Biobank) |
| rs6498759 | 16 | LOC107984893 |  |
| rs3730399 | 16 | E2F4 | Mean corpuscular haemoglobin, (PMID: 27863252), High density lipoprotein (PMID: 24097068) |
| rs4073003 | 17 | EPN2 |  |
| rs7209653 | 17 | AKAP10 | Mean corpuscular haemoglobin (PMID: 27863252), Reticulocyte count (PMID: 27863252) |
| rs12603813 | 17 | PLCD3 | Arm fat-free mass (UK Biobank), Arm predicted mass (UK Biobank)  Basal metabolic rate (UK Biobank), Height (UK Biobank), Impedance of arm/leg/whole body (UK Biobank), Leg fat-free mass (UK Biobank), Leg predicted mass (UK Biobank), Sleeplessness or insomnia (UK Biobank), Systolic blood pressure (UK Biobank), Trunk fat-free mass (UK Biobank), Trunk predicted mass (UK Biobank), Whole body fat-free mass (UK Biobank) |
| rs66643547 | 17 | BAHCC1- MIR4740 | Height (UK Biobank), Forced vital capacity (UK Biobank), Qualifications-degree (UK Biobank) |
| rs784256 | 18 | LOC105372130 | Body mass index (UK Biobank), Creatinine in urine (UK Biobank), Impedance of arm/leg/whole body (UK Biobank), Mood swings (UK Biobank), Fed-up feelings (UK Biobank) Overall health rating (UK Biobank), Potassium in urine (UK Biobank), Usual walking pace (UK Biobank), Qualifications-degree (UK Biobank), Hand grip strength (UK Biobank), Fuchss corneal dystrophy (UK Biobank) |
| rs6129084 | 20 | CDH4 | Educational attainment years of education (PMID: 27225129), Qualifications-degree (UK Biobank) |
| rs9608249 | 22 |  |  |
| rs136553 | 22 | LOC110091768 |  |
| rs7288455 | 22 | CACNA1I |  |
| **Television** |  |  |  |
| rs11810109 | 1 | ST3GAL3 | Age at menarche (UK Biobank), Comparative height size at age 10 (UK Biobank), Fluid intelligence score (UK Biobank), Years of educational attainment (PMID:27225129) |
| rs112632642 | 1 | FAF1 |  |
| rs6670915 | 1 |  |  |
| rs12725114 | 1 | PATJ |  |
| rs984409 | 1 | SGIP1 |  |
| rs10890123 | 1 | TNNI3K |  |
| rs71658797 | 1 | AK5 | Lung cancer (PMID: 28604730), Arm fat mass (UK Biobank), Arm fat percentage (UK Biobank), Arm fat-free mass (UK Biobank), Arm predicted mass (UK Biobank), Basal metabolic rate (UK Biobank), Body mass index (UK Biobank), Creatinine in urine (UK Biobank), Height (UK Biobank), Hip circumference (UK Biobank), Impedance of leg/whole body (UK Biobank), Leg fat mass (UK Biobank), Leg fat percentage (UK Biobank), Leg fat-free mass (UK Biobank), Leg predicted mass (UK Biobank), Trunk fat mass (UK Biobank), Trunk fat percentage (UK Biobank), Trunk fat-free mass (UK Biobank), Trunk predicted mass (UK Biobank), Waist circumference (UK Biobank), Weight (UK Biobank), Whole body fat mass (UK Biobank), Whole body fat-free mass (UK Biobank) |
| rs114328297 | 1 |  | Average weekly beer plus cider intake (UK Biobank), Morning or evening person (UK Biobank), Chronotype (PMID: 27494321) |
| rs631130 | 1 |  |  |
| rs148648822 | 1 | DPYD |  |
| rs17379561 | 1 | DPYD | Arm fat mass (UK Biobank), Arm fat percentage (UK Biobank), Body fat percentage (UK Biobank), Leg fat mass (UK Biobank), Leg fat percentage (UK Biobank), Overall health rating (UK Biobank), Trunk fat mass (UK Biobank), Trunk fat percentage (UK Biobank), Waist circumference (UK Biobank), Whole body fat mass (UK Biobank), Years of educational attainment (PMID: 27225129), ), Qualifications-degree (UK Biobank) |
| rs55909997 | 1 |  | Arm fat mass (UK Biobank), Leg fat mass (UK Biobank), Trunk fat mass (UK Biobank), Waist circumference (UK Biobank), Weight (UK Biobank), Whole body fat mass (UK Biobank) |
| rs1278847 | 1 | CYB561D1 | Impedance of arm/leg/whole body (UK Biobank), Time spent using computer (UK Biobank), Low density lipoprotein (PMID: 24097068), Total cholesterol (PMID: 24097068) |
| rs141425667 | 1 |  |  |
| rs4845364 | 1 | TPM3 | Platelet count (PMID: 27863252) |
| rs6673341 | 1 |  | Leg fat percentage (UK Biobank), Trunk fat percentage (UK Biobank), Body fat percentage (UK Biobank), Body mass index (UK Biobank) |
| rs72725224 | 1 |  |  |
| rs10737620 | 1 | RO60 |  |
| rs6721975 | 2 | SOX11 |  |
| rs11896615 | 2 |  |  |
| rs72781699 | 2 | FKBP1B | Lymphocyte count (PMID: 27863252), Red blood cell count (PMID: 27863252), White blood cell count (PMID: 27863252), Height (UK Biobank), Trunk fat-free mass (UK Biobank), Trunk predicted mass (UK Biobank), Alcohol intake frequency (UK Biobank), Whole body fat-free mass (UK Biobank) |
| rs12105701 | 2 | LOC105374506 |  |
| rs7564130 | 2 | NRXN1 |  |
| rs11125515 | 2 |  |  |
| rs12475168 | 2 | LINC01122 | Mean corpuscular hemoglobin (PMID:27863252), Mean corpuscular volume (PMID:27863252), Red blood cell count (PMID:27863252) |
| rs183566425 | 2 |  |  |
| rs10189857 | 2 | BCL11A | Red blood cell count (PMID: 27863252), Height (UK Biobank), Years of educational attainment (PMID: 27225129), Cognitive ability multi trait analysis (PMID: 29186694), Qualifications-degree (UK Biobank) |
| rs62641636 | 2 | PPP3R1 |  |
| rs59210467 | 2 |  |  |
| rs11689199 | 2 |  | Arm fat mass (UK Biobank), Arm fat percentage (UK Biobank), Body fat percentage (UK Biobank), Body mass index (UK Biobank), Hip circumference (UK Biobank), Leg fat mass (UK Biobank), Leg fat percentage (UK Biobank), Trunk fat mass (UK Biobank), Trunk fat percentage, Waist circumference (UK Biobank), Weight (UK Biobank), Whole body fat mass (UK Biobank), Years of educational attainment (PMID: 27225129), Cognitive ability multi trait analysis (PMID: 29186694) |
| rs2916251 | 2 |  | Alcohol intake versus 10 years previously (UK Biobank), Body mass index (UK Biobank), Impedance of arm (UK Biobank), Leg fat mass (UK Biobank), Leg fat percentage (UK Biobank), Past tobacco smoking (UK Biobank), Trunk fat mass (UK Biobank), Weight (UK Biobank), Whole body fat mass (UK Biobank) |
| rs1451533 | 2 | PANTR1 | Arm fat mass left (UK Biobank), Arm fat percentage (UK Biobank), Body mass index (UK Biobank), Impedance of leg/whole body (UK Biobank), Leg fat mass (UK Biobank) |
| rs2971640 | 2 |  |  |
| rs12476388 | 2 | DPP10 |  |
| rs374722 | 2 |  |  |
| rs3754970 | 2 | TANK | Fluid intelligence score (UK Biobank), Years of educational attainment (PMID: 27225129), Cognitive ability multi trait analysis (PMID: 29186694), Qualifications-degree (UK Biobank) |
| rs1913808 | 2 | SLC4A10 |  |
| rs2060199 | 2 | SCN2A |  |
| rs263771 | 2 |  |  |
| rs4577309 | 2 | MFSD6 |  |
| rs34224261 | 2 |  | Leg fat mass (UK Biobank), Leg fat percentage (UK Biobank), Qualifications-degree (UK Biobank) |
| rs4675246 | 2 |  |  |
| rs13029509 | 2 | VWC2L | Average weekly champagne plus white wine intake (UK Biobank) |
| rs10932837 | 2 | LOC105373893 |  |
| rs4973576 | 2 | NGEF | Height (UK Biobank) |
| rs138256022 | 3 | VGLL4 |  |
| rs9834970 | 3 |  | Bipolar disorder (PMID: 28072414) |
| rs1352890 | 3 | BSN | High light scatter percentage of red cells (PMID: 27863252), High light scatter reticulocyte count (PMID: 27863252), Immature fraction of reticulocytes (PMID: 27863252), Reticulocyte count (PMID: 27863252), Reticulocyte fraction of red cells (PMID: 27863252), Crohns disease (PMID: 26192919), Inflammatory bowel disease (PMID: 26192919), Ulcerative colitis (PMID: 26192919), Body fat percentage (UK Biobank), Pulse rate (UK Biobank), Qualifications: college or university degree (UK Biobank), Trunk fat percentage (UK Biobank) |
| rs3796386 | 3 | CAMKV | Reticulocyte count (PMID: 27863252), Inflammatory bowel disease (PMID: 26192919), Ulcerative colitis (PMID: 23128233), Age at menarche (PMID: 25231870), Years of educational attainment (PMID: 27225129), Age at first live birth (UK Biobank), Age at menarche (UK Biobank), Arm fat mass (UK Biobank), Arm fat percentage (UK Biobank), Arm fat-free mass (UK Biobank), Arm predicted mass (UK Biobank), Average weekly beer plus cider intake (UK Biobank), Basal metabolic rate (UK Biobank), Body fat percentage (UK Biobank), Body mass index (UK Biobank), Hip circumference (UK Biobank), Impedance of arm/leg/whole body (UK Biobank), Leg fat mass (UK Biobank), Leg fat percentage (UK Biobank), Leg fat-free mass (UK Biobank), Leg predicted mass (UK Biobank), Overall health rating (UK Biobank), Trunk fat mass (UK Biobank), Trunk fat percentage (UK Biobank), Trunk fat-free mass (UK Biobank), Trunk predicted mass (UK Biobank), Waist circumference (UK Biobank), Weight (UK Biobank), Whole body fat mass (UK Biobank), Whole body fat-free mass (UK Biobank), Average weekly beer plus cider intake (UK Biobank), Qualifications-degree (UK Biobank) |
| rs11917871 | 3 | CACNA2D3 |  |
| rs11130793 | 3 | FHIT |  |
| rs11714337 | 3 | FOXP1 | Alcohol intake frequency (UK Biobank), Forced vital capacity (UK Biobank), Educational attainment years of education (PMID: 27225129), Qualifications-degree (UK Biobank) |
| rs12496084 | 3 |  |  |
| rs6797840 | 3 | CADM2 | Arm fat mass (UK Biobank), Arm fat-free mass (UK Biobank), Body fat percentage (UK Biobank), Body mass index (UK Biobank), Hip circumference (UK Biobank), Impedance of arm (UK Biobank), Leg fat mass (UK Biobank), Leg fat percentage (UK Biobank), Qualifications-degree (UK Biobank), Trunk fat mass (UK Biobank), Waist circumference (UK Biobank), Weight (UK Biobank), Whole body fat mass (UK Biobank) |
| rs56398417 | 3 | HTR1F | Arm fat mass (UK Biobank), Arm fat-free mass (UK Biobank), Arm predicted mass (UK Biobank), Basal metabolic rate (UK Biobank), Body mass index (UK Biobank),  Leg fat mass (UK Biobank), Leg fat-free mass (UK Biobank), Leg predicted mass (UK Biobank), Trunk fat mass (UK Biobank), Trunk fat-free mass (UK Biobank), Trunk predicted mass (UK Biobank), Waist circumference (UK Biobank), Weight (UK Biobank), Whole body fat mass (UK Biobank), Whole body fat-free mass (UK Biobank), Whole body water mass (UK Biobank) |
| rs2034768 | 3 |  | Arm fat mass (UK Biobank), Arm fat percentage (UK Biobank), Basal metabolic rate (UK Biobank), Body fat percentage (UK Biobank), Body mass index (UK Biobank), Hip circumference (UK Biobank), Impedance of leg/whole body (UK Biobank), Leg fat mass (UK Biobank), Leg fat percentage (UK Biobank), Leg fat-free mass (UK Biobank), Leg predicted mass (UK Biobank), Trunk fat mass (UK Biobank), Trunk fat percentage (UK Biobank), Waist circumference (UK Biobank), Weight (UK Biobank), Whole body fat mass (UK Biobank),Whole body fat-free mass (UK Biobank) |
| rs12632778 | 3 | LOC105374029 |  |
| rs9867121 | 3 | ZBTB20 |  |
| rs12491503 | 3 |  | Serum butyrylcholinesterase activity (PMID: 21862451) |
| rs114600294 | 3 | SOX2-OT |  |
| rs66852340 | 4 | RGS12 | Impedance of arm (UK Biobank), Qualifications-degree (UK Biobank) |
| rs7700107 | 4 | LCORL | Height (PMID: 25282103), Arm fat percentage (UK Biobank), Arm fat-free mass (UK Biobank), Arm predicted mass (UK Biobank), Basal metabolic rate (UK Biobank), Birth weight (UK Biobank), Body fat percentage (UK Biobank), Forced vital capacity (UK Biobank), Leg fat percentage (UK Biobank), Leg fat-free mass (UK Biobank), Leg predicted mass (UK Biobank), Self-reported hypertension (UK Biobank), Trunk fat-free mass (UK Biobank), Trunk predicted mass (UK Biobank), Weight (UK Biobank), Whole body fat-free mass (UK Biobank), Hand grip strength (UK Biobank) |
| rs34811474 | 4 | ANAPC4 | Age at menarche (UK Biobank), Arm fat mass (UK Biobank), Arm fat percentage (UK Biobank), Body fat percentage (UK Biobank), Body mass index (UK Biobank), Comparative body size at age 10 (UK Biobank), Diastolic blood pressure (UK Biobank), Forced expiratory volume in 1-second (UK Biobank), Forced vital capacity (UK Biobank), Height (UK Biobank), Hip circumference (UK Biobank), Impedance of arm (UK Biobank), Impedance of leg (UK Biobank), Impedance of whole body (UK Biobank), Leg fat mass (UK Biobank), Leg fat percentage right (UK Biobank), Overall health rating (UK Biobank), Qualifications: college or university degree (UK Biobank), Usual walking pace (UK Biobank), Waist circumference (UK Biobank), Weight (UK Biobank), Whole body fat mass (UK Biobank) |
| rs77215114 | 4 |  |  |
| rs28971796 | 4 |  | Hip circumference (UK Biobank), Weight (UK Biobank) |
| rs6850494 | 4 |  | Arm fat-free mass (UK Biobank), Arm predicted mass (UK Biobank), Basal metabolic rate (UK Biobank), Height (UK Biobank), Leg fat-free mass (UK Biobank), Leg predicted mass (UK Biobank), Trunk fat mass (UK Biobank), Trunk fat-free mass (UK Biobank), Trunk predicted mass (UK Biobank), Weight (UK Biobank), Whole body fat-free mass (UK Biobank) |
| rs13107325 | 4 | SLC39A8 | Hematocrit (PMID: 27863252), Childhood BMI (PMID: 26604143), Body mass index (PMID: 29273807), Height (PMID: 28146470), High density lipoprotein (PMID: 24097068), Diastolic blood pressure (PMID: 21909110), Systolic blood pressure (PMID: 21909110), Crohns disease (PMID: 26192919), Schizophrenia (PMID: 26198764), Alcohol intake frequency (UK Biobank), Arm fat mass (UK Biobank), Arm fat percentage (UK Biobank), Fluid intelligence score (UK Biobank), Hip circumference (UK Biobank), Impedance of leg (UK Biobank), Leg fat mass (UK Biobank), Leg fat percentage (UK Biobank), Prospective memory result (UK Biobank), Qualifications-degree (UK Biobank), Trunk fat mass (UK Biobank), Trunk fat percentage (UK Biobank), Usual walking pace (UK Biobank), Waist circumference (UK Biobank), Weight (UK Biobank), Sleep duration (UK Biobank), Overall health rating (UK Biobank) |
| rs62332760 | 4 | TET2 | Lymphocyte count (PMID: 27863252), Lymphocyte percentage of white cells (PMID: 27863252), Neutrophil percentage of white cells (PMID: 27863252), Arm fat-free mass (UK Biobank), Arm predicted mass (UK Biobank), Basal metabolic rate (UK Biobank), Birth weight of first child (UK Biobank), Comparative height size at age 10 (UK Biobank), Fluid intelligence score (UK Biobank), Forced expiratory volume in 1-second (UK Biobank), Forced vital capacity (UK Biobank), Height (UK Biobank), Leg fat-free mass (UK Biobank), Leg predicted mass (UK Biobank), Qualifications: college or university degree (UK Biobank), Trunk fat-free mass (UK Biobank), Trunk predicted mass (UK Biobank), Weight (UK Biobank), Whole body fat-free mass (UK Biobank), Whole body water mass (UK Biobank) |
| rs7693703 | 4 | LINC01378 |  |
| rs4334769 | 4 |  |  |
| rs6825241 | 4 | FAM160A1 | Platelet count (PMID: 27863252), Height (UK Biobank) |
| rs7693082 | 4 | C4orf45 | Overall health rating (UK Biobank), Pulse rate (UK Biobank), Time spent using computer (UK Biobank) |
| rs10940659 | 5 | PDE4D | Qualifications-degree (UK Biobank) |
| rs2409838 | 5 | ERCC8 | Mean platelet volume (PMID: 27863252), Qualifications: college or university degree (UK Biobank), Years of educational attainment (PMID: 27225129) |
| rs262890 | 5 |  | Qualifications-degree (UK Biobank) |
| rs7716447 | 5 |  | Body mass index (UK Biobank), Height (UK Biobank), Impedance of arm/leg/whole body (UK Biobank) |
| rs1031423 | 5 | FAM172A |  |
| rs57585211 | 5 | FBXL17 |  |
| rs9326862 | 5 | APC | Arm fat-free mass (UK Biobank), Arm predicted mass (UK Biobank), Height (UK Biobank), Trunk fat-free mass (UK Biobank), Trunk predicted mass (UK Biobank) |
| rs10041724 | 5 |  |  |
| rs10057545 | 5 | SIL1 |  |
| rs62379379 | 5 |  |  |
| rs10054327 | 5 | HTR4 |  |
| rs42210 | 5 | TENM2 | Qualifications-degree (UK Biobank) |
| rs2336895 | 5 | TENM2 | Current tobacco smoking (UK Biobank), Past tobacco smoking (UK Biobank) |
| rs72828890 | 5 | WWC1 |  |
| rs255292 | 5 | BNIP1 | Height (UK Biobank), Pulse rate (UK Biobank) |
| rs72834698 | 6 |  | Mean corpuscular hemoglobin (PMID: 27863252), Arm fat mass (UK Biobank), Arm fat percentage (UK Biobank), Arm fat-free mass (UK Biobank), Arm predicted mass (UK Biobank), Basal metabolic rate (UK Biobank), Body fat percentage (UK Biobank), Height (UK Biobank), Hip circumference (UK Biobank), Leg fat mass (UK Biobank), Leg fat percentage (UK Biobank), Leg fat-free mass (UK Biobank), Leg predicted mass (UK Biobank), Trunk fat mass (UK Biobank), Trunk fat percentage (UK Biobank), Trunk fat-free mass (UK Biobank), Trunk predicted mass (UK Biobank), Waist circumference (UK Biobank), Weight (UK Biobank), Whole body fat mass (UK Biobank), Whole body fat-free mass (UK Biobank), Qualifications-degree (UK Biobank) |
| rs3135044 | 6 | PRRC2A | Eosinophil count (PMID: 27863252), Granulocyte count (PMID: 27863252), Immature fraction of reticulocytes (PMID: 27863252), Lymphocyte count (PMID: 27863252), Mean corpuscular hemoglobin (PMID: 27863252), Mean corpuscular volume (PMID: 27863252), Monocyte count (PMID: 27863252), Myeloid white cell count (PMID: 27863252), Neutrophil count (PMID: 27863252), Platelet distribution width (PMID: 27863252), Sum basophil neutrophil counts (PMID: 27863252), Sum eosinophil basophil counts (PMID: 27863252), Sum neutrophil eosinophil counts (PMID: 27863252), White blood cell count (PMID: 27863252), IgA deficiency (PMID: 27723758), Primary sclerosing cholangitis (PMID: 27992413), Arm fat percentage (UK Biobank), Arm fat-free mass (UK Biobank), Arm predicted mass (UK Biobank), Basal metabolic rate (UK Biobank), Comparative height size at age 10 (UK Biobank), Forced vital capacity (UK Biobank), Hand grip strength (UK Biobank), Height (UK Biobank), Intestinal malabsorption (UK Biobank), Leg fat percentage (UK Biobank), Leg fat-free mass (UK Biobank), Leg predicted mass (UK Biobank), Overall health rating (UK Biobank), Self-reported high cholesterol (UK Biobank), Self-reported hypertension (UK Biobank), Self-reported hyperthyroidism or thyrotoxicosis (UK Biobank), Self-reported malabsorption or coeliac disease (UK Biobank), Self-reported multiple sclerosis (UK Biobank), Self-reported psoriasis (UK Biobank), Self-reported rheumatoid arthritis (UK Biobank), Trunk fat-free mass (UK Biobank), Trunk predicted mass (UK Biobank), Whole body fat-free mass (UK Biobank), Whole body water mass (UK Biobank), Rheumatoid arthritis (UK Biobank), |
| rs576432906 | 6 |  |  |
| rs10947452 | 6 |  | Platelet count (PMID: 27863252), Plateletcrit (PMID: 27863252) |
| rs9471333 | 6 | LRFN2 | Arm fat mass (UK Biobank), Arm fat percentage (UK Biobank), Arm fat-free mass (UK Biobank), Arm predicted mass (UK Biobank), Basal metabolic rate (UK Biobank), Body fat percentage (UK Biobank), Body mass index (UK Biobank), Hip circumference (UK Biobank), Impedance of arm/whole body (UK Biobank), Leg fat mass (UK Biobank), Leg fat percentage (UK Biobank), Leg fat-free mass (UK Biobank), Leg predicted mass (UK Biobank), Trunk fat mass (UK Biobank), Trunk fat percentage (UK Biobank), Usual walking pace (UK Biobank), Waist circumference (UK Biobank), Weight (UK Biobank), Whole body fat mass (UK Biobank), Whole body fat-free mass (UK Biobank) |
| rs4523073 | 6 |  |  |
| rs77589760 | 6 | PM20D2 |  |
| rs6905544 | 6 |  | Gut microbiota bacterial taxa (PMID: 27694959), Alcohol intake frequency (UK Biobank), Arm fat mass (UK Biobank), Arm fat percentage (UK Biobank), Body fat percentage (UK Biobank), Body mass index (UK Biobank), Fluid intelligence score (UK Biobank), Leg fat mass (UK Biobank), Leg fat percentage (UK Biobank), Overall health rating (UK Biobank), Qualifications-degree (UK Biobank), Sodium in urine (UK Biobank), Time spent using computer (UK Biobank), Trunk fat mass (UK Biobank), Trunk fat percentage (UK Biobank), Waist circumference (UK Biobank), Weight (UK Biobank), Whole body fat mass (UK Biobank), Years of educational attainment (PMID: 27225129) |
| rs17789218 | 6 | LOC105377911 | Diastolic blood pressure (UK Biobank), Hip circumference (UK Biobank), Impedance of leg (UK Biobank), Leg fat-free mass (UK Biobank), Leg predicted mass (UK Biobank), Time spent using computer (UK Biobank), Trunk fat mass (UK Biobank), Weight (UK Biobank), Whole body fat mass (UK Biobank), Arm fat mass (UK Biobank), Basal metabolic rate (UK Biobank), Comparative height size at age 10 (UK Biobank) |
| rs78394231 | 6 | PDSS2 |  |
| rs2184364 | 6 | HIVEP2 | Body fat percentage (UK Biobank), Trunk fat mass (UK Biobank), Trunk fat percentage (UK Biobank) |
| rs9718104 | 6 | PDE10A | Bipolar disorder (UK Biobank) |
| rs17568389 | 6 | GLCCI1 | Height (PMID: 25282103), Trunk fat-free mass (UK Biobank), Trunk predicted mass (UK Biobank), Qualifications-degree (UK Biobank) |
| rs10234444 | 7 | LOC107986788 |  |
| rs6973656 | 7 | TMEM60 | Chronic kidney disease (PMID: 20383146), Glomerular filtration rate in non diabetics creatinine (PMID: 26831199), Serum creatinine (PMID: 20383146), Arm fat mass (UK Biobank), Body fat percentage (UK Biobank), Hip circumference (UK Biobank), Leg fat mass (UK Biobank), Leg fat percentage left (UK Biobank), Leg fat-free mass left (UK Biobank), Trunk fat mass (UK Biobank), Trunk fat percentage (UK Biobank), Weight (UK Biobank), Whole body fat mass (UK Biobank), basal metabolic rate (UK Biobank) |
| rs973734 | 7 |  |  |
| rs3137 | 7 | ZKSCAN5 | Arm fat mass (UK Biobank), Arm fat percentage (UK Biobank), Body fat percentage (UK Biobank), Body mass index (UK Biobank), Height (UK Biobank), Leg fat mass (UK Biobank), Leg fat percentage (UK Biobank), Sitting height (UK Biobank), Waist circumference (UK Biobank), Whole body fat mass (UK Biobank), Dihydroepiandrosterone sulphate (PMID: 28887542) |
| rs2694151 | 7 | CUX1 |  |
| rs62471080 | 7 |  |  |
| rs7788008 | 7 |  | Arm fat mass (UK Biobank), Arm fat percentage (UK Biobank), Body fat percentage (UK Biobank), Body mass index (UK Biobank), Leg fat mass (UK Biobank), Leg fat percentage (UK Biobank), Trunk fat mass (UK Biobank), Weight (UK Biobank) |
| rs10246289 | 7 |  |  |
| rs11763734 | 7 | GRM8 | Neuroticism score (UK Biobank) |
| rs7834121 | 8 | XKR6 | Neuroticism (PMID: 29292387), Granulocyte count (PMID: 27863252), Neutrophil count (PMID: 27863252), Platelet distribution width (PMID: 27863252), Sum basophil neutrophil counts (PMID: 27863252), Sum neutrophil eosinophil counts (PMID: 27863252), White blood cell count (PMID: 27863252), Body mass index (UK Biobank), Diastolic blood pressure (UK Biobank), Heel bone mineral density (UK Biobank), Impedance of arm/leg/whole body (UK Biobank), Systolic blood pressure (UK Biobank), Nervous feelings (UK Biobank) |
| rs1421334 | 8 | PURG | Arm fat mass (UK Biobank), Arm fat percentage (UK Biobank), Body fat percentage (UK Biobank), Body mass index (UK Biobank), Leg fat mass (UK Biobank), Leg fat percentage (UK Biobank), Overall health rating (UK Biobank), Trunk fat mass (UK Biobank), Trunk fat percentage (UK Biobank), Waist circumference (UK Biobank), Whole body fat mass (UK Biobank), Qualifications-degree (UK Biobank) |
| rs6996198 | 8 |  |  |
| rs6472942 | 8 |  | Arm fat mass (UK Biobank), Arm fat percentage (UK Biobank), Body fat percentage (UK Biobank), Body mass index (UK Biobank), Hip circumference (UK Biobank), Leg fat mass (UK Biobank), Leg fat percentage (UK Biobank), Systolic blood pressure (UK Biobank),Trunk fat mass (UK Biobank), Trunk fat percentage (UK Biobank), Waist circumference (UK Biobank),Weight (UK Biobank), Whole body fat mass (UK Biobank), Comparative body size at age 10 (UK Biobank) |
| rs11985394 | 8 |  |  |
| rs72671494 | 8 |  |  |
| rs12541615 | 8 | EXT1 |  |
| rs2717559 | 8 |  |  |
| rs2616830 | 9 | LOC105375951 | Years of educational attainment (PMID: 27225129) |
| rs2291681 | 9 | FREM1 |  |
| rs72671494 | 9 |  |  |
| rs34864022 | 9 |  |  |
| rs12554512 | 9 |  | Years of educational attainment (PMID: 27225129), Bipolar disorder (PMID: 27329760), Basal metabolic rate (UK Biobank), Fluid intelligence score (UK Biobank), Impedance of leg (UK Biobank), Leg fat-free mass (UK Biobank), Leg predicted mass (UK Biobank), Time spent using computer (UK Biobank), Whole body fat-free mass (UK Biobank), Qualifications-degree (UK Biobank), Cognitive ability (UK Biobank) |
| rs2791574 | 9 | TLE4 | Lymphocyte count (PMID: 27863252) |
| rs141184308 | 9 | KIF27 |  |
| rs870151 | 9 |  | Age at menarche (UK Biobank), Arm fat percentage (UK Biobank), Body fat percentage (UK Biobank), Body mass index (UK Biobank), Forced vital capacity (UK Biobank), Leg fat percentage (UK Biobank), Qualifications-degree (UK Biobank), Fibrinogen levels (PMID: 28107422), Trunk fat percentage (UK Biobank), Whole body fat mass (UK Biobank) |
| rs2787374 | 9 | INVS | Hip circumference (UK Biobank) |
| rs10739499 | 9 |  |  |
| rs7043521 | 9 | TTLL11 | Age completed full time education (UK Biobank), Body fat percentage (UK Biobank), Body mass index (UK Biobank), Leg fat mass (UK Biobank), Leg fat percentage (UK Biobank), Qualifications-degree (UK Biobank) |
| rs12342024 | 9 | CRB2 |  |
| rs4382592 | 9 | MED27 |  |
| rs2073869 | 9 | SPACA9 |  |
| rs7469569 | 9 | FUT7 | Lymphocyte count (PMID: 27863252), Monocyte percentage of white cells (PMID: 27863252) |
| rs1623025 | 10 | CELF2 | Qualifications-degree (UK Biobank) |
| rs1243182 | 10 | MLLT10 | Arm fat mass (UK Biobank), Arm fat percentage (UK Biobank), Body fat percentage (UK Biobank), Body mass index (UK Biobank), Creatinine in urine (UK Biobank), Hip circumference (UK Biobank), Leg fat mass (UK Biobank), Leg fat percentage (UK Biobank), Sodium in urine (UK Biobank), Trunk fat mass (UK Biobank), Trunk fat percentage (UK Biobank), Waist circumference (UK Biobank), Weight (UK Biobank), Whole body fat mass (UK Biobank) |
| rs2045147 | 10 | PCDH15 |  |
| rs10994943 | 10 | LINC02625 | Age completed full time education (UK Biobank) |
| rs11201422 | 10 |  |  |
| rs10786658 | 10 | ARMH3 | Years of educational attainment (PMID: 27225129) |
| rs7089973 | 10 | ABLIM1 | Bipolar disorder or attention deficit hyperactivity disorder |
| rs11245482 | 10 | CTBP2 |  |
| rs17727474 | 10 |  |  |
| rs12289262 | 11 | TEAD1 |  |
| rs801733 | 11 | PACS1 | Arm fat mass (UK Biobank), Arm fat percentage (UK Biobank), Arm fat-free mass (UK Biobank), Arm predicted mass (UK Biobank), Basal metabolic rate (UK Biobank), Body fat percentage (UK Biobank), Body mass index (UK Biobank), Hip circumference (UK Biobank), Leg fat mass (UK Biobank), Leg fat percentage (UK Biobank), Trunk fat mass (UK Biobank), Trunk fat percentage (UK Biobank), Waist circumference (UK Biobank), Weight (UK Biobank), Whole body fat mass (UK Biobank) |
| rs116175338 | 11 | STARD10 | Leg fat-free mass (UK Biobank), Leg predicted mass (UK Biobank) |
| rs12272012 | 11 | DLG2 |  |
| rs11020045 | 11 | FAT3 |  |
| rs17207890 | 11 |  | Qualifications-degree (UK Biobank) |
| rs10791546 | 11 | CEP126 |  |
| rs113584870 | 11 | CWF19L2 |  |
| rs328277 | 11 |  |  |
| rs648044 | 11 | ZBTB16 | Non glioblastoma glioma (PMID: 28346443) |
| rs11218575 | 11 | MIR100HG |  |
| rs74802478 | 11 | NTM |  |
| rs4937842 | 11 |  |  |
| rs10772643 | 12 |  | Years of educational attainment (PMID: 27225129) |
| rs74996610 | 12 | SOX5 |  |
| rs10771746 | 12 | IPO8 |  |
| rs10876864 | 12 | IKZF4 | Years of educational attainment (PMID: 27225129), Eosinophil count (PMID: 27863252), Eosinophil percentage of granulocytes (PMID: 27863252), Eosinophil percentage of white cells (PMID: 27863252), Neutrophil percentage of granulocytes (PMID: 27863252), Sum eosinophil basophil counts (PMID: 27863252), Allergic disease (PMID: 29083406), Type 1 diabetes (PMID: 18198356),Arm fat-free mass (UK Biobank), Arm predicted mass left (UK Biobank), Asthma (UK Biobank), Basal metabolic rate (UK Biobank), Body mass index (UK Biobank), Hayfever, allergic rhinitis or eczema (UK Biobank), Impedance of arm/leg/whole body (UK Biobank), Leg fat-free mass (UK Biobank), Leg predicted mass (UK Biobank), Trunk fat-free mass (UK Biobank), Trunk predicted mass (UK Biobank), Whole body fat-free mass (UK Biobank), Qualifications-degree (UK Biobank) |
| rs8756 | 12 | HMGA2 | Birthweight (PMID: 23202124), Height (PMID: 28146470), Hip circumference (PMID: 25673412), Weight (PMID: 23754948), Arm fat-free mass (UK Biobank), Arm predicted mass (UK Biobank), Basal metabolic rate (UK Biobank), Comparative body size at age 10 (UK Biobank), Comparative height size at age 10 (UK Biobank),Forced vital capacity (UK Biobank), Impedance of leg (UK Biobank), Leg fat-free mass (UK Biobank), Leg predicted mass (UK Biobank), Trunk fat mass (UK Biobank), Trunk fat-free mass (UK Biobank), Trunk predicted mass (UK Biobank), Whole body fat-free mass (UK Biobank), Pulse pressure (UK Biobank) |
| rs11179838 | 12 |  |  |
| rs2164744 | 12 | MYO1H | Red cell distribution width (PMID: 27863252) |
| rs2173650 | 12 | TESC |  |
| rs1790135 | 12 | MPHOSPH9 | Lymphocyte count (PMID: 27863252), Lymphocyte percentage of white cells (PMID: 27863252), Neutrophil percentage of white cells (PMID: 27863252), Platelet count (PMID: 27863252), Plateletcrit (PMID: 27863252), Basal metabolic rate (UK Biobank), Comparative height size at age 10 (UK Biobank), Diastolic blood pressure (UK Biobank), Height (UK Biobank), Leg fat-free mass (UK Biobank), Leg predicted mass (UK Biobank), Qualifications: college or university degree (UK Biobank), Trunk fat-free mass (UK Biobank), Trunk predicted mass (UK Biobank), Whole body fat-free mass (UK Biobank), Whole body water mass (UK Biobank), Years of educational attainment (UK Biobank) |
| rs9563168 | 13 |  |  |
| rs9569734 | 13 |  | Arm fat mass (UK Biobank), Basal metabolic rate (UK Biobank), Body mass index (UK Biobank), Impedance of leg/whole body (UK Biobank), Leg fat mass (UK Biobank), Leg fat-free mass (UK Biobank), Leg predicted mass left (UK Biobank), Waist circumference (UK Biobank), Weight (UK Biobank) |
| rs9528023 | 13 |  |  |
| rs56858768 | 13 |  | Body mass index (UK Biobank) |
| rs553397860 | 13 | FARP1 |  |
| rs7991062 | 13 |  |  |
| rs11629120 | 14 | C14orf93 | Red cell distribution width (PMID: 27863252), Reticulocyte fraction of red cells (PMID: 27863252), Systolic blood pressure (UK Biobank), Years of educational attainment (PMID: 27225129) |
| rs405797 | 14 | NOVA1 | Qualifications-degree (UK Biobank) |
| rs1022785 | 14 | LOC107984685 | Qualifications-degree (UK Biobank) |
| rs10145592 | 14 |  |  |
| rs7157001 | 14 | LOC105370660 |  |
| rs8043253 | 14 | EXD1 | Diastolic blood pressure (UK Biobank), Hayfever, allergic rhinitis or eczema (UK Biobank), Height (UK Biobank), Self-reported hypertension (UK Biobank), Systolic blood pressure (UK Biobank), Age at menopause (PMID: 26414677) |
| rs2460 | 15 | ONECUT1 |  |
| rs4775373 | 15 | RORA |  |
| rs61331678 | 15 | LINGO1 | Miserableness (UK Biobank), Neuroticism score (UK Biobank), Years of educational attainment (PMID: 27225129), Qualifications-degree (UK Biobank) |
| rs35574015 | 16 | PDXDC1 | Arm fat-free mass (UK Biobank), Arm predicted mass (UK Biobank), Basal metabolic rate (UK Biobank), Height (UK Biobank), Leg fat-free mass (UK Biobank), Leg predicted mass (UK Biobank), Trunk fat-free mass (UK Biobank), Trunk predicted mass (UK Biobank), Weight (UK Biobank), Whole body fat-free mass (UK Biobank), Arachidonic acid 20:4n6 (PMID: 24823311), Dihomo-gamma-linolenic acid (PMID: 24823311), Gamma-linolenic acid 18:3n6 (PMID: 24823311), Linoleic acid 18:2n6 (PMID: 24823311), Plasma omega 6 polyunsaturated fatty acid levels linoleic acid (PMID: 24823311), Blood metabolite ratios (PMID: 24816252) |
| rs7189927 | 16 | ATP2A1 | Red blood cell count (PMID: 27863252), Body mass index (PMID: 25673413), Hip circumference (PMID: 25673412), Waist circumference (PMID: 25673412), Weight (PMID: 23754948), Alcohol intake frequency (UK Biobank), Arm fat mass (UK Biobank), Arm fat percentage (UK Biobank), Arm fat-free mass left (UK Biobank), Arm predicted mass left (UK Biobank), Basal metabolic rate (UK Biobank), Body fat percentage (UK Biobank), Hip circumference (UK Biobank), Impedance of leg (UK Biobank), Leg fat mass (UK Biobank), Leg fat percentage (UK Biobank), Leg fat-free mass (UK Biobank), Leg predicted mass (UK Biobank), Trunk fat mass (UK Biobank), Trunk fat percentage (UK Biobank), Whole body fat mass (UK Biobank), Whole body fat-free mass (UK Biobank), Qualifications-degree (UK Biobank) |
| rs749671 | 16 | ZNF646 | Lymphocyte count (PMID: 27863252), Body mass index (PMID: 25673413), Hip circumference (PMID: 25673412), Waist circumference (PMID: 25673412), Triglycerides (PMID: 24097068), Arm fat mass (UK Biobank), Arm fat percentage (UK Biobank), Arm fat-free mass (UK Biobank), Arm predicted mass (UK Biobank), Basal metabolic rate (UK Biobank), Body fat percentage (UK Biobank), Body mass index (UK Biobank), Impedance of arm/leg/whole body (UK Biobank), Leg fat mass (UK Biobank), Leg fat percentage (UK Biobank), Leg fat-free mass (UK Biobank), Leg predicted mass (UK Biobank), Overall health rating (UK Biobank), Trunk fat mass (UK Biobank), Trunk fat percentage (UK Biobank), Trunk fat-free mass (UK Biobank), Trunk predicted mass (UK Biobank), Weight (UK Biobank), Whole body fat mass (UK Biobank), Whole body fat-free mass (UK Biobank), Alcohol intake frequency (UK Biobank) |
| rs7184800 | 16 | RBL2 | Arm fat-free mass (UK Biobank), Arm predicted mass (UK Biobank), Basal metabolic rate (UK Biobank), Fluid intelligence score (UK Biobank), Height (UK Biobank), Hip circumference (UK Biobank), Impedance of leg (UK Biobank), Leg fat-free mass (UK Biobank), Leg predicted mass (UK Biobank), Pulse rate (UK Biobank), Trunk fat-free mass (UK Biobank), Trunk predicted mass (UK Biobank), Weight (UK Biobank), Whole body fat-free mass (UK Biobank), Qualifications-degree (UK Biobank) |
| rs142710267 | 16 | PMFBP1 |  |
| rs2447098 | 17 | SGSM2 | Plateletcrit (PMID: 27863252) |
| rs11654952 | 17 |  |  |
| rs2584597 | 17 | TCAM1P |  |
| rs9902312 | 17 | HELZ |  |
| rs11657730 | 17 | BAHCC1 | Height (UK Biobank), Time spent using computer (UK Biobank), Forced vital capacity (UK Biobank), Qualifications-degree (UK Biobank) |
| rs303753 | 18 |  | Years of educational attainment (PMID: 27225129), Granulocyte count (PMID: 27863252), Myeloid white cell count (PMID: 27863252), Neutrophil count (PMID: 27863252), Alcohol intake frequency (UK Biobank), Arm fat mass (UK Biobank), Arm fat percentage (UK Biobank), Arm fat-free mass (UK Biobank), Arm predicted mass (UK Biobank), Basal metabolic rate (UK Biobank), Body fat percentage (UK Biobank), Body mass index (UK Biobank), Hip circumference (UK Biobank), Leg fat mass (UK Biobank), Leg fat percentage (UK Biobank), Leg fat-free mass (UK Biobank), Leg predicted mass (UK Biobank), Trunk fat mass (UK Biobank), Trunk fat percentage (UK Biobank), Trunk fat-free mass (UK Biobank), Trunk predicted mass (UK Biobank), Waist circumference (UK Biobank), Weight (UK Biobank), Whole body fat mass (UK Biobank), Whole body fat-free mass (UK Biobank), Qualifications-degree (UK Biobank) |
| rs71362609 | 18 |  |  |
| rs11081851 | 18 | NOL4 |  |
| rs9964724 |  |  | Educational attainment years of education (PMID: 27225129), Mood swings (UK Biobank), Neuroticism score (UK Biobank), Overall health rating (UK Biobank), Worrier or anxious feelings (UK Biobank), Qualifications-degree (UK Biobank) |
| rs1156541 | 18 | LINC00907 |  |
| rs17512836 | 18 | TCF4 | Schizophrenia (PMID: 21926974) |
| rs7248205 | 19 | ILF3 | Platelet count (PMID: 27863252), Arm fat-free mass (UK Biobank), Arm predicted mass (UK Biobank), Basal metabolic rate (UK Biobank), Height (UK Biobank), Hip circumference (UK Biobank), Leg fat-free mass (UK Biobank), Leg predicted mass (UK Biobank), Trunk fat-free mass (UK Biobank), Trunk predicted mass (UK Biobank), Weight (UK Biobank), Whole body fat-free mass (UK Biobank) |
| rs111901094 | 19 | GATAD2A | Low density lipoprotein (PMID: 24097068),Total cholesterol (PMID: 24097068), Triglycerides (PMID: 24097068), Plateletcrit (PMID: 27863252), Age at first live birth (UK Biobank), Height (UK Biobank), Leg fat percentage (UK Biobank), Leg fat-free mass (UK Biobank), Leg predicted mass (UK Biobank) |
| rs6131281 | 20 | BTBD3 |  |
| rs6141814 | 20 | DNMT3B |  |
| rs4810312 | 20 | LPIN3 | Low density lipoprotein (PMID: 24097068), Total cholesterol (PMID: 24097068) |
| rs6124476 | 20 | PTPRT |  |
| rs6031440 | 20 | JPH2 | Granulocyte percentage of myeloid white cells (PMID: 27863252), Monocyte percentage of white cells (PMID: 27863252) |
| rs55700114 | 20 |  | Qualifications-degree (UK Biobank) |
| rs6125906 | 20 | UBE2V1 |  |
| rs56103247 | 20 |  | Leg fat percentage (UK Biobank) |
| rs10427502 | 21 | BRWD1 | Age at menarche (UK Biobank) |
| rs7609 | 22 | NIPSNAP1 |  |
| rs2092829 | 22 |  |  |
| rs150399729 | 22 | RRP7BP |  |
| Abbreviations: Chr, chromosome; PMID, Pubmed ID; SNP, single nucleotide polymorphism | | | |

| **Supplemental Table 25: Mendelian Randomization estimates for the effects of sedentary behaviour on the secondary traits** | | | | | | | | | | |
| --- | --- | --- | --- | --- | --- | --- | --- | --- | --- | --- |
| **Methods** | **Leisure television watching** | | | | | **Leisure computer use** | | | | |
|  | **Estimates** | **95% CI** | **P-value** |  | **P-value for pleiotropy^†^ or heterogeneity^‡^** | **Estimates (OR)** | **95% CI** | **P-value** |  | **P-value for pleiotropy^†^ or heterogeneity^‡^** |
| **Education** |  |  |  |  |  |  |  |  |  |  |
| Inverse-variance weighted | -0.54 | -0.58, -0.49 | 2×10^-125^ |  | 2×10^-156^ | 0.59 | 0.48, 0.70 | 7×10^-26^ |  | 4×10^-83^ |
| MR-Egger | -0.56 | -0.75, -0.36 | 2×10^-8^ |  | 0.83 | 1.02 | 0.32, 1.72 | 0.004 |  |  |
| Weighted median | -0.52 | -0.55, -0.48 | 7×10^-169^ |  |  | 0.59 | 0.51, 0.67 | 9×10^-46^ |  |  |
| Inverse-variance weighted* | -0.38 | -0.41, -0.34 | 8×10^-112^ |  | 9×10^-21^ | 0.33 | 0.25, 0.41 | 3×10^-17^ |  | 3×10^-8^ |
| **BMI** |  |  |  |  |  |  |  |  |  |  |
| Inverse-variance weighted | 0.38 | 0.32, 0.45 | 2×10^-31^ |  | 0 | 0 | -0.13, 0.12 | 0.96 |  | 5×10^-105^ |
| MR-Egger | 0.42 | 0.13, 0.72 | 0.005 |  | 0.79 | -0.09 | -0.88, 0.70 | 0.82 |  | 0.82 |
| Weighted median | 0.37 | 0.33, 0.42 | 8×10^-69^ |  |  | 0.02 | -0.06, 0.09 | 0.65 |  |  |
| Inverse-variance weighted* | 0.23 | 0.18, 0.28 | 8×10^-23^ |  | 6×10^-78^ | 0.02 | -0.08, 0.12 | 0.70 |  | 7×10^-37^ |
| **Smoking** |  |  |  |  |  |  |  |  |  |  |
| Inverse-variance weighted | 0.17 | 0.14, 0.20 | 2×10^-33^ |  | 2×10^-69^ | -0.06 | -0.12, -0.01 | 0.02 |  | 2×10^-17^ |
| MR-Egger | 0.10 | -0.03, 0.22 | 0.12 |  | 0.26 | -0.22 | -0.55, 0.12 | 0.21 |  | 0.37 |
| Weighted median | 0.16 | 0.14, 0.19 | 1×10^-38^ |  |  | -0.08 | -0.12, -0.03 | 0.002 |  |  |
| Inverse-variance weighted* | 0.15 | 0.13, 0.18 | 1×10^-31^ |  | 3×10^-45^ |  |  |  |  |  |
| **Alcohol consumption** |  |  |  |  |  |  |  |  |  |  |
| Inverse-variance weighted | -0.02 | -0.05, 0.02 | 0.39 |  | 1×10^-55^ | 0.00 | -0.07, 0.07 | 0.90 |  | 8×10^-14^ |
| MR-Egger | -0.13 | -0.28, 0.03 | 0.12 |  | 0.17 | 0.00 | -0.44, 0.44 | 1.00 |  | 0.98 |
| Weighted median | 0.00 | -0.03, 0.04 | 0.97 |  |  | -0.02 | -0.08, 0.04 | 0.57 |  |  |
| Inverse-variance weighted* | 0.00 | -0.04, 0.03 | 0.79 |  | 4×10^-37^ | 0.01 | -0.05, 0.08 | 0.72 |  | 4×10^-9^ |
| Abbreviations: CI, confidence intervals; MR: Mendelian Randomization; OR: odds ratio; MR-PRESSO: MR pleiotropy residual sum and outlier test  * Excluding leisure television watching/leisure computer use SNPs which are genome-wide significant for the secondary trait  † P-value or pleiotropy based on MR-Egger intercept  ‡ P-value for heterogeneity based on Q statistic | | | | | | | | | | |

| **Supplemental Table 26: Mendelian Randomization estimates for the effects of the secondary traits on sedentary behaviour** | | | | | | | | | | |
| --- | --- | --- | --- | --- | --- | --- | --- | --- | --- | --- |
| **Methods** | **Leisure television watching** | | | | | **Leisure computer use** | | | | |
|  | **Estimates** | **95% CI** | **P-value** |  | **P-value for pleiotropy^†^ or heterogeneity^‡^** | **Estimates (OR)** | **95% CI** | **P-value** |  | **P-value for pleiotropy^†^ or heterogeneity^‡^** |
| **Education** |  |  |  |  |  |  |  |  |  |  |
| Inverse-variance weighted | -0.63 | -0.66, -0.59 | 2×10^-277^ |  | 2×10^-81^ | 0.34 | 0.30, 0.37 | 2×10^-83^ |  | 5×10^-70^ |
| MR-Egger | -0.64 | -0.77, -0.52 | 0 |  | 0.77 | 0.42 | 0.30, 0.54 | 2×10^-11^ |  | 0.17 |
| Weighted median | -0.63 | -0.67, -0.60 | 4×10^-249^ |  |  | 0.34 | 0.30, 0.37 | 1×10^-76^ |  |  |
| Inverse-variance weighted* | -0.55 | -0.58, -0.52 | 8×10^-244^ |  | 4×10^-35^ | 0.30 | 0.27, 0.33 | 8×10^-75^ |  | 5×10^-45^ |
| **BMI** |  |  |  |  |  |  |  |  |  |  |
| Inverse-variance weighted | 0.23 | 0.21, 0.26 | 3×10^-63^ |  | 1×10^-256^ | 0.02 | -0.01, 0.04 | 0.14 |  | 5×10^-80^ |
| MR-Egger | 0.09 | 0.02, 0.16 | 0.02 |  | 5×10^-6^ | 0.01 | -0.04, 0.07 | 0.62 |  | 0.94 |
| Weighted median | 0.20 | 0.18, 0.23 | 4×10^-54^ |  |  | 0.01 | -0.01, 0.03 | 0.44 |  |  |
| Inverse-variance weighted* | 0.20 | 0.17, 0.22 | 1×10^-57^ |  | 4×10^-134^ | 0.02 | 0.00, 0.04 | 0.08 |  | 3×10^-72^ |
| **Smoking** |  |  |  |  |  |  |  |  |  |  |
| Inverse-variance weighted | 0.49 | 0.40, 0.57 | 6×10^-29^ |  | 4×10^-54^ | -0.10 | -0.17, -0.02 | 0.01 |  | 1×10^-29^ |
| MR-Egger | -0.17 | -0.49, 0.15 | 0.29 |  | 3×10^-5^ | 0.08 | -0.21, 0.37 | 0.60 |  | 0.22 |
| Weighted median | 0.52 | 0.44, 0.60 | 3×10^-38^ |  |  | -0.11 | -0.18, -0.04 | 0.004 |  |  |
| Inverse-variance weighted* | 0.45 | 0.37, 0.53 | 2×10^-28^ |  | 1×10^-38^ |  |  |  |  |  |
| **Alcohol consumption** |  |  |  |  |  |  |  |  |  |  |
| Inverse-variance weighted | 0.02 | -0.07, 0.10 | 0.67 |  | 7×10^-50^ | -0.09 | -0.17, -0.01 | 0.03 |  | 2×10^-38^ |
| MR-Egger | 0.07 | -0.09, 0.24 | 0.39 |  | 0.46 | -0.19 | -0.34, -0.03 | 0.02 |  | 0.15 |
| Weighted median | 0.14 | 0.07, 0.21 | 8×10^-4^ |  |  | -0.16 | -0.24, -0.09 | 1×10^-5^ |  |  |
| Inverse-variance weighted* | 0.04 | -0.02, 0.27 | 0.09 |  | 5×10^-31^ |  |  |  |  |  |
| Abbreviations: CI, confidence intervals; MR: Mendelian Randomization; OR: odds ratio; MR-PRESSO: MR pleiotropy residual sum and outlier test  * Excluding secondary traits related SNPs which are genome-wide significant for leisure television watching/leisure computer use  † P-value or pleiotropy based on MR-Egger intercept  ‡ P-value for heterogeneity based on Q statistic | | | | | | | | | | |

| **Supplemental Table 27: Multivariable Mendelian randomization estimates for years of education and cancer risk** | | | | | | |
| --- | --- | --- | --- | --- | --- | --- |
|  | **Years of education** | | | | | |
|  | **Leisure television watching analysis** | | |  | **Leisure computer use analysis** | |
|  | **Estimates (OR)** | **95% CI** |  | | **Estimates (OR)** | **95% CI** |
| **Breast cancer** | 0.92 | 0.78, 1.09 |  | | 0.89 | 0.79, 0.99 |
| **Luminal A breast cancer** | 0.90 | 0.72, 1.11 |  | | 0.81 | 0.70, 0.94 |
| **Luminal B breast cancer** | 0.84 | 0.58, 1.20 |  | | 1.11 | 0.86, 1.45 |
| **Luminal B HER2 negative breast cancer** | 0.92 | 0.68, 1.27 |  | | 0.81 | 0.66, 1.01 |
| **HER2 enriched breast cancer** | 0.76 | 0.44, 1.28 |  | | 0.82 | 0.57, 1.17 |
| **Triple negative breast cancer** | 0.81 | 0.61, 1.08 |  | | 0.90 | 0.73, 1.11 |
| **Colorectal cancer** | 0.73 | 0.60, 0.90 |  | | 0.71 | 0.61, 0.82 |
| **Colorectal cancer in men** | 0.67 | 0.52, 0.88 |  | | 0.68 | 0.56, 0.82 |
| **Colorectal cancer in women** | 0.80 | 0.61, 1.05 |  | | 0.74 | 0.61, 0.89 |
| **Colon cancer** | 0.71 | 0.57, 0.90 |  | | 0.70 | 0.59, 0.83 |
| **Rectal cancer** | 0.59 | 0.43, 0.82 |  | | 0.61 | 0.49, 0.76 |
| Abbreviations: CI, confidence intervals; OR: odds ratio | | | | | | |

**Supplemental Figure 1**: Scatter plots showing the correlation of genetic associations of leisure television watching with genetic associations with breast cancer. Coloured lines represent the slopes of the different regression analyses.

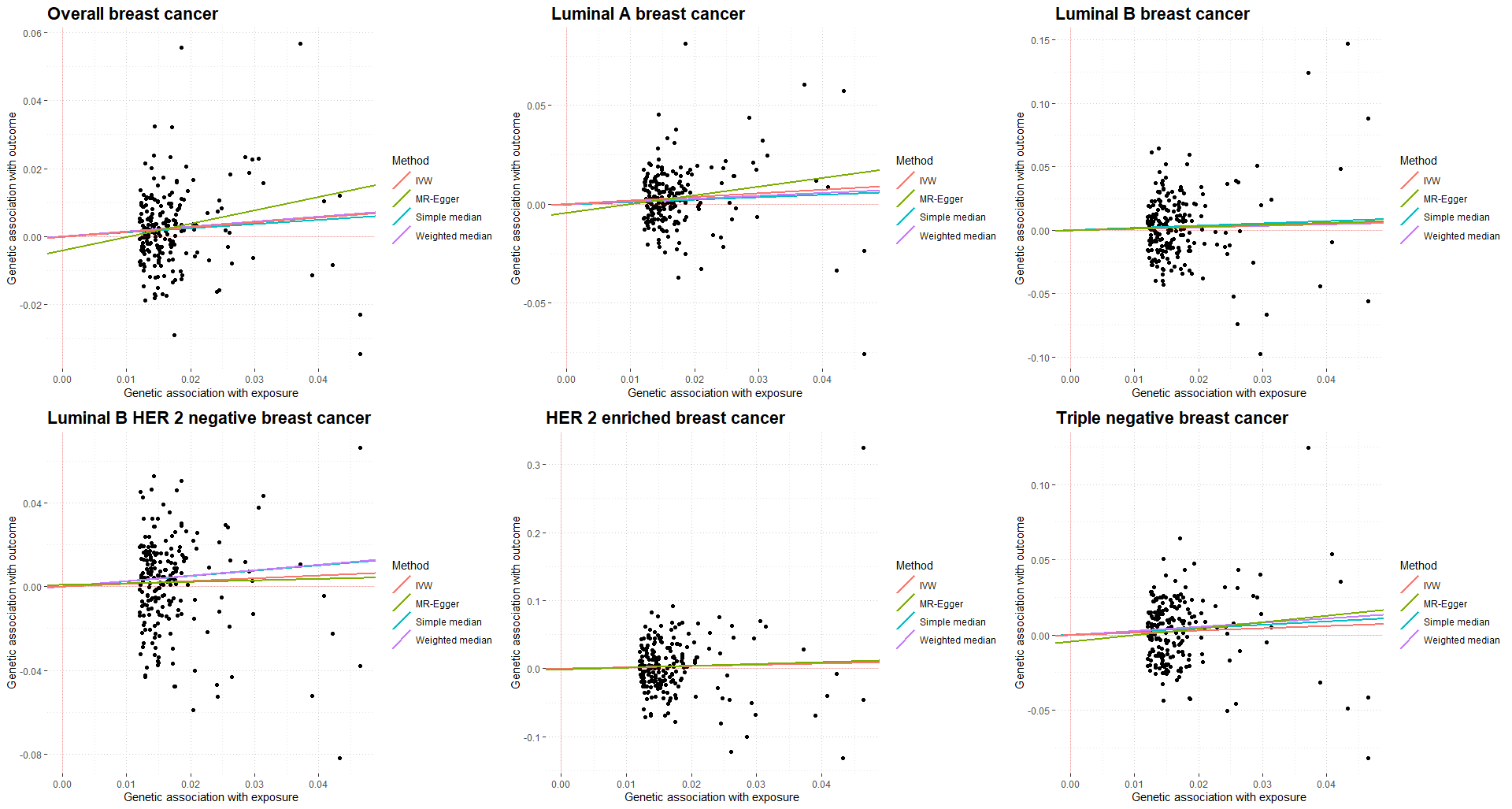

**Supplemental Figure 2**: Scatter plots showing the correlation of genetic associations of leisure television watching with genetic associations with colorectal cancer. Coloured lines represent the slopes of the different regression analyses.

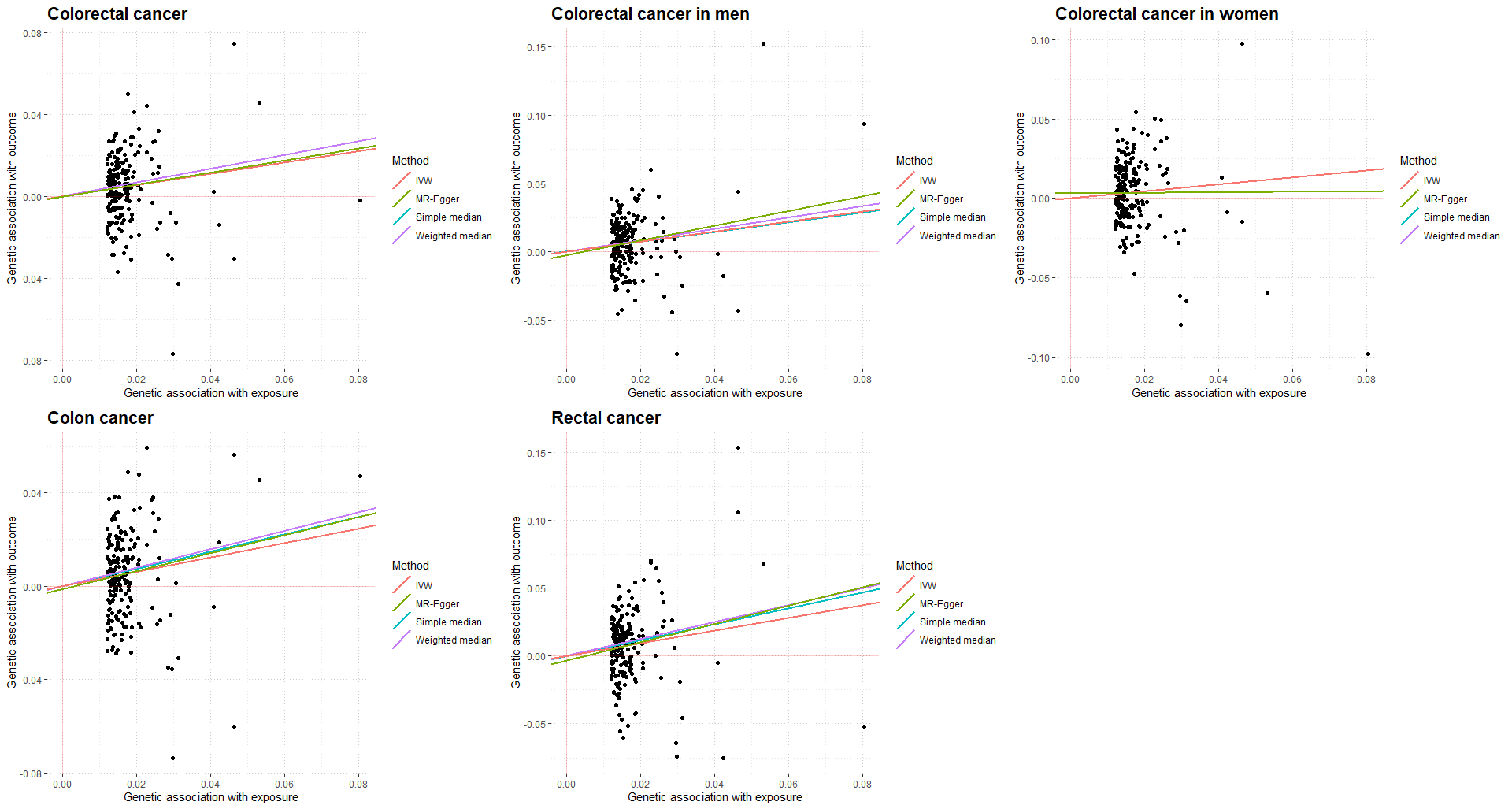

**Supplemental Figure 3**: Scatter plots showing the correlation of genetic associations of leisure television watching with genetic associations with prostate cancer. Coloured lines represent the slopes of the different regression analyses.

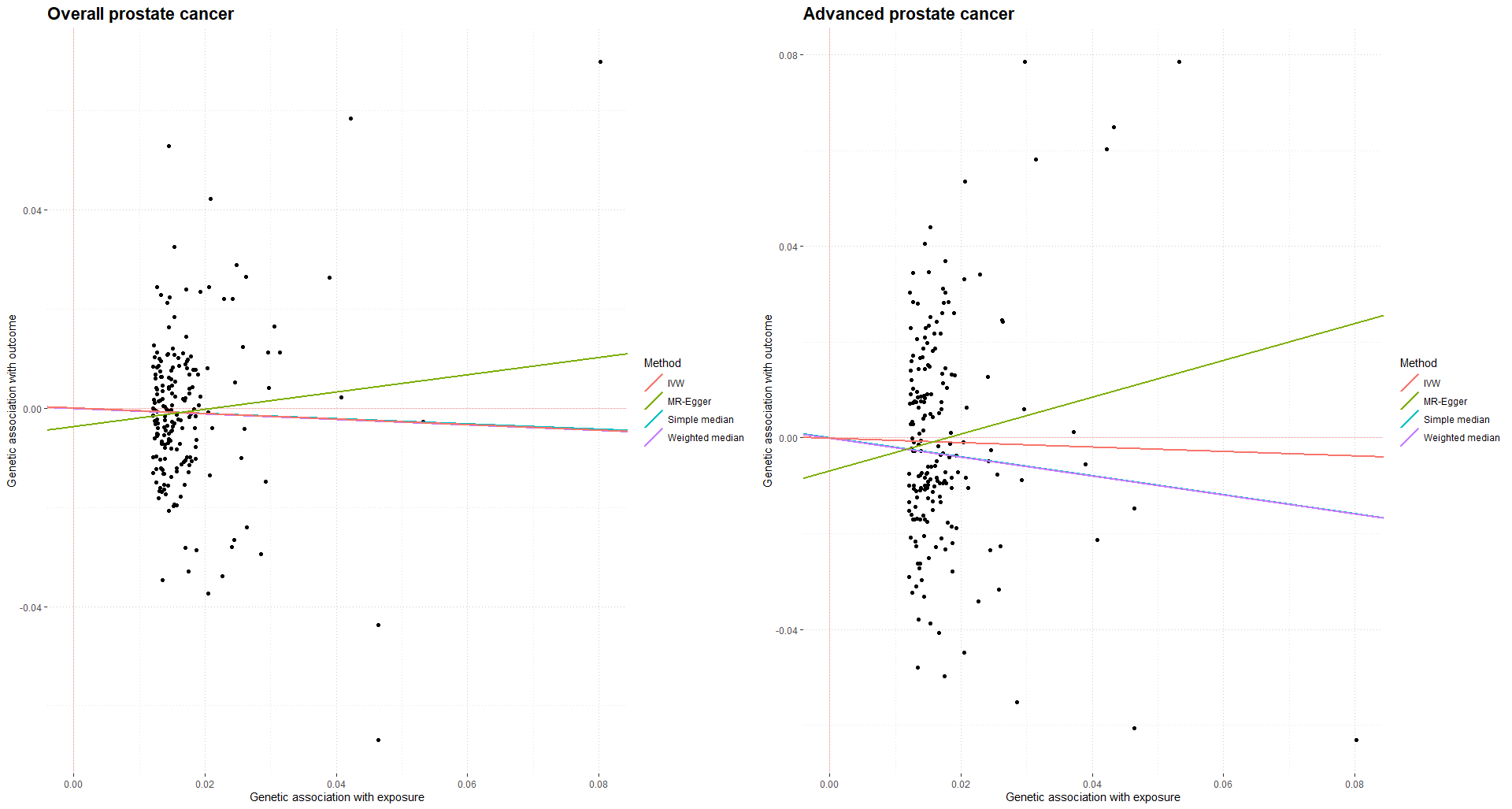

**Supplemental Figure 4**: Funnel plots of risk estimates of leisure television watching and breast cancer against instrumental strength

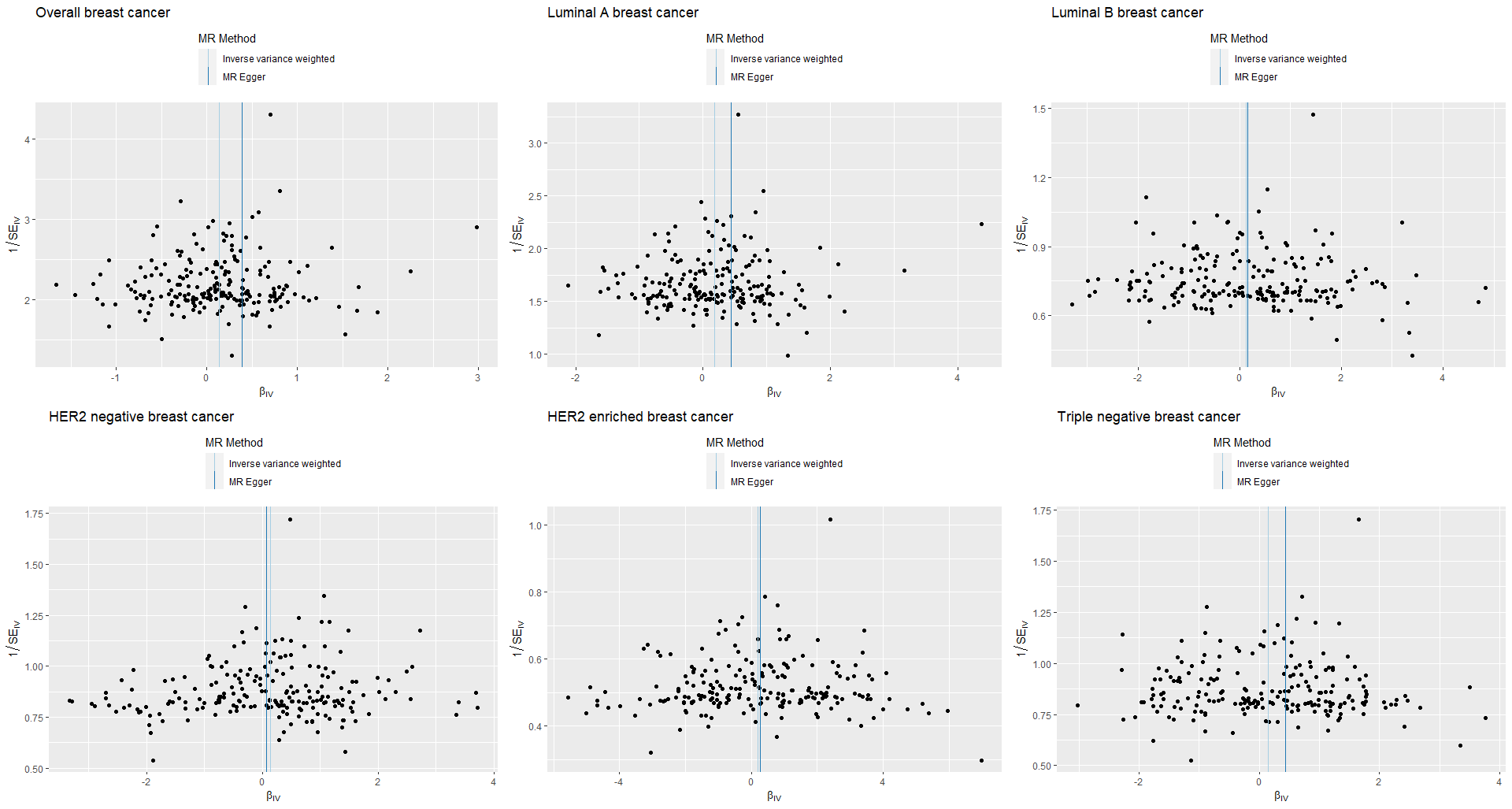

**Supplemental Figure 5**: Funnel plots of risk estimates of leisure television watching and colorectal cancer against instrumental strength

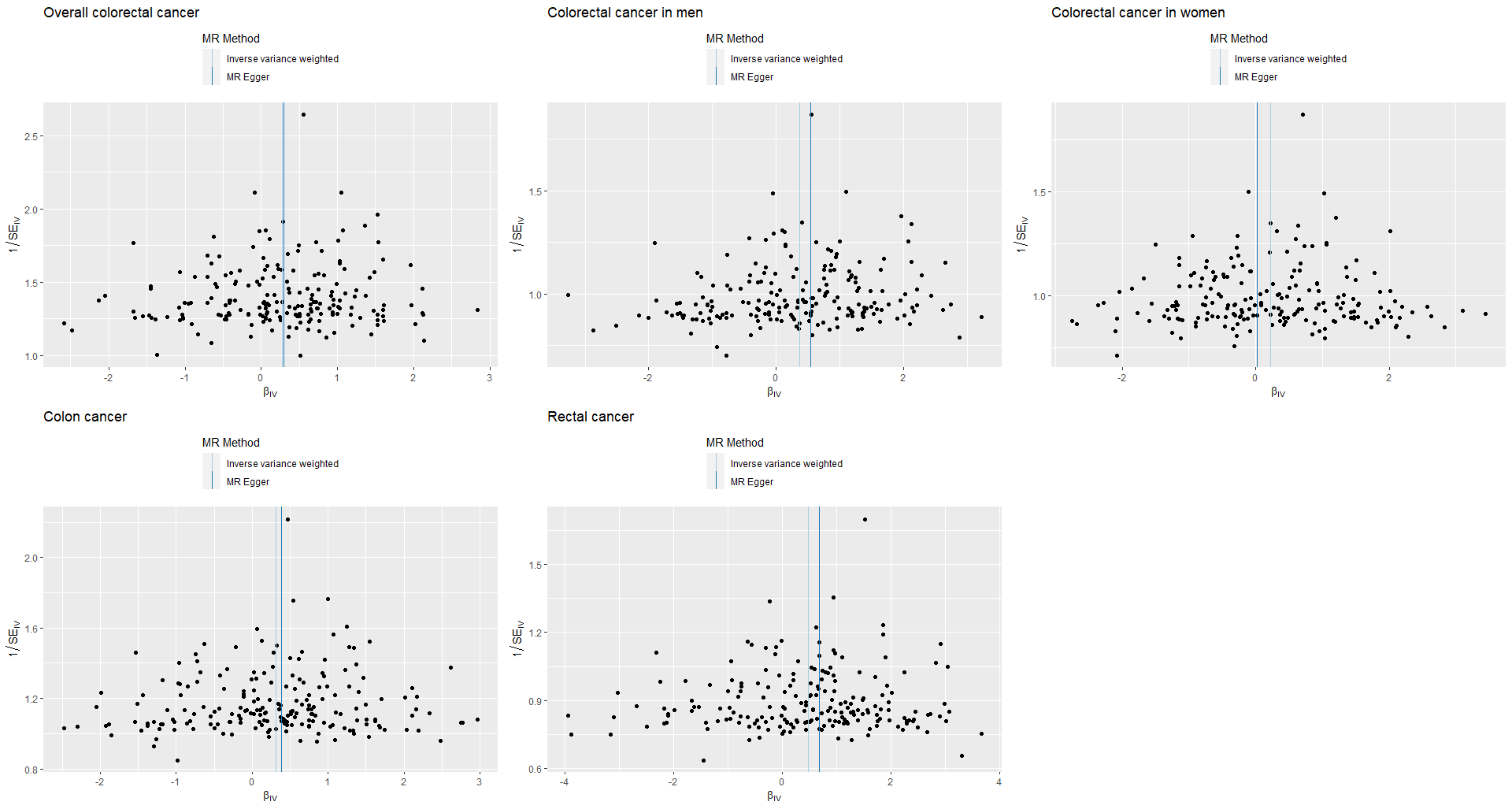

**Supplemental Figure 6**: Funnel plots of risk estimates of leisure television watching and prostate cancer against instrumental strength

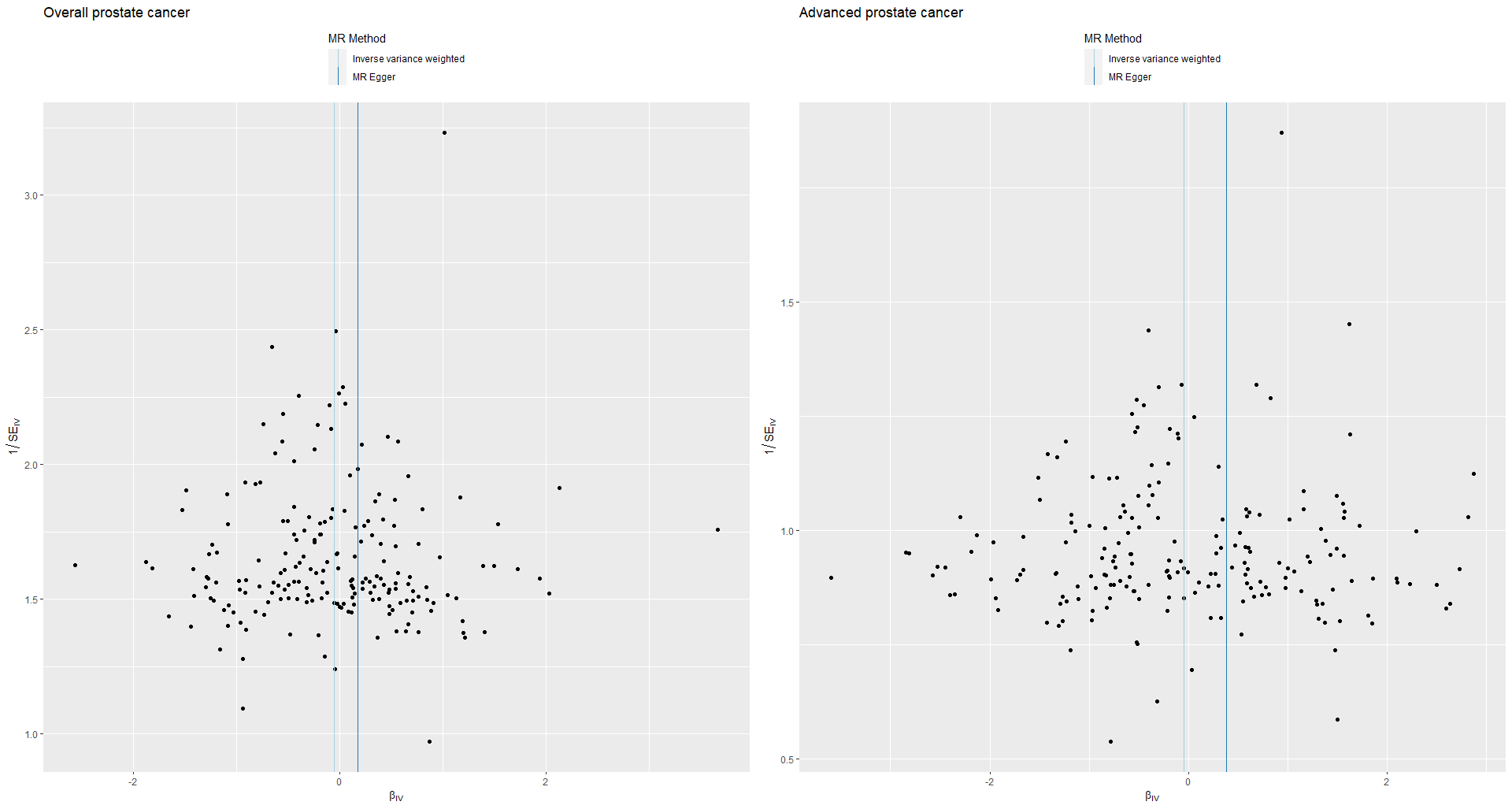

**Supplemental Figure 7**: Scatter plots showing the correlation of genetic associations of leisure computer use with genetic associations with breast cancer. Coloured lines represent the slopes of the different regression analyses.

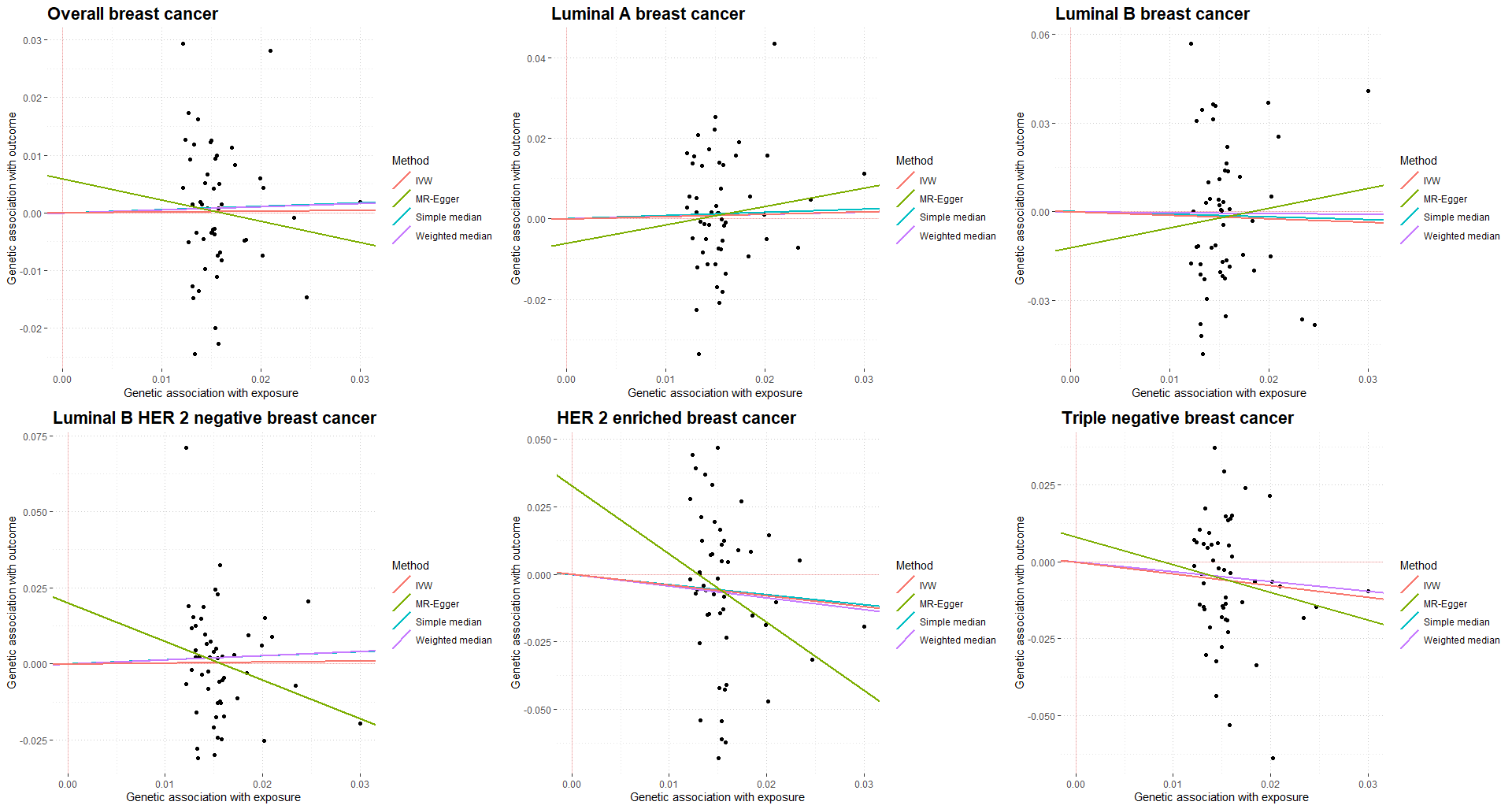

**Supplemental Figure 8**: Scatter plots showing the correlation of genetic associations of leisure computer use with genetic associations with colorectal cancer. Coloured lines represent the slopes of the different regression analyses.

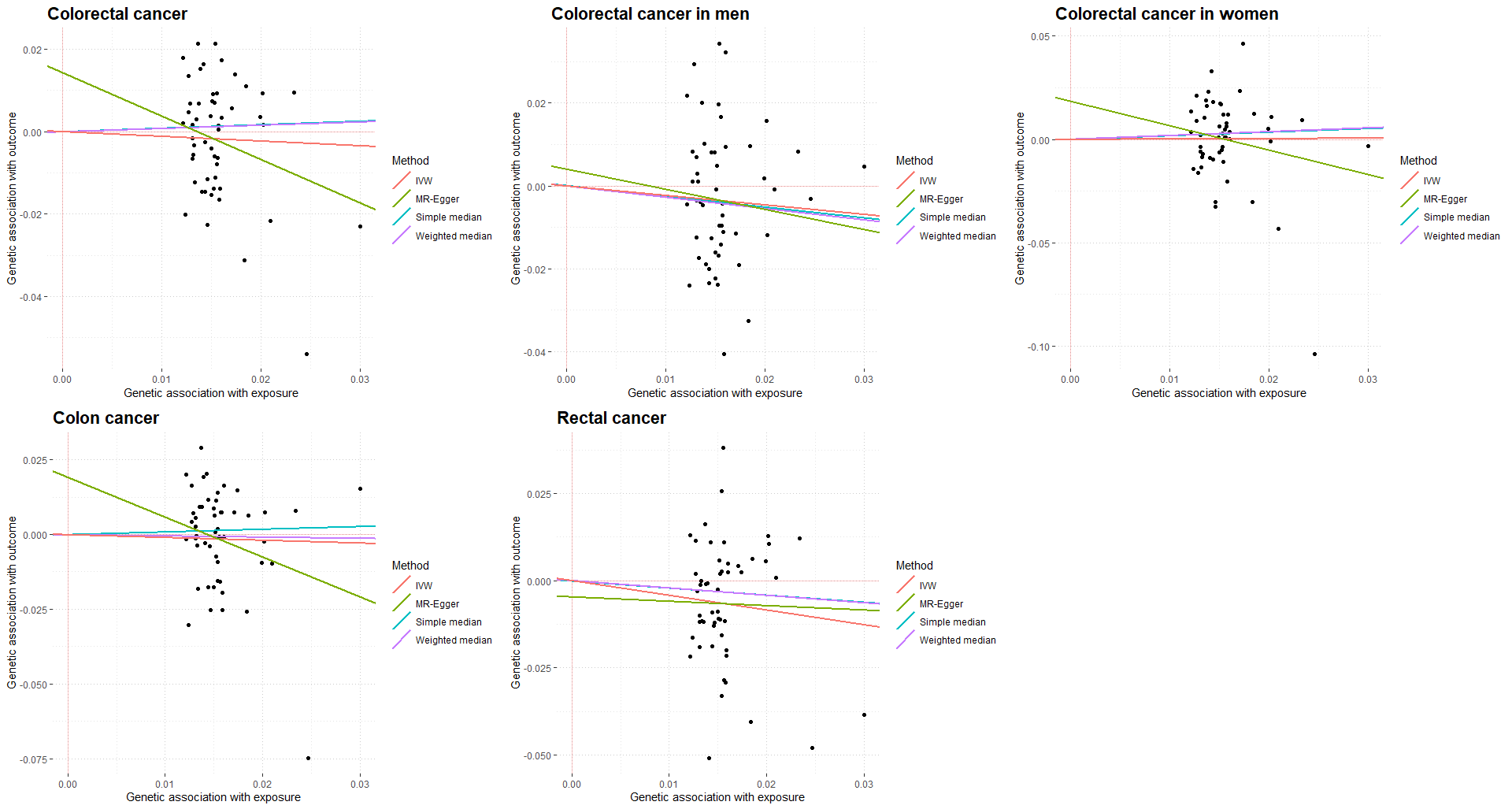

**Supplemental Figure 9**: Scatter plots showing the correlation of genetic associations of leisure computer use with genetic associations with prostate cancer. Coloured lines represent the slopes of the different regression analyses

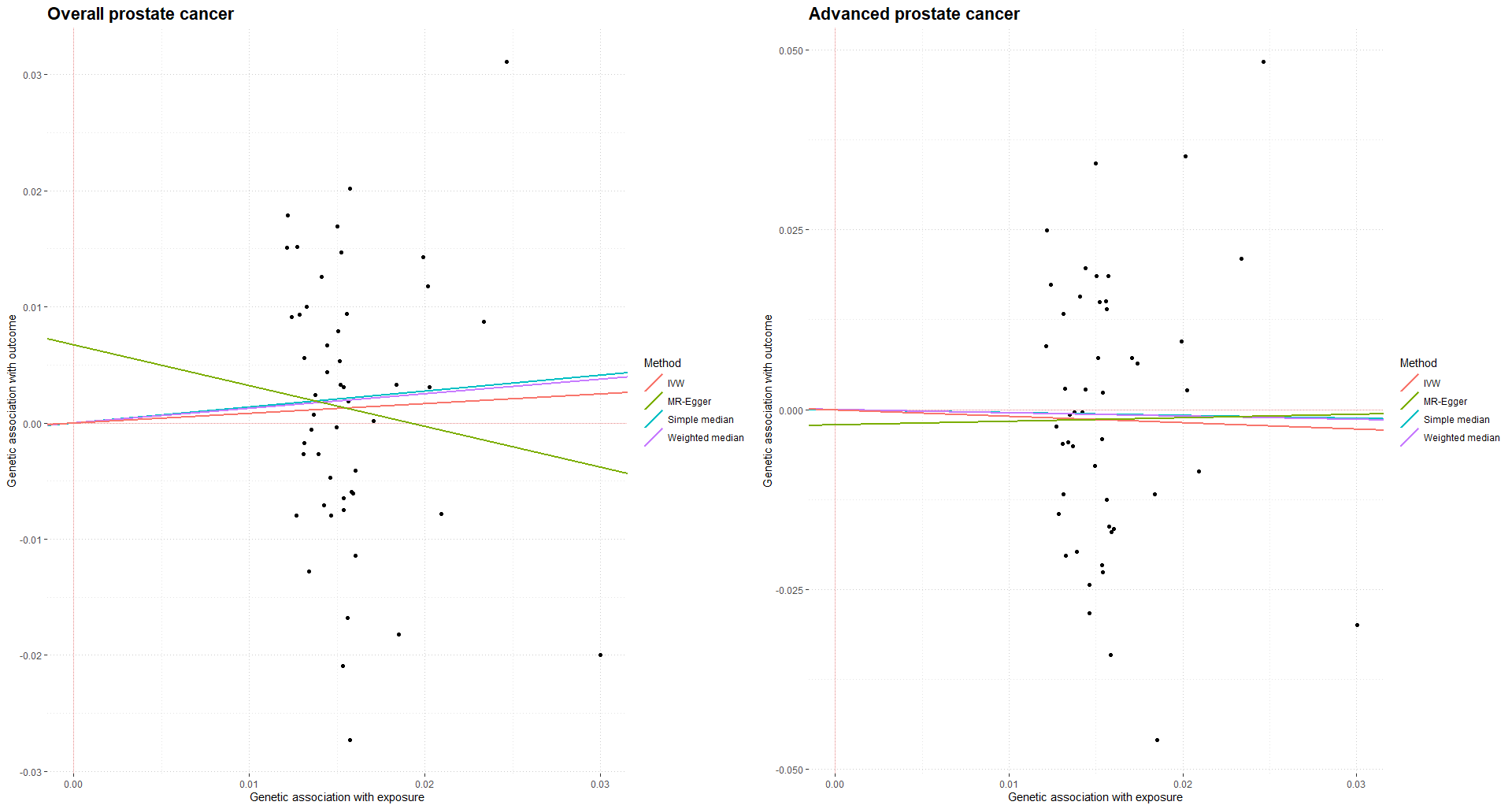

**Supplemental Figure 10**: Funnel plots of risk estimates of leisure computer use and breast cancer against instrumental strength

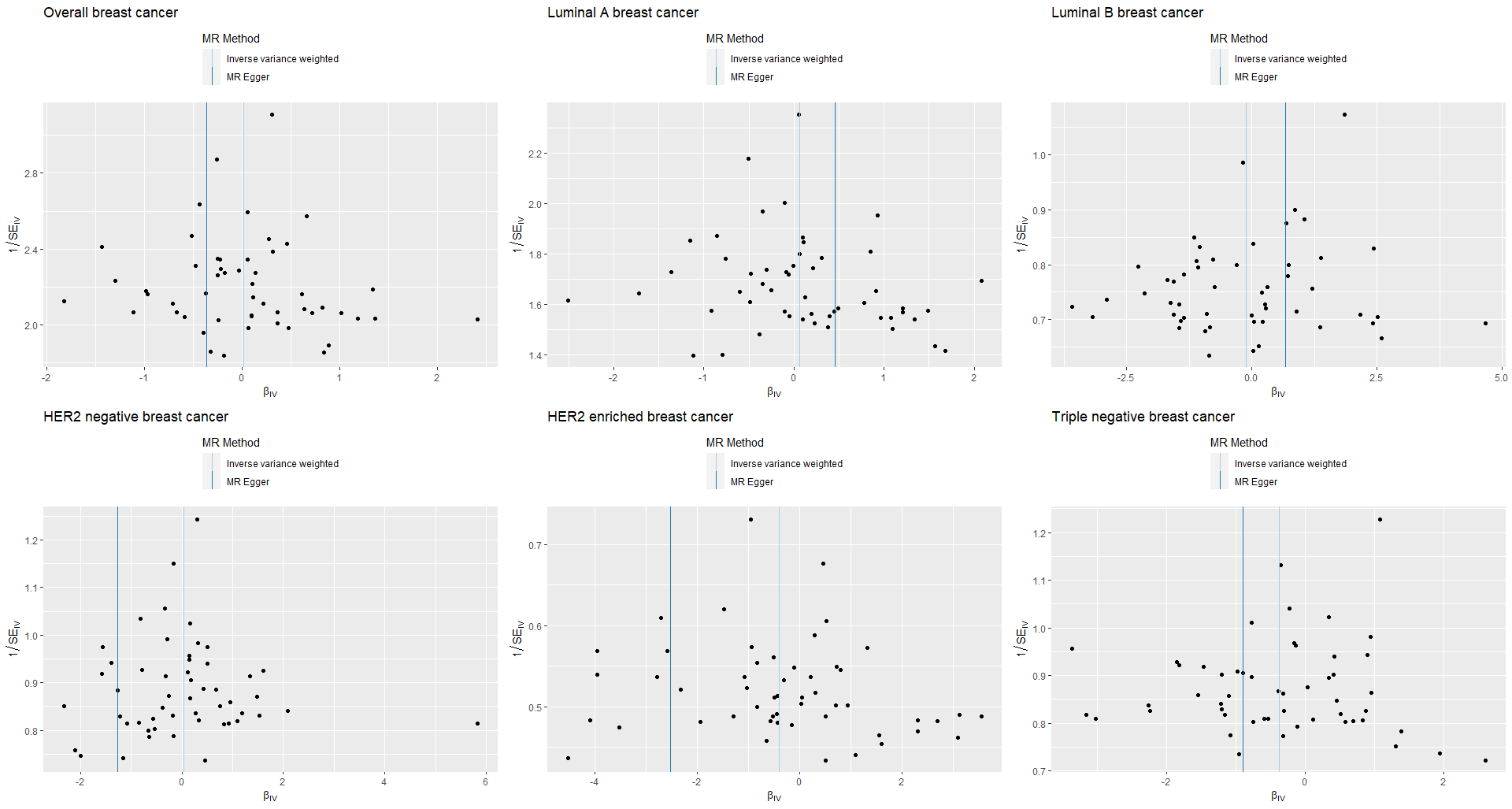

**Supplemental Figure 11**: Funnel plots of risk estimates of leisure computer use and colorectal cancer against instrumental strength

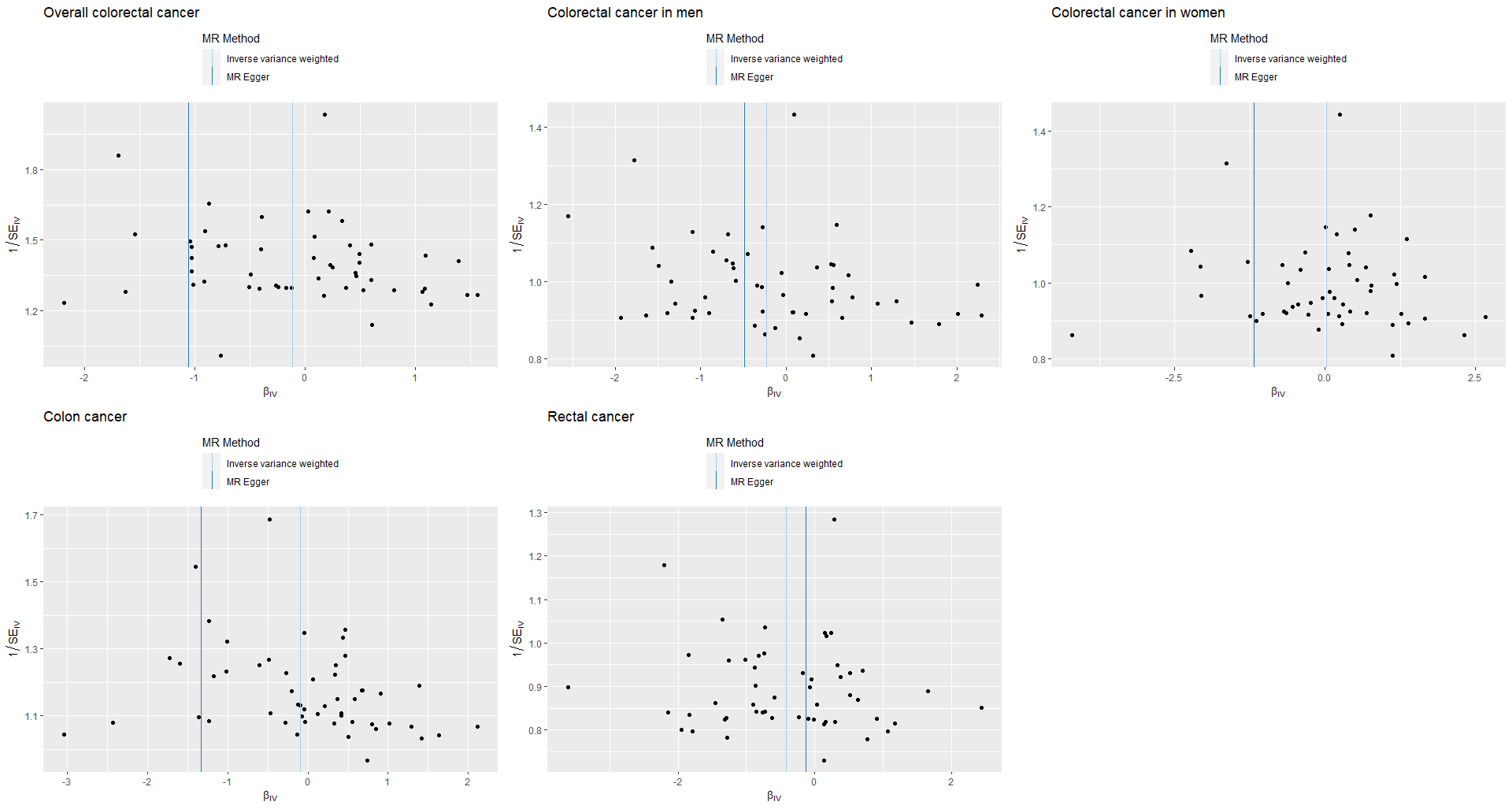

**Supplemental Figure 12**: Funnel plots of risk estimates of leisure computer use and prostate cancer against instrumental strength

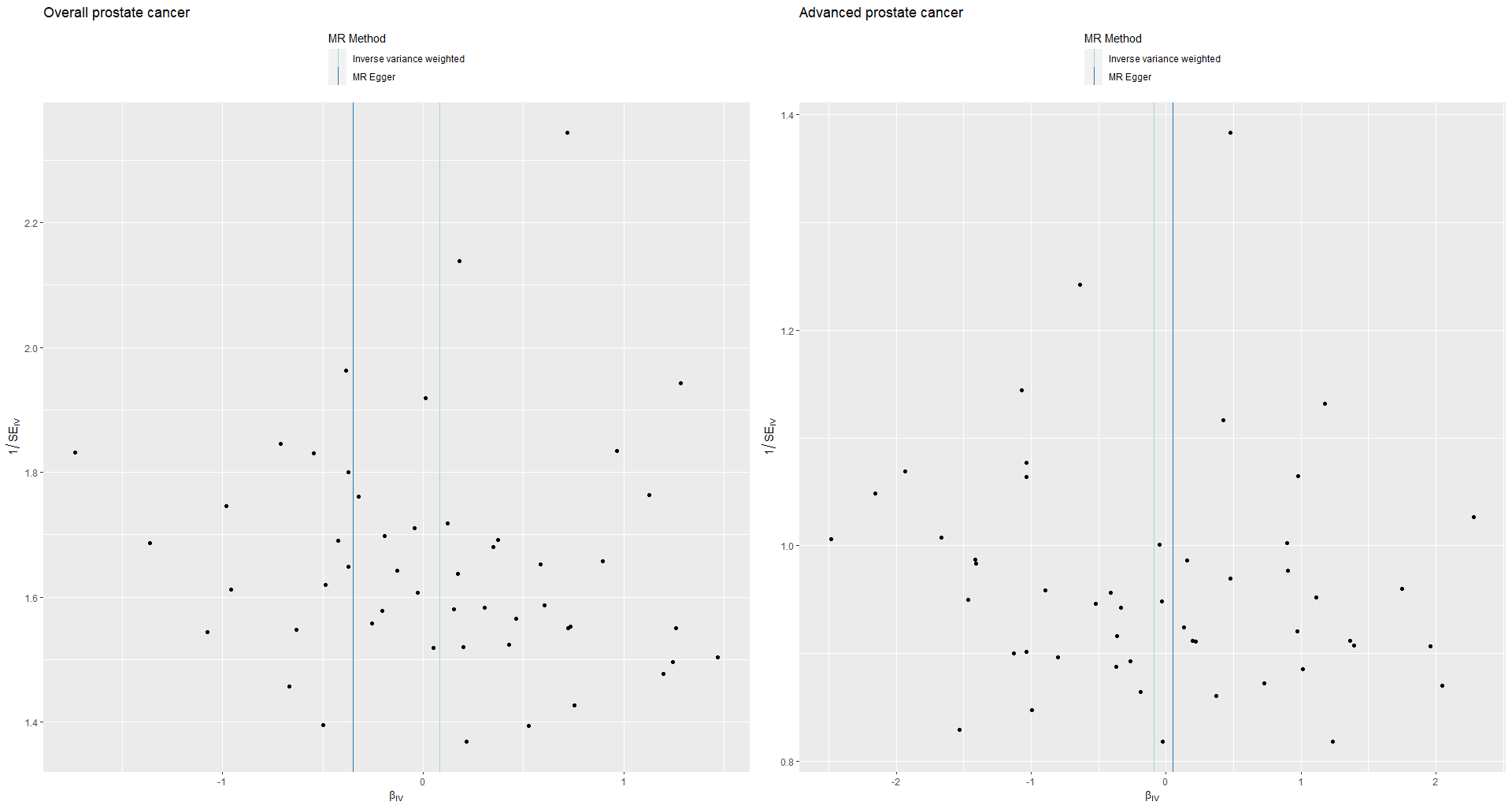
